# Use of AI-Related CPT Codes by Patient, Geographic, and Payer Characteristics in the U.S., 2021-2026

**DOI:** 10.64898/2026.11.09.26367689

**Authors:** Eileen Kim, Sophie Chang, Jean Feng, Poorva Satish Bedmutha, Lily Poursoltan, Jie Cao, Alexandra S. Wellsjo, Eric Floyd, Danqi Luo, Kevin Zhu, Aaron Boussina, Jennifer Holland, Amy M. Sitapati, Shahin Hallaj, Shamim Nemati, Rodney A. Gabriel, Alexander A. Khalessi, Christopher A. Longhurst, Karandeep Singh, Mattheus Ramsis

**Affiliations:** Division of General Internal Medicine, Department of Medicine, University of California, San Diego, California, USA; Joan & Irwin Jacobs Center for Health Innovation, University of California San Diego Health, San Diego, California, USA; School of Medicine, University of California, San Diego, La Jolla, California, USA; Department of Epidemiology and Biostatistics, University of California, San Francisco, California, USA; Bakar Computational Health Sciences Institute, University of California, San Francisco, California, USA; Electrical and Computer Engineering Department, University of California, San Diego, California; Rady School of Management, University of California, San Diego, California, USA; Information Services, University of California San Diego Health, San Diego, California, USA; Division of Biomedical Informatics, Department of Medicine, University of California, San Diego, California, USA; Division of Cardiology, Department of Medicine, University of California, San Diego, California, USA; Division of Perioperative Informatics, Department of Anesthesiology, University of California, San Diego, California, USA; Department of Neurosurgery, University of California, San Diego, CA, USA; Center for Clinical & Translational Research, Seattle Children’s Hospital, Seattle, WA, USA

## Abstract

**Importance:** Payment policy for clinical artificial intelligence (AI) is changing quickly, yet national data on which AI-enabled services are used, and for whom, are limited.

**Objective:** To describe national trends in the use of AI-related Current Procedural Terminology (CPT) codes and how use differs by patient, geographic, and payer characteristics.

**Design, Setting, and Participants:** This serial cross-sectional study used Epic Cosmos, a deidentified electronic health record dataset, to examine 11,605,867,309 encounters among 233,628,292 US patients from January 1, 2021, to April 14, 2026.

Exposures: Calendar year, age, sex, race, ethnicity, rurality, area-level social vulnerability, census region, and primary payer.

**Main Outcomes and Measures:** Encounters with any of 65 CPT codes that require AI, identified by matching the CPT code set to US Food and Drug Administration-listed AI-enabled devices and classified as assistive, augmentative, or autonomous using the CPT Appendix S taxonomy.

**Results:** Of 11.6 billion encounters, 335,349 (2.9 per 100,000) included an AI-required CPT code. The odds of an encounter with an AI-required code were 9.88 (95% CI, 9.71-10.06) times higher in 2025 than in 2021. In unadjusted comparisons, use was higher among patients aged 65 to 85 years and among those with Medicare (OR, 5.11) or Medicare Advantage (OR, 5.12) vs commercial insurance. Use was lower among Black (OR, 0.72) and Hispanic or Latino (OR, 0.34) patients, Medicaid enrollees (OR, 0.65), residents of the most socially vulnerable areas (OR, 0.53), and residents of the most rural areas (OR, 0.86). Use of automated diabetic retinopathy screening increased after the service was added to a HEDIS quality measure. One newer code (quantitative brain MRI) appeared in rural areas within months of becoming effective, whereas older codes such as retinopathy screening took years.

**Conclusions and Relevance:** In this cross-sectional study, billed use of AI-enabled services remained rare but grew substantially and was concentrated among older and Medicare-insured patients, with lower use among Medicaid enrollees and residents of socially vulnerable areas. As the Centers for Medicare & Medicaid Services restructures payment for software and launches outcome-based models, tracking who receives AI-enabled care across payment pathways will be important for equitable access.

**KEY POINTS:** *Question:* How has billed use of artificial intelligence (AI)-related CPT codes changed nationally since 2021, and how does use differ by patient, geographic, and payer characteristics?

*Findings:* In this cross-sectional study of 11.6 billion US encounters, 2.9 per 100,000 included an AI-required CPT code. Odds of use were nearly 10 times higher in 2025 than in 2021, higher among Medicare beneficiaries, and lower among Medicaid enrollees and residents of socially vulnerable areas.

*Meaning:* Billed AI use remains rare and uneven across payers and communities, so payment policy for AI may need to be paired with monitoring of who receives AI-enabled care.

## INTRODUCTION

National adoption of clinical artificial intelligence (AI) tools spans applications in screening, diagnosis, treatment, risk stratification, and workflow automation.^1^ Uptake of clinical AI has previously been characterized through a combination of public and proprietary claims databases and health system surveys.^1–3^ These studies have collectively found imaging and radiology applications to be the most mature area of clinical adoption among health systems, with AI devices assessing coronary artery disease and diabetic retinopathy generating the highest number of claims. AI-related claims are an increasing source of revenue for health systems, with nearly 50% of radiology AI claims to Medicare reimbursed between 2018 and 2023 with year-over-year growth.^2^ Since then, the number of FDA-approved AI medical devices has nearly doubled from 726 to 1544,^4^ with 23 new AI-related CPT codes becoming available.^5^ Recent policy changes have also driven the availability of autonomous AI, with AI-driven fractional flow reserve (FFR) calculation becoming the first software-as-a-service CPT code in 2024,^6^ and diabetic retinopathy becoming a HEDIS measure alongside the availability of autonomous AI tools that can fulfill this measure.^7^

While this growth is widely recognized, less is understood about the use of clinical AI CPT codes in real-world practice at a national level, and their patterns of diffusion after new CPT codes become available. Prior work suggests that adoption has been uneven. Health systems have been slow to implement AI tools due to concerns around clinical or financial value,^8^ and lack of reimbursement has further limited uptake despite widespread technical availability.^9^ The presence of geographic disparities in AI readiness and implementation capacity may also limit use, with metropolitan areas having higher rates of adoption as compared to rural settings.^10^ Federal policy now targets this gap. The Rural Health Transformation Program, launched in 2026, directs $50 billion through the Centers for Medicare & Medicaid Services (CMS) to rural health care, including technology adoption. Whether new AI services reach rural patients, and how quickly, is therefore a policy-relevant question. ^11^

Most AI services enter CPT as temporary Category III codes. These codes carry no national relative value units, are priced by individual Medicare Administrative Contractors, and are often not covered by commercial payers^12^. Only after showing widespread use and clinical efficacy can a service move to a permanent Category I code with national payment under the Medicare Physician Fee Schedule. Hospitals are paid differently. Inpatient AI can receive a new technology add-on payment (NTAP) on top of diagnosis-related group payments^13^. Outpatient services can be placed in New Technology Ambulatory Payment Classifications, and CMS has proposed handling algorithm-based services as a separate category called “software as a medical service”^14^. Most recently, the CMS ACCESS Model, which began in July 2026, pays organizations for technology-enabled chronic care on the basis of patient outcomes rather than individual services.^15^ Several of these pathways are changing at once. CMS will end the NTAP alternative pathway for breakthrough devices beginning with fiscal year 2028 applications, a change expected to affect US clinical AI innovation.^13,14^

Monitoring the reach of these initiatives requires a mechanism to monitor AI adoption nationally and throughout our communities. In this study, we describe national use of AI-related CPT codes from 2021 to 2026 and how use differs by age, sex, race, ethnicity, rurality, area-level social vulnerability, region, and payer. We also describe how use changed after codes became payable or tied to quality measures, and how quickly new codes reached rural areas.

## METHODS

This study follows the Strengthening the Reporting of Observational Studies in Epidemiology (STROBE) reporting guidelines for cross-sectional studies. The University of California San Diego Institutional Review Board determined that this study was not human subjects research because it used deidentified limited data.

### Data Source and Inclusion Criteria

Data used in this study came from Epic Cosmos, a deidentified dataset created in collaboration with a community of health systems using the Epic electronic health record, representing more than 300 million patient records from over 2067 hospitals and 47,100 clinics as of May 2026. All analyses used data from patients in the United States.

We included encounters from January 1, 2021 through April 14, 2026. We queried from the patient and encounter data models. Of the 88 AI-associated CPT codes we identified, 65 appeared in Cosmos and formed the basis of our analysis.

Billed procedures were extracted along with a set of variables chosen to capture demographic, geographic, and socioeconomic variation: age, race, ethnicity, rural-urban commuting area designation, social vulnerability index percentile, census region, state of residence, and primary payer financial class.

### Creating a Comprehensive Database of CPT Codes

To build a list of CPT codes associated with AI, we used a multi-step approach. We defined an AI-related CPT code as a code whose descriptor or associated FDA-listed device uses an algorithm that analyzes clinical data to produce an output beyond a summation of the input, consistent with the CPT Appendix S taxonomy. We searched the CPT code book for terms that frequently appear in AI-related billing: “autonomous”, “augmentative”, “assistive”, “automated”, “automatic”, “algorithmic”, and “algorithm”. We also compiled descriptors of all 1,544 FDA 510(k) devices building on prior work^16^, filtered to AI-enabled devices, and matched each to its most plausible CPT code through researching the CPT code book, vendor materials, and literature reviews. Each pair was reviewed by two authors, with differences adjudicated by a third author. Large language model-based queries were used to identify additional candidates. Because emerging technologies are often assigned a temporary Category III code prior to a permanent Category I code, the same procedure can be represented by multiple CPT codes over time; we grouped such codes with their successors. For instance, 0502T and 75580 represent the same procedure across different timeframes and were combined in our data.

We also evaluated 32 CPT codes from Wu *et al*. and 23 CPT codes from Zhang *et al*. for inclusion.^2,17^ In total, this process produced 88 AI CPT codes. Of these, 51 required the use of AI (primary analysis) and 37 were AI-optional (included in sensitivity analysis). AI-optional codes were ones without AI use described or alluded to in their definition in the CPT code book and have known associations with products that do and do not use AI. The complete listing of AI CPT codes is in **eTable 1**.

**Table 1.** Demographic breakdown of all patients with an AI-required CPT code from 1/1/2021 to 4/14/2026 (n = 284501). For certain variables, one patient could be counted multiple times. For example, if their primary payer changed within the timeframe, if they had multiple AI-required services at different ages, or if they reported multiple races.

| Characteristic | Value | AI-required CPT code use |
|---|---|---|
| **Age (years)** | <18 | 605 (0.21%) |
|  | [18, 30) | 3295 (1.15%) |
|  | [30, 40) | 8862 (3.11%) |
|  | [40, 50) | 22837 (8%) |
|  | [50, 65) | 94484 (33.11%) |
|  | [65, 75) | 96269 (33.74%) |
|  | [75, 85) | 51213 (17.95%) |
|  | ≥85 | 7800 (2.73%) |
|  | No value | 0 (0%) |
| **Sex** | Female | 129974 (45.68%) |
|  | Male | 152293 (53.53%) |
|  | None of the above | 2234 (0.79%) |
| **Race** | American Indian or Alaska Native | 2932 (0.91%) |
|  | Asian | 11711 (3.62%) |
|  | Black or African American | 32879 (10.16%) |
|  | Native Hawaiian or Other Pacific Islander | 1225 (0.38%) |
|  | Other Race | 40082 (12.38%) |
|  | White | 199412 (61.6%) |
|  | None of the above | 35477 (10.96%) |
| **Ethnicity** | Hispanic or Latino | 17346 (6.1%) |
|  | Not Hispanic or Latino | 225516 (79.27%) |
|  | None of the above | 41639 (14.64%) |
| **US census region** | Midwest | 78411 (27.56%) |
|  | Northeast | 50658 (17.81%) |
|  | South | 125353 (44.06%) |
|  | West | 29936 (10.52%) |
|  | None of the above | 143 (0.05%) |
| **RUCA** | 1 Metropolitan area core | 214447 (75.38%) |
|  | 2-3 Metropolitan commuting | 28445 (10%) |
|  | 4-6 Micropolitan | 22586 (7.94%) |
|  | 7-9 Small town | 10630 (3.74%) |
|  | 10 Rural areas | 5961 (2.1%) |
|  | None of the above | 2432 (0.85%) |
| **SVI** | <25% | 63882 (22.45%) |
|  | [25%, 50%) | 49642 (17.45%) |
|  | [50%, 75%) | 40869 (14.37%) |
|  | ≥75% | 32611 (11.46%) |
|  | No value | 97497 (34.27%) |
| **Primary payer** | Commercial | 151980 (33.16%) |
|  | Medicaid | 58178 (12.69%) |
|  | Medicare | 110420 (24.09%) |
|  | Medicare Advantage | 83743 (18.27%) |
|  | VA/Tricare | 6469 (1.41%) |
|  | Other | 47557 (14.33%) |
| **Year** | 2021 | 13301 (4.54%) |
|  | 2022 | 19436 (6.64%) |
|  | 2023 | 31832 (10.87%) |
|  | 2024 | 58088 (19.84%) |
|  | 2025 | 122287 (41.77%) |
|  | 2026 (to April 14) | 47806 (16.33%) |

### Maturity of AI Use

AI use was determined to be assistive, augmentative, or autonomous using prior definitions published by the American Medical Association’s CPT Appendix S.^5^ Assistive AI refers to software that provides clinically relevant data without generating an interpretation or providing conclusions. Augmentative AI refers to software that analyzes or quantifies data to produce a clinically meaningful output that represents more than a summation of the source data. Autonomous AI refers to software that independently generates clinically meaningful interpretations, with degrees of automation ranging from providing recommendations (Level I) to initiating management actions (Level III).^5^

### Diffusion of AI from Urban to Rural Areas

For a subset of AI CPT codes that were new and grew in use within the study’s timeframe, adoption was visually assessed over time and across RUCA scores to characterize how usage of the CPT code diffused from urban to rural areas indexed to the availability of the code. This analysis was descriptive. These CPT codes were: 92229 for automated retinopathy screening, 0865T-0866T for quantitative MRI of the brain, 0623T-0626T for plaque analysis on coronary CTA, 0932T for heart failure detection on echocardiogram.

### Statistical Analyses

Because the aim was descriptive, we report unadjusted odds ratios. Differences across groups reflect both clinical need for the underlying services and access to them. Because the outcome was rare, odds ratios closely approximate relative risks. Unadjusted odds ratios were used to compare the likelihood of AI use across demographic categories, with 95% confidence intervals calculated using the Wald method. Analyses were performed at the patient level, with demographic categories compared against the outcome of patient exposure to any CPT code indicative of AI use during any encounter. Patients were counted multiple times if they had multiple encounters across different variables, e.g. age/year, insurance payer, or moving to a different region of the U.S. Reference groups were: age 50 to 65 years, female, White race, Not Hispanic or Latino, Midwest US census region, rural-urban commuting area (RUCA) of 1 Metropolitan area core, social vulnerability index (SVI) <25%, and commercial primary payer. Social vulnerability was measured with the CDC SVI for the patient’s area of residence by census tract. Data visualizations over time accounted for the overall increasing number of encounters captured in Cosmos by showing proportions rather than raw counts. Missing data were excluded from analyses involving the variable in question.

### Sensitivity Analysis

In addition to recently introduced CPT codes that are definitive indicators of AI use, there are a set of older CPT codes for which AI technology has subsequently become available or more accurate (for example, computer-aided mammography). Use of these CPT codes may indicate AI use but is not definitive. We refer to these codes as “AI-optional” since the use of AI within these codes is optional. As a sensitivity analysis, we repeated our primary analysis with these AI-optional codes included.

### Software

All analyses were conducted in R version 4.5.1. Data visualization was done using ggplot2 in R. An interactive Health AI Adoption Tracker is available online at https://health-ai-adoption-tracker.web.app/review.

## RESULTS

Of 233,628,292 patients and 11,605,867,309 encounters between January 1, 2021 and April 14, 2026, there were 335,349 encounters that involved an AI CPT code (2.9 per 100,000 encounters) among 284,501 patients (0.12% of patients). Among patients that had encountered an AI CPT code, most were between 50 and 75 years old (67%), male (54%), white (62%), not Hispanic or Latino (79%), and live in the most urban RUCA (75%). Medicare or Medicare Advantage was the most common group of insurance among patients receiving an AI-required CPT code, representing 42% of the cohort (**Table 1**).

Overall, use of AI has increased nationally between 2021 and 2026 (**Figure 1)**. The odds of an encounter with an AI-required code were 9.88 (95% CI, 9.71-10.06) times higher in 2025 than in 2021 (**Figure 2**). The increase reflected growth in assistive, augmentative, and autonomous AI codes, and several increases followed payment or quality measure changes (**Figure 3**). For example, use of AI for retinopathy screening (CPT 92229) began to increase after changes to the Healthcare Effectiveness Data and Information Set (HEDIS) diabetic eye exam measure requiring explicit result, retiring the hybrid collection method as of measurement year 2025. A handful of category III codes, generally considered experimental, have transitioned to the more permanent Category I codes. There remain many AI codes that have near-zero usage, such as 0740T for autonomous algorithm-based insulin dose calculation.

**Figure 1.**
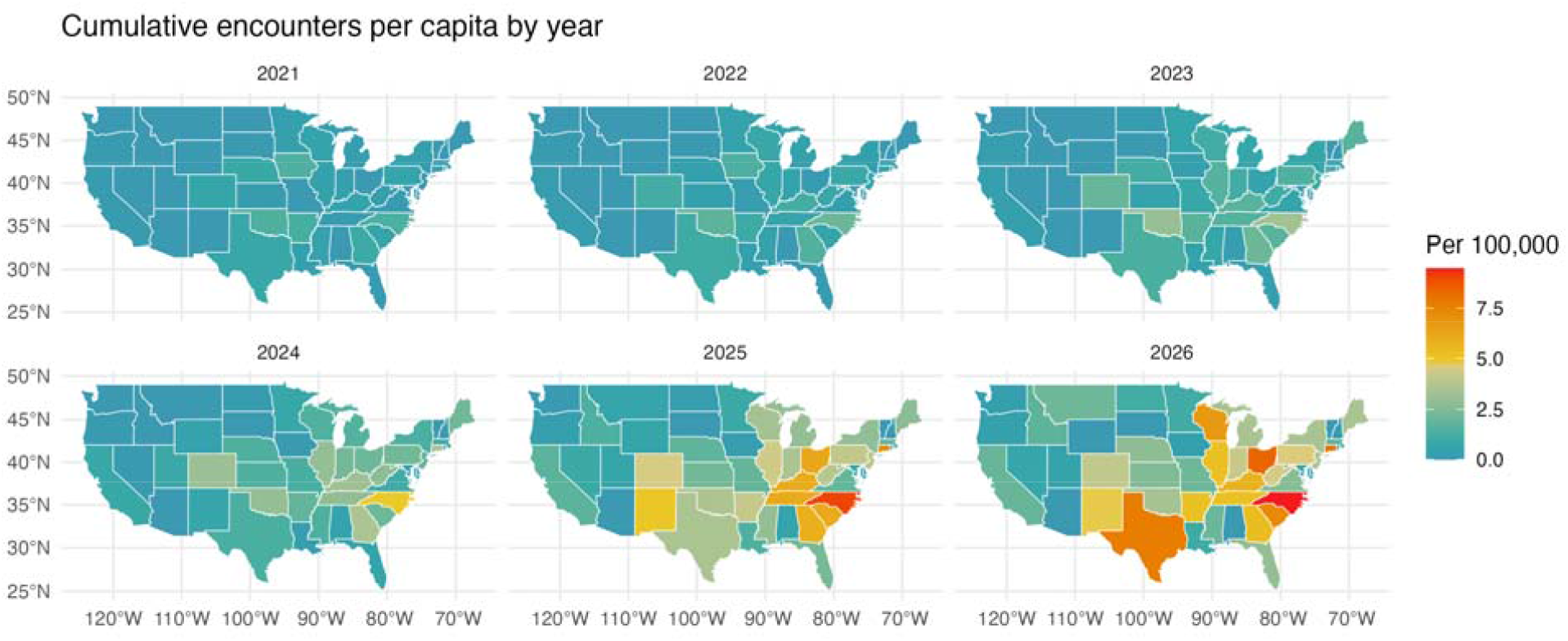
Heat map of the continental United States showing state-level encounters per capita (per 100,000 residents that have a record in Cosmos) that included an AI CPT code by year.

**Figure 2.**
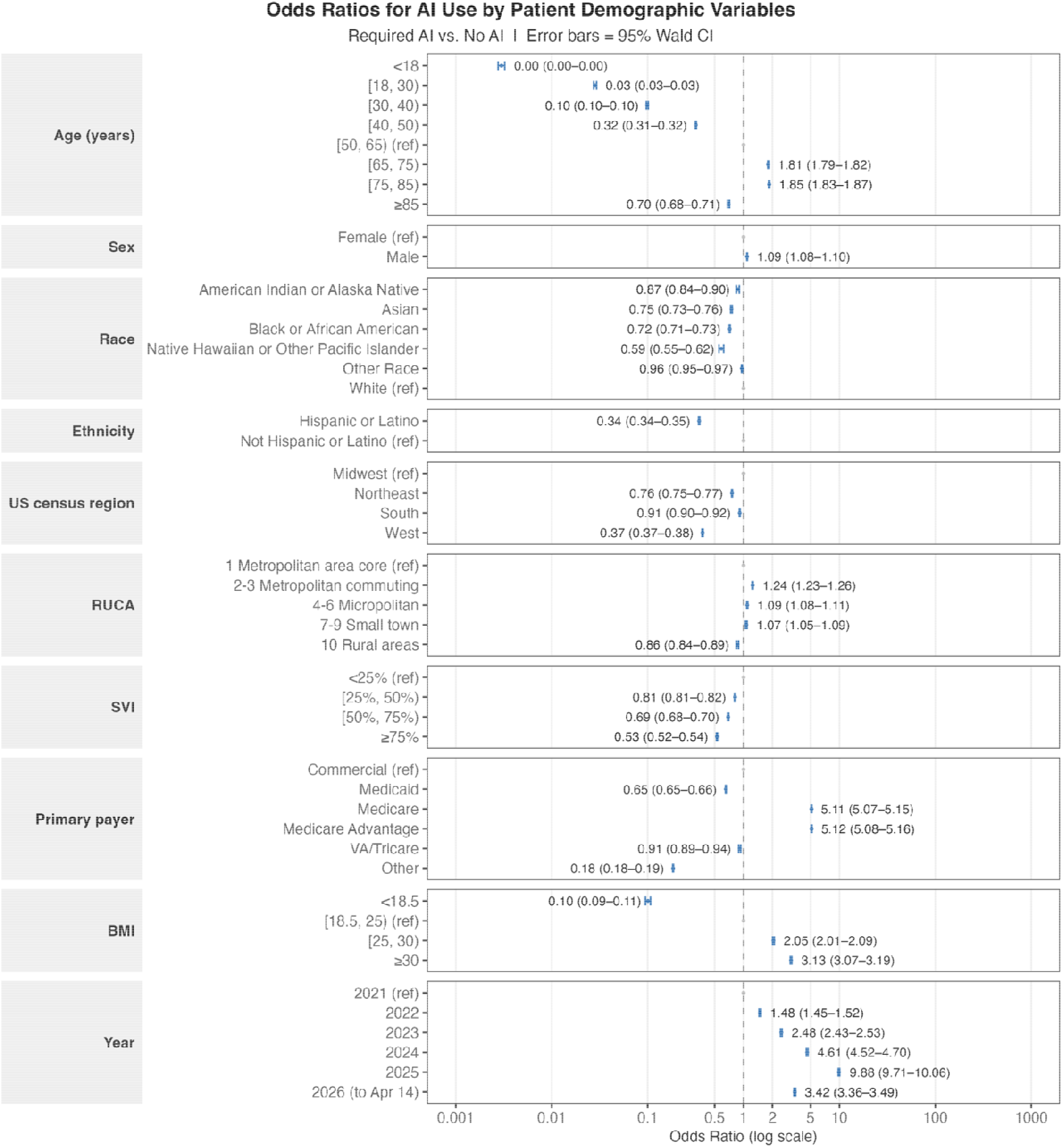
Odds ratios by demographic factors comparing patients with any encounter involving an AI-required CPT code versus no recorded AI use. Odds ratios > 1 are indicative of greater AI use. Patients were counted multiple times in these analyses if they had multiple encounters across different variables, e.g. age/year, insurance payer, or moving to a different region of the U.S.

**Figure 3.**
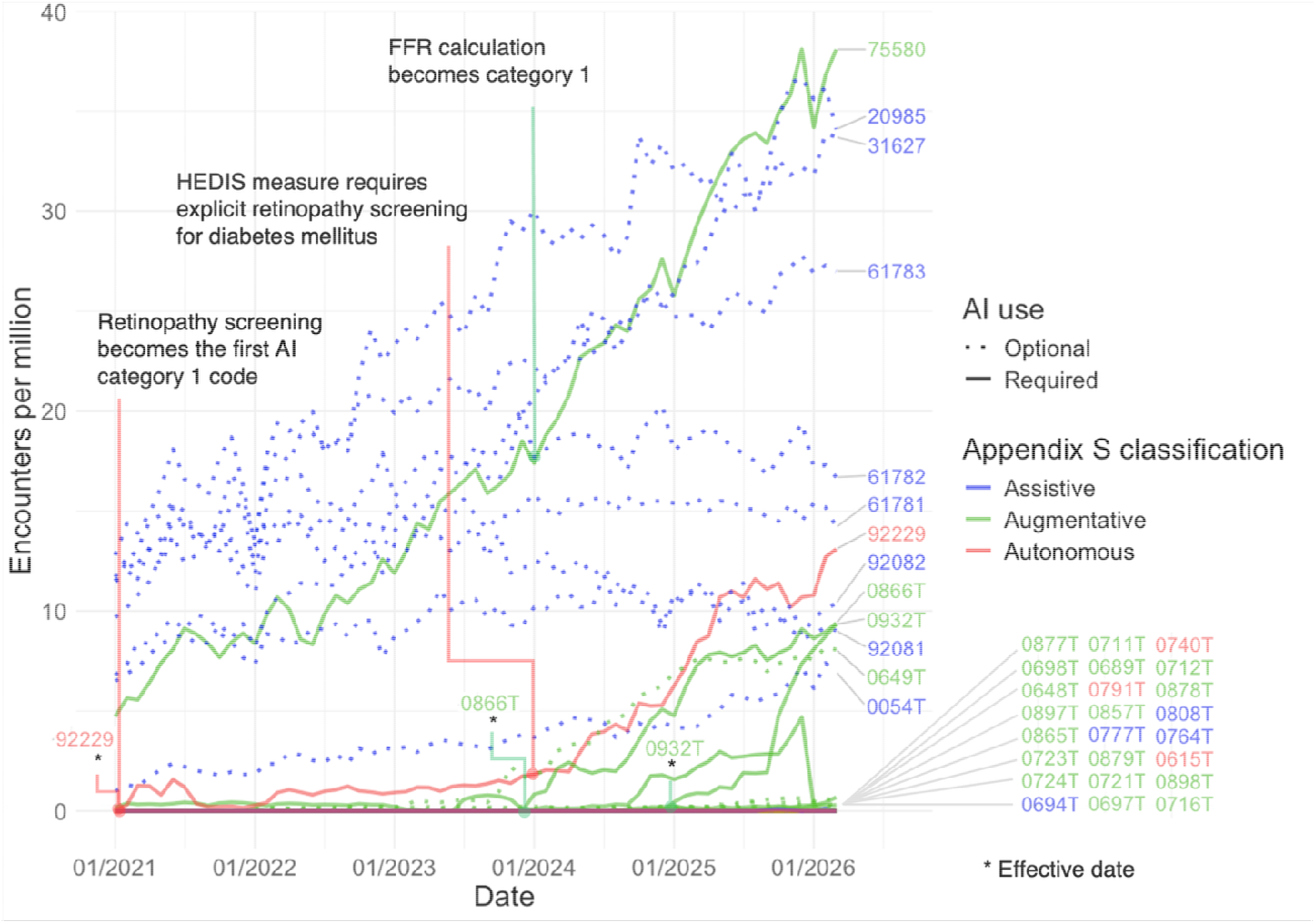
Proportion of encounters per million, per month, that included an AI-related CPT code in the Cosmos data set from 1/1/2021 to 3/30/2026. This plot excludes AI-optional CPT codes 85027, 95027, 77067, 88175, 92083, 93356 (with 0399T), 77280 (with 77285 and 77290), and 92227-92228 for complete blood counts with and without differentials, screening mammography, pap smear, visual field exams, myocardial strain imaging, therapeutic radiology simulation-aided field setting, and non-automated retinal imaging respectively, given their much greater scale. CPT 75580 (which is grouped with preceding codes 0501T-0504T) is used for coronary fractional flow reserve estimation from computed tomography angiography. CPT code 92229 is used for automated retinopathy screening. CPT code 0866T is used for quantitative MRI brain analyses. CPT code 0932T is used for heart failure detection on echocardiogram.

### Factors Associated with AI Adoption

Associations between demographic factors and AI adoption are described in **Figure 2**.

#### Age

With age [50, 65) as a reference, the highest AI adoption was among those aged [75, 85) (OR 1.85; 95% CI 1.83–1.87) and [65, 75) (OR 1.81; 95% CI 1.79–1.82), while the lowest adoption was among people under the age of 50 and people aged 85 and older.

#### Sex

Males had higher odds of AI use than females (OR 1.09; 95% CI 1.08–1.10).

#### Race and ethnicity

Non-White race was associated with a lower use of AI, including American Indian or Alaska Natives (OR 0.87), Asian (OR 0.75), Black or African Americans (OR 0.72), and Hawaiian/Pacific Islanders (OR 0.59). Hispanic or Latino ethnicity was also associated with lower use of AI (OR 0.34).

#### Region and rurality

As compared to the Midwest, all regions had a lower use of AI, including the West (OR 0.37), Northeast (OR 0.76), and South (OR 0.91). As compared to metropolitan areas, metropolitan commuting (OR 1.24), micropolitan (OR 1.09), and small town (OR 1.07) areas saw a higher use of AI, whereas rural areas saw a lower use of AI (OR 0.86).

#### Social vulnerability

*Compared with residents of the least socially vulnerable areas (SVI <25%), residents of more vulnerable areas had lower use of AI, most pronounced in the most vulnerable areas (SVI ≥75%; OR, 0*.*53)*.

#### Primary payer

As compared to commercial insurance, Medicare (OR 5.11) and Medicare Advantage (OR 5.12) had higher use of AI, whereas Medicaid (OR 0.65),VA/Tricare (OR 0.91), and other insurance (OR 0.18) were associated with lower use of AI.

### Diffusion of AI from Urban to Rural Areas

Looking at highly used CPT codes reveals patterns of diffusion from the most urban to the most rural areas (**Figure 4**). In the case of older codes, such as automated retinopathy and plaque analysis on coronary CT angiogram, the codes were used in primarily urban areas for years until recent use in rural areas. On the other hand, the CPT code quantitative MRI brain was introduced in January of 2024, with the code seeing use in rural areas within months. In the case of heart failure on echocardiogram, its use of the code in rural, small town, and micropolitan areas is disproportionately high as compared to overall encounters.

**Figure 4.**
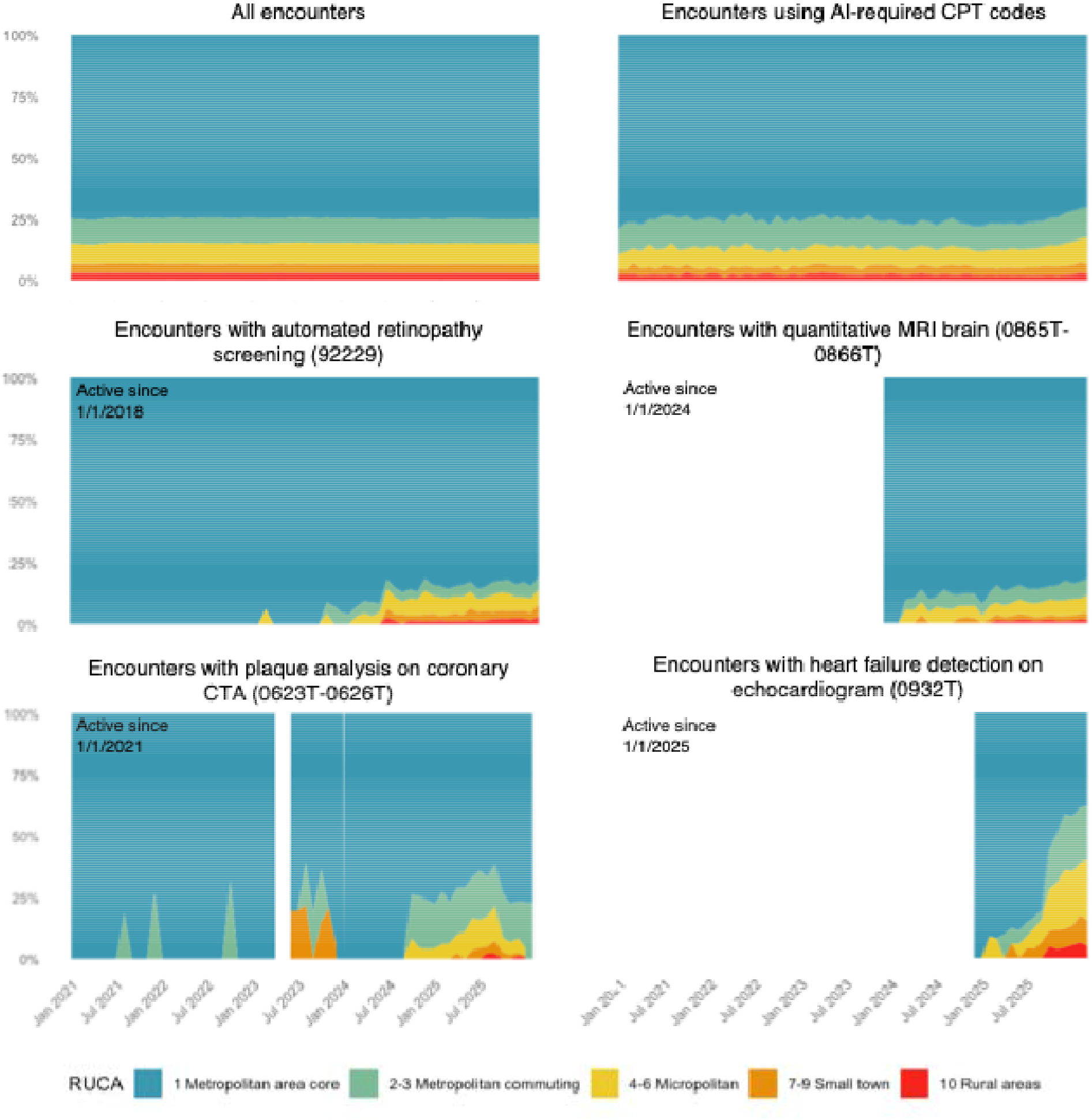
Breakdown of encounters that use a given AI-related CPT code per month by RUCA. Plots either begin at the start of our data on 1/1/2021 or the date that the CPT code became effective.

### Sensitivity Analysis

Inclusion of AI-optional CPT codes expanded the outcome of AI CPT use to 472,807,750 encounters representing 112,435,216 patients. The latter showed similar patterns across most variables, but with reduced magnitudes (**eFigure 1**). Notable differences were that males had lower odds of encountering an AI-required or AI-optional CPT code (OR 0.64), and certain racial minorities had higher odds of encountering these codes, including Black or African American patients (OR 1.06), American Indian or Alaska Native (OR 1.10), and Native Hawaiian or other Pacific Islander (OR 1.14). Higher SVI remained associated with lower use of AI, whereas rural areas had a higher use of AI-optional codes as compared with metropolitan areas.

## DISCUSSION

In this national analysis of AI CPT code use, we found that the use of assistive, augmentative, and autonomous AI in healthcare has risen since 2021. We found several social and geographic variables associated with the use of AI and lower use of AI among those with the highest SVI and those in the most rural (RUCA 10) areas. In contrast to prior work,^10^ use of several AI codes reached rural areas over time (**Figure 4**), although the lag ranged from a few months to several years after a code became effective. Lower use in the most socially vulnerable patients, however, persisted.

Our findings are consistent with the payment pathway affecting whether, and for whom, an AI service is used. Many Category III AI codes had near-zero use, such as 0740T for autonomous insulin dose calculation. This is consistent with how those codes are paid: contractor-specific pricing and frequent commercial noncoverage give clinicians little reason to adopt a tool. Yet moving to Category I requires evidence of widespread use, so an AI tool cannot show broad use without payment and cannot get predictable payment without broad use. The largest increases in use coincided with a service becoming both paid and measured. FFR-CT use appeared to increase after Category I conversion (**Figure 3**). Autonomous retinopathy screening grew only modestly under national payment, then more than doubled within a year after a HEDIS quality measure created demand from health plans (**Figure 3**). Even Category I payment remains unsettled, because the fee schedule’s practice expense method was not built for software bought by subscription or per use, and CMS has asked for comment on a consistent payment approach across care settings.^18,19^ The proposed “software as a medical service” policy would place outpatient AI in New Technology APCs while CMS develops a permanent method.^14^ Pairing that interim payment with requirements to report outcomes could help promising Category III services build the evidence they need for permanent valuation.

Payment pathways outside the CPT system raise separate concerns about access and measurement. NTAPs apply only to hospitals paid under the inpatient prospective payment system. Critical access hospitals, which Medicare pays on the basis of reasonable cost, are excluded.^20,21^ Recent AI NTAPs have also been much smaller than the first one, at $241.39 per case in fiscal year 2025 and $137.53 in fiscal year 2027.^21,22^ Once the alternative pathway ends, AI developers will have to show substantial clinical improvement to qualify. Wong and colleagues warn that this change will affect US clinical AI innovation, and it may narrow the main inpatient payment route for AI just as more tools reach the market.^13^ ACCESS moves in a different direction: it pays for outcomes that algorithms may help achieve rather than for the algorithms themselves.^15^ This design may discourage low-value use, but it carries equity risks. Organizations may favor patients who are most likely to meet targets. The model initially covers only Original Medicare, so it excludes Medicaid enrollees, who in our data already had lower AI use. Neither inpatient AI paid through NTAPs nor AI funded through ACCESS shows up in CPT-based data. Public reporting of AI use within these pathways, broken down by patient and community characteristics, would be needed to tell whether new payment policies are narrowing or widening gaps in access.

Our report has limitations. Our study includes only patients in Epic’s Cosmos, which may overrepresent larger health systems and miss patients who receive care in a setting with a different electronic health record or those who did not receive care. The Cosmos data set included 65 of 88 identified AI-related CPT codes, which could indicate that certain codes were not used or were not present in the data. We found variable levels of data missingness; for example, 34% of patients do not have an SVI, 11% do not have a race documented, 15% do not have ethnicity documented (**Table 1**). Comparisons were unadjusted, so differences by age, payer, and region partly reflect clinical need for the underlying services, and Medicare differences partly reflect age. We did not distinguish care settings, so we cannot separate inpatient, outpatient, and ambulatory surgery use, which are paid through different pathways. Temporal associations with payment and quality measure changes do not establish that those changes caused the increases. Lastly, AI CPT codes are merely a proxy for overall AI use and may overrepresent higher-risk use cases (e.g., autonomous AI) with separate billing mechanisms.

Billed use of AI-enabled services in the U.S. is growing quickly from a very low base and remains uneven across payers and communities. As payment for AI moves toward new pathways, including software-specific payment and outcome-based models such as ACCESS, monitoring use by patient and community characteristics will help show whether these policies widen or narrow gaps in access.

## Supporting information

eFigure 1

eTable 2

eTable 1

## Data Availability

Cosmos data are available to Cosmos participating organizations under Epic's terms. The compiled CPT code list can be found in eTable 1.

## ACKNOWLEDGEMENTS

The authors thank Hanyang Li and Xiao He, for generating the FDA device descriptors that helped us in capturing AI-related CPT codes more completely. The authors also thank Phil Lindeman and the Epic Cosmos team for helping to establish our data queries and for building the Cosmos database for research. AI-related tools used to supplement our CPT code database were Anthropic’s Claude Sonnet 4.6, OpenAI’s ChatGPT 5.3, and Google’s Gemini 3 Flash, from February 2026 through April 2026. Anthropic’s Claude Opus 5.5 was used to edit the manuscript in October 2026. The authors take responsibility for the integrity of the content generated.

