## Supplementary material for "Use of AI-Related CPT Codes by Patient, Geographic, and Payer Characteristics in the U.S., 2021-2026": eFigure 1

**eFigure 1.** Odds ratios by demographic factors comparing patients with any encounter involving an AI-required or AI-optional CPT code versus no recorded AI use. Odds ratios > 1 are indicative of greater AI use.

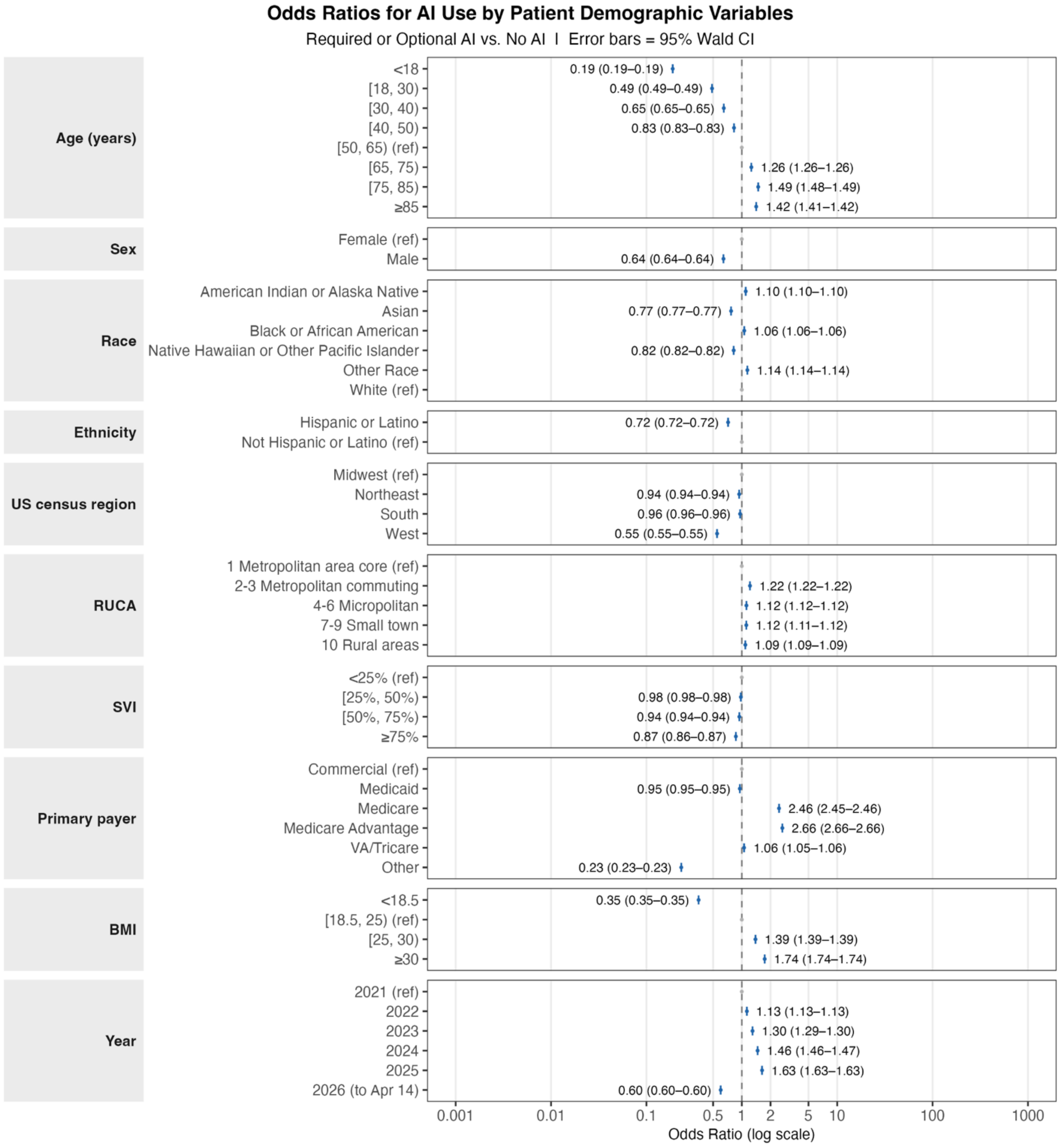
