## Supplementary material for "Use of AI-Related CPT Codes by Patient, Geographic, and Payer Characteristics in the U.S., 2021-2026": eTable 2

**eTable 2.** Diagnoses included in the diagnosis groupers for hypertension, dysplipidemia, and heart disease.

| Hypertension | Dyslipidemia | Heart disease |
| --- | --- | --- |
| Accelerated essential hypertension | Abnormally low high density lipoprotein (HDL) cholesterol with hypertriglyceridemia | 2-vessel coronary artery disease |
| Accelerated hypertension | Acquired hyperlipoproteinemia | 3-vessel CAD |
| Accelerated hypertension with diastolic congestive heart failure, NYHA class 3 | Acquired hypertriglyceridemia | 3-vessel coronary artery disease |
| Accelerated hypertension with NYHA class 1 diastolic congestive heart failure | Alpha-lipoproteinemia | Accelerated coronary artery disease in transplanted heart |
| Accelerated hypertension with NYHA class 4 systolic heart failure | Alphalipoproteinemia | Acute diastolic heart failure secondary to coronary artery disease |
| Accelerated secondary hypertension | Anapolipoproteinemia | Acute on chronic diastolic heart failure due to coronary artery disease |
| Acute spont intraparenchymal hemorrhage assoc w/ hypertension | Apo-c-II deficiency | Acute on chronic diastolic heart failure secondary to coronary artery disease |
| Acute spont subarachnoid intracranial hemorrhage assoc w/ hypertension | APOB-related hypercholesterolemia, autosomal dominant | Angina concurrent with and due to arteriosclerosis of autologous arterial coronary artery bypass graft |
| Acute spontaneous intraparenchymal intracranial hemorrhage associated with hypertension | Apolipoprotein C-III deficiency associated with mutation in APOC3 gene | Angina concurrent with and due to arteriosclerosis of CABG |
| Acute spontaneous intraventricular hemorrhage assoc w/ hypertension | Apolipoprotein E deficiency | Angina concurrent with and due to arteriosclerosis of coronary artery |
| Acute spontaneous intraventricular hemorrhage associated with hypertension | Autosomal dominant hypercholesterolemia associated with APOE p.Leu167del mutation | Angina concurrent with and due to arteriosclerosis of coronary artery bypass graft |
| Acute spontaneous subarachnoid intracranial hemorrhage associated with hypertension | Autosomal dominant hypercholesterolemia associated with mutation in APOB gene | Angina due to arteriosclerosis of autologous vein coronary artery bypass graft |
| Adams-Nance tachycardia, hypertension, microphthalmos, and hyperglycinuria syndrome (CMS/HCC) | Autosomal dominant hypercholesterolemia associated with mutation in LDLR gene | Angina due to arteriosclerosis of coronary artery of transplanted heart |
| Adrenal hypertension | Autosomal dominant hypercholesterolemia associated with mutation in PCSK9 gene | Arteriosclerosis of arterial coronary artery bypass graft |
| AGTR1-related essential hypertension | Autosomal dominant hyperlipoproteinemia type 3 | Arteriosclerosis of autologous arterial coronary artery bypass graft |
| Alteration in activity associated with pre-eclampsia | Autosomal recessive hypercholesterolemia | Arteriosclerosis of autologous arterial coronary artery bypass graft, angina presence unspecified |
| Alteration in activity due to pre-eclampsia | Autosomal recessive hyperlipoproteinemia type 3 | Arteriosclerosis of autologous arterial coronary artery bypass graft, unspecified whether angina present |
| Antepartum hypertension | Beta-sitosterolemia | Arteriosclerosis of autologous coronary artery bypass graft |
| Antepartum hypertension, first trimester | Borderline hypercholesterolemia | Arteriosclerosis of autologous vein coronary artery bypass graft |
| Antepartum hypertension, second trimester | Borderline hyperlipidemia | Arteriosclerosis of bypass graft of coronary artery |
| Antepartum hypertension, third trimester | Broad beta disease | Arteriosclerosis of coronary artery |
| Antepartum hypertension, unspecified trimester | Broad- or floating-betalipoproteinemia | Arteriosclerosis of coronary artery bypass graft |
| Antepartum hypertensive vascular disease | Buerger-Gruetz syndrome | Arteriosclerosis of coronary artery bypass graft of transplanted heart |
| Antepartum hypertensive vascular disease, first trimester | Burger-Grutz disease | Arteriosclerosis of coronary artery in patient with history of myocardial infarction |
| Antepartum hypertensive vascular disease, second trimester | Burger-Grutz syndrome | Arteriosclerosis of internal mammary artery coronary artery bypass graft |
| Antepartum hypertensive vascular disease, third trimester | Carbohydrate induced hyperlipoproteinemia | Arteriosclerosis of nonautologous biological coronary artery bypass graft |
| Antepartum hypertensive vascular disease, unspecified trimester | Chylomicronemia syndrome | Arteriosclerosis of nonautologous coronary artery bypass graft |
| Antepartum mild pre-eclampsia | Combined fat and carbohydrate induced hyperlipemia | Arteriosclerosis of nonautologous coronary artery bypass graft without angina pectoris |
| Antepartum mild preeclampsia | Combined heterozygous LDLR and heterozygous LDLRAP1 gene mutations | Arteriosclerotic coronary artery disease |
| Antepartum mild preeclampsia, second trimester | Combined hyperlipidemia | Arteriosclerotic heart disease (ASHD) |
| Antepartum mild preeclampsia, third trimester | Combined hyperlipidemia associated with type 2 diabetes mellitus | Arteriosclerotic heart disease |
| Antepartum mild preeclampsia, unspecified trimester | Congenital disorder of glycosylation type 2P (CMS/HCC) | Artscl ntv coron trnspl heart |
| Antepartum renal hypertension | Diet-controlled hyperlipidemia | ASHD (arteriosclerotic heart disease) |
| Antepartum renal hypertension in first trimester | Disorder of lipoprotein metabolism | Asymptomatic arteriosclerosis of coronary artery |
| Antepartum renal hypertension in second trimester | Disorder of lipoprotein metabolism, unspecified | Asymptomatic coronary artery disease |
| Antepartum renal hypertension in third trimester | DM type 1 with diabetic mixed hyperlipidemia | Asymptomatic coronary heart disease |
| Antepartum renal hypertension, first trimester | DM type 2 with diabetic mixed hyperlipidemia | Atheroscler of coronary artery bypass graft w/unstable angina pectoris |
| Antepartum renal hypertension, second trimester | Drug-induced diabetes mellitus with hyperlipidemia | Atherosclerosic heart disease of native coronary artery with refractory angina pectoris |
| Antepartum renal hypertension, third trimester | Dysbetalipoproteinemia | Atherosclerosis of arterial coronary artery bypass graft |
| Antepartum renal hypertension, unspecified trimester | Dyslipidemia, goal LDL below 100 | Atherosclerosis of autologous artery coronary artery bypass graft |
| Antepartum transient hypertension | Dyslipidemia, goal LDL below 130 | Atherosclerosis of autologous artery coronary artery bypass graft with angina pectoris |
| Antepartum transient hypertension in first trimester | Dyslipidemia, goal LDL below 160 | Atherosclerosis of autologous artery coronary artery bypass graft with angina pectoris with documented spasm |
| Antepartum transient hypertension in second trimester | Dyslipidemia, goal LDL below 55 | Atherosclerosis of autologous artery coronary artery bypass graft with other forms of angina pectoris |
| Antepartum transient hypertension in third trimester | Dyslipidemia, goal LDL below 70 | Atherosclerosis of autologous artery coronary artery bypass graft with refractory angina pectoris |
| Antepartum transient hypertension, first trimester | Elevated cholesterol | Atherosclerosis of autologous artery coronary artery bypass graft with stable angina |
| Antepartum transient hypertension, second trimester | Elevated cholesterol with elevated triglycerides | Atherosclerosis of autologous artery coronary artery bypass graft with unstable angina pectoris |
| Antepartum transient hypertension, third trimester | Elevated cholesterol with high triglycerides | Atherosclerosis of autologous artery coronary artery bypass graft(s) with angina pectoris with documented spasm |
| Antepartum transient hypertension, unspecified trimester | Elevated chylomicrons | Atherosclerosis of autologous artery coronary artery bypass graft(s) with other forms of angina pectoris |
| Asymptomatic hypertension | Elevated lipoprotein A level | Atherosclerosis of autologous artery coronary artery bypass graft(s) with refractory angina pectoris |
| Asymptomatic hypertensive urgency | Elevated lipoprotein(a) | Atherosclerosis of autologous artery coronary artery bypass graft(s) with unspecified angina pectoris |
| Asymptomatic systolic hypertension in pediatric patient | Elevated Lp(a) | Atherosclerosis of autologous artery coronary artery bypass graft(s) with unstable angina pectoris |
| Autosomal dominant pseudohypoaldosteronism type 1 with exacerbation of hypertension during pregnancy (CMS/HCC) | Elevated triglycerides with high cholesterol | Atherosclerosis of autologous coronary artery bypass graft |
| Ben hyp kid w cr kid V | Endogenous hyperglyceridemia | Atherosclerosis of autologous vein coronary artery bypass graft |
| Benign diastolic hypertension | Endogenous hyperlipemia | Atherosclerosis of autologous vein coronary artery bypass graft with angina pectoris |
| Benign essential HTN | Endogenous hyperlipidemia | Atherosclerosis of autologous vein coronary artery bypass graft with angina pectoris with documented spasm |
| Benign essential HTN, chronic, antepartum | Endogenous hypertriglyceridemia | Atherosclerosis of autologous vein coronary artery bypass graft with other forms of angina pectoris |
| Benign essential HTN, chronic, antepartum, first trimester | Essential familial hypercholesterolemia | Atherosclerosis of autologous vein coronary artery bypass graft with refractory angina pectoris |
| Benign essential HTN, chronic, antepartum, second trimester | Essential familial hyperlipidemia | Atherosclerosis of autologous vein coronary artery bypass graft with stable angina pectoris |
| Benign essential HTN, chronic, antepartum, third trimester | Essential hypertriglyceridemia | Atherosclerosis of autologous vein coronary artery bypass graft with unstable angina pectoris |
| Benign essential HTN, chronic, antepartum, unspecified trimester | Exogenous hyperlipidemia | Atherosclerosis of autologous vein coronary artery bypass graft, angina presence unspecified |
| Benign essential hypertension | Exogenous hypertriglyceridemia | Atherosclerosis of autologous vein coronary artery bypass graft, unspecified whether angina present |
| Benign essential hypertension affecting pregnancy | Familial apolipoprotein C-II deficiency | Atherosclerosis of autologous vein coronary artery bypass graft(s) with angina pectoris with documented spasm |
| Benign essential hypertension affecting pregnancy in first trimester | Familial apolipoprotein C-III deficiency | Atherosclerosis of autologous vein coronary artery bypass graft(s) with other forms of angina pectoris |
| Benign essential hypertension affecting pregnancy in second trimester | Familial chylomicronemia due to circulating inhibitor of lipoprotein lipase | Atherosclerosis of autologous vein coronary artery bypass graft(s) with refractory angina pectoris |
| Benign essential hypertension affecting pregnancy in third trimester | Familial chylomicronemia syndrome | Atherosclerosis of autologous vein coronary artery bypass graft(s) with unspecified angina pectoris |
| Benign essential hypertension affecting pregnancy, antepartum | Familial combined hyperlipidemia | Atherosclerosis of autologous vein coronary artery bypass graft(s) with unstable angina pectoris |
| Benign essential hypertension affecting pregnancy, childbirth, and puerperium | Familial combined hyperlipidemia associated with mutation in LPL gene | Atherosclerosis of bypass graft of coronary artery of transplanted heart with angina pectoris |
| Benign essential hypertension affecting pregnancy, childbirth, or puerperium, postpartum | Familial combined hyperlipidemia associated with mutation in USF1 gene | Atherosclerosis of bypass graft of coronary artery of transplanted heart with angina pectoris with documented spasm |
| Benign essential hypertension antepartum | Familial combined hyperlipidemia associated with p.Leu167del mutation in APOE gene | Atherosclerosis of bypass graft of coronary artery of transplanted heart with other forms of angina pectoris |
| Benign essential hypertension antepartum in first trimester | Familial combined hyperlipidemia type 2 | Atherosclerosis of bypass graft of coronary artery of transplanted heart with refractory angina pectoris |
| Benign essential hypertension antepartum in second trimester | Familial combined hyperlipidemia with APOE p.Leu167del variant | Atherosclerosis of bypass graft of coronary artery of transplanted heart with unspecified angina pectoris |
| Benign essential hypertension antepartum in third trimester | Familial defective apolipoprotein B-100 | Atherosclerosis of bypass graft of coronary artery of transplanted heart with unstable angina |
| Benign essential hypertension causing postpartum complication, delivered during previous episode of care | Familial dysbetalipoproteinemia | Atherosclerosis of bypass graft of coronary artery of transplanted heart without angina pectoris |
| Benign essential hypertension complicating childbirth | Familial fat-induced hypertriglyceridemia | Atherosclerosis of CABG w angina pectoris w documented spasm (CMS/HCC) |
| Benign essential hypertension complicating pregnancy | Familial hyperalphalipoproteinemia | Atherosclerosis of CABG w oth angina pectoris (CMS/HCC) |
| Benign essential hypertension complicating pregnancy, childbirth, and the puerperium | Familial hyperbetalipoproteinemia | Atherosclerosis of CABG w unsp angina pectoris (CMS/HCC) |
| Benign essential hypertension complicating pregnancy, childbirth, and the puerperium, unspecified as to episode of care | Familial hypercholesteremia | Atherosclerosis of CABG w unstable angina pectoris (CMS/HCC) |
| Benign essential hypertension complicating pregnancy, first trimester | Familial hypercholesterolemia | Atherosclerosis of CABG w/o angina pectoris |
| Benign essential hypertension complicating pregnancy, second trimester | Familial hypercholesterolemia due to biallelic mutation in LDLR gene | Atherosclerosis of CABG, unsp, w oth angina pectoris (CMS/HCC) |
| Benign essential hypertension complicating pregnancy, third trimester | Familial hypercholesterolemia due to combined heterozygous LDLR and heterozygous LDLRAP1 gene mutations | Atherosclerosis of CABG, unsp, w unsp angina pectoris (CMS/HCC) |
| Benign essential hypertension complicating pregnancy, unspecified trimester | Familial hypercholesterolemia due to genetic disorder of apolipoprotein b | Atherosclerosis of CABG, unsp, w unstable angina pectoris (CMS/HCC) |
| Benign essential hypertension complicating puerperium | Familial hypercholesterolemia due to heterozygous low density lipoprotein (LDL) receptor mutation | Atherosclerosis of coronary artery |
| Benign essential hypertension during pregnancy | Familial hypercholesterolemia due to homozygous low density lipoprotein (LDL) receptor mutation | Atherosclerosis of coronary artery bypass graft |
| Benign essential hypertension during pregnancy, antepartum | Familial hypercholesterolemia due to monoallelic mutation in LDLR gene | Atherosclerosis of coronary artery bypass graft of native heart |
| Benign essential hypertension in child | Familial hypercholesterolemia type 4 | Atherosclerosis of coronary artery bypass graft of native heart with angina pectoris |
| Benign essential hypertension in first trimester | Familial hypercholesterolemia, unspecified | Atherosclerosis of coronary artery bypass graft of native heart with angina pectoris with documented spasm |
| Benign essential hypertension in obstetric context | Familial hypercholesterolemia, unspecified type | Atherosclerosis of coronary artery bypass graft of native heart with other forms of angina pectoris |
| Benign essential hypertension in obstetric context in first trimester | Familial hyperchylomicronemia | Atherosclerosis of coronary artery bypass graft of native heart with refractory angina pectoris |
| Benign essential hypertension in obstetric context in second trimester | Familial hyperlipidemia | Atherosclerosis of coronary artery bypass graft of native heart with stable angina pectoris |
| Benign essential hypertension in obstetric context in third trimester | Familial hyperlipidemia, high LDL | Atherosclerosis of coronary artery bypass graft of native heart with unstable angina pectoris |
| Benign essential hypertension in obstetric context, antepartum | Familial hyperlipoproteinemia | Atherosclerosis of coronary artery bypass graft of native heart without angina pectoris |
| Benign essential hypertension in obstetric context, first trimester | Familial hyperlipoproteinemia type I | Atherosclerosis of coronary artery bypass graft of native heart, angina presence unspecified |
| Benign essential hypertension in obstetric context, second trimester | Familial hyperlipoproteinemia type II | Atherosclerosis of coronary artery bypass graft of native heart, unspecified whether angina present |
| Benign essential hypertension in obstetric context, third trimester | Familial hyperlipoproteinemia type IIa | Atherosclerosis of coronary artery bypass graft of transplanted heart |
| Benign essential hypertension in obstetric context, unspecified trimester | Familial hyperlipoproteinemia type IIb | Atherosclerosis of coronary artery bypass graft of transplanted heart with stable angina pectoris |
| Benign essential hypertension in pediatric patient | Familial hyperlipoproteinemia type III | Atherosclerosis of coronary artery bypass graft of transplanted heart, angina presence unspecified |
| Benign essential hypertension in second trimester | Familial hyperlipoproteinemia type IV | Atherosclerosis of coronary artery bypass graft of transplanted heart, unspecified whether angina present |
| Benign essential hypertension in third trimester | Familial hyperlipoproteinemia type V | Atherosclerosis of coronary artery bypass graft w/o angina pectoris |
| Benign essential hypertension with delivery | Familial hyperlipoproteinemia, type I | Atherosclerosis of coronary artery bypass graft with angina pectoris |
| Benign essential hypertension with target blood pressure below 140/90 | Familial hyperprebetalipoproteinemia | Atherosclerosis of coronary artery bypass graft with angina pectoris with documented spasm |
| Benign essential hypertension, age 0-18 | Familial hypertriglyceridemia | Atherosclerosis of coronary artery bypass graft with angina pectoris with documented spasm, unspecified whether native or transplanted heart |
| Benign essential hypertension, antepartum | Familial hypolipoproteinemia | Atherosclerosis of coronary artery bypass graft with angina pectoris, unspecified whether native or transplanted heart |
| Benign essential hypertension, delivered, with postpartum complication | Familial lipoprotein deficiency | Atherosclerosis of coronary artery bypass graft with other forms of angina pectoris |
| Benign essential hypertension, postpartum | Familial lipoprotein lipase deficiency | Atherosclerosis of coronary artery bypass graft with other forms of angina pectoris, unspecified whether native or transplanted heart |
| Benign essential hypertension, previous postpartum complication | Familial lipoprotein lipase deficiency with type 1 hyperlipoproteinemia phenotype | Atherosclerosis of coronary artery bypass graft with refractory angina pectoris |
| Benign essential hypertension, with delivery, with current postpartum complication | Familial lipoprotein lipase deficiency with type 5 hyperlipoproteinemia phenotype | Atherosclerosis of coronary artery bypass graft with refractory angina pectoris, unspecified whether native or transplanted heart |
| Benign hypertension | Familial LPL deficiency | Atherosclerosis of coronary artery bypass graft with stable angina pectoris |
| Benign hypertension affecting pregnancy, delivered, current hospitalization | Familial multiple lipoprotein-type hyperlipidemia | Atherosclerosis of coronary artery bypass graft with stable angina pectoris, unspecified whether native or transplanted heart |
| Benign hypertension complic pregnancy, with delivery, curr hospitaliz | Familial type 3 hyperlipoproteinemia | Atherosclerosis of coronary artery bypass graft with unstable angina pectoris |
| Benign hypertension complicating pregnancy, with delivery, current hospitalization | Familial type 5 hyperlipoproteinemia | Atherosclerosis of coronary artery bypass graft with unstable angina pectoris, unspecified whether native or transplanted heart |
| Benign hypertension due to increased intracranial pressure | Familial type III hyperlipoproteinemia | Atherosclerosis of coronary artery bypass graft without angina pectoris |
| Benign hypertension secondary to endocrine disorder | Familial type V hyperlipoproteinemia | Atherosclerosis of coronary artery bypass graft without angina pectoris, unspecified whether native or transplanted heart |
| Benign hypertension with chronic kidney disease | Fat induced hyperlipidemia | Atherosclerosis of coronary artery bypass graft, angina presence unspecified, unspecified whether native or transplanted heart |
| Benign hypertension with chronic kidney disease, stage I | FCHL (familial combined hyperlipidemia) | Atherosclerosis of coronary artery bypass graft, unspecified whether angina present, unspecified whether native or transplanted heart |
| Benign hypertension with chronic kidney disease, stage II | Floating beta disease | Atherosclerosis of coronary artery bypass graft(s) without angina pectoris |
| Benign hypertension with chronic kidney disease, stage III | Fredrickson type 2a hyperlipoproteinemia | Atherosclerosis of coronary artery bypass graft(s), unspecified, with angina pectoris with documented spasm |
| Benign hypertension with chronic kidney disease, stage IV | Fredrickson type 4 hyperlipoproteinemia | Atherosclerosis of coronary artery bypass graft(s), unspecified, with other forms of angina pectoris |
| Benign hypertension with chronic kidney disease, stage V | Fredrickson type 5 hyperlipoproteinemia | Atherosclerosis of coronary artery bypass graft(s), unspecified, with refractory angina pectoris |
| Benign hypertension with CKD (chronic kidney disease) stage I | Fredrickson type I hyperlipoproteinemia | Atherosclerosis of coronary artery bypass graft(s), unspecified, with unspecified angina pectoris |
| Benign hypertension with CKD (chronic kidney disease) stage III | Fredrickson type I or V hyperlipoproteinemia | Atherosclerosis of coronary artery bypass graft(s), unspecified, with unstable angina pectoris |
| Benign hypertension with CKD (chronic kidney disease) stage IV | Fredrickson type IIa hyperlipoproteinemia | Atherosclerosis of coronary artery of native heart |
| Benign hypertension with CKD (chronic kidney disease) stage V | Fredrickson type IIa hyperlipoproteinemia with target LDL less than 130 | Atherosclerosis of coronary artery of native heart with angina pectoris |
| Benign hypertension with CKD (chronic kidney disease), stage II | Fredrickson type IIa hyperlipoproteinemia with target LDL less than 160 | Atherosclerosis of coronary artery of native heart with angina pectoris and documented spasm |
| Benign hypertension with coincident congestive heart failure | Fredrickson type IIa hyperlipoproteinemia with target LDL less than 70 | Atherosclerosis of coronary artery of native heart with angina pectoris and documented spasm, unspecified vessel or lesion type |
| Benign hypertension with end-stage renal disease | Fredrickson type IIa hyperlipoproteinemia with target low density lipoprotein (LDL) cholesterol less than 130 mg/dL | Atherosclerosis of coronary artery of native heart with angina pectoris, unspecified vessel or lesion type |
| Benign hypertension with ESRD (end-stage renal disease) | Fredrickson type IIa hyperlipoproteinemia with target low density lipoprotein (LDL) cholesterol less than 160 mg/dL | Atherosclerosis of coronary artery of native heart with other form of angina pectoris |
| Benign hypertension with stage 3a chronic kidney disease | Fredrickson type IIa hyperlipoproteinemia with target low density lipoprotein (LDL) cholesterol less than 70 mg/dL | Atherosclerosis of coronary artery of native heart with other form of angina pectoris, unspecified vessel or lesion type |
| Benign hypertension with stage 3b chronic kidney disease | Fredrickson type IIb hyperlipoproteinemia | Atherosclerosis of coronary artery of native heart with stable angina pectoris |
| Benign hypertension without CHF | Fredrickson type IIb or III hyperlipoproteinemia | Atherosclerosis of coronary artery of native heart with stable angina pectoris, unspecified vessel or lesion type |
| Benign hypertension without congestive heart failure | Fredrickson type III hyperlipoproteinemia | Atherosclerosis of coronary artery of native heart with unstable angina pectoris |
| Benign hypertension, endocrine | Fredrickson type IV hyperlipoproteinemia | Atherosclerosis of coronary artery of native heart with unstable angina pectoris, unspecified vessel or lesion type |
| Benign hypertensive kidney disease with end stage renal disease | Fredrickson type IV lipidemia | Atherosclerosis of coronary artery of native heart without angina pectoris |
| Benign hypertensive kidney disease, stage V | Fredrickson type V hyperlipoproteinemia | Atherosclerosis of coronary artery of native heart without angina pectoris, unspecified vessel or lesion type |
| Benign labile hypertension | Genetically determined hyperlipidemia | Atherosclerosis of coronary artery of native heart, angina presence unspecified, unspecified vessel or lesion type |
| Benign reactive hypertension | Genetically determined mixed hyperlipidemia | Atherosclerosis of coronary artery of native heart, unspecified vessel or lesion type, unspecified whether angina present |
| Benign renal hypertension, non-vascular | Heterozygous familial hypercholesterolemia (hefh) | Atherosclerosis of coronary artery of transplanted heart |
| Benign renovascular hypertension | Heterozygous familial hypercholesterolemia | Atherosclerosis of coronary artery of transplanted heart with angina pectoris |
| Benign renovascular hypertension in child | Heterozygous hyperlipoproteinemia type 1 | Atherosclerosis of coronary artery of transplanted heart with angina pectoris and documented spasm |
| Benign renovascular hypertension in pediatric patient | Heterozygous phytosterolemia | Atherosclerosis of coronary artery of transplanted heart with angina pectoris and documented spasm, unspecified vessel or lesion type |
| Benign secondary hypertension | Heterozygous sitosterolemia | Atherosclerosis of coronary artery of transplanted heart with angina pectoris, unspecified vessel or lesion type |
| Benign secondary hypertension due to cyclosporine | High blood cholesterol | Atherosclerosis of coronary artery of transplanted heart with other form of angina pectoris |
| Benign secondary hypertension due to glomerulosclerosis | High blood cholesterol level | Atherosclerosis of coronary artery of transplanted heart with other form of angina pectoris, unspecified vessel or lesion type |
| Benign secondary hypertension due to glomerulosclerosis, stage 1-4 or unspecified chronic kidney disease | High blood triglycerides | Atherosclerosis of coronary artery of transplanted heart with stable angina pectoris |
| Benign secondary hypertension due to glomerulosclerosis, stage 5 chronic kidney disease or end stage renal disease | High cholesterol | Atherosclerosis of coronary artery of transplanted heart with stable angina pectoris, unspecified vessel or lesion type |
| Benign secondary hypertension due to pheochromocytoma | High triglycerides | Atherosclerosis of coronary artery of transplanted heart with unstable angina pectoris |
| Benign secondary hypertension due to polycystic kidney | HLD (hyperlipidemia) | Atherosclerosis of coronary artery of transplanted heart with unstable angina pectoris, unspecified vessel or lesion type |
| Benign secondary hypertension due to primary aldosteronism (CMS/HCC) | Homozygous familial hypercholesterolemia (hofh) | Atherosclerosis of coronary artery of transplanted heart without angina pectoris |
| Benign secondary hypertension due to renal artery stenosis | Homozygous familial hypercholesterolemia | Atherosclerosis of coronary artery of transplanted heart without angina pectoris, unspecified vessel or lesion type |
| Benign secondary hypertension due to renal disease | Homozygous sitosterolemia | Atherosclerosis of coronary artery of transplanted heart, angina presence unspecified, unspecified vessel or lesion type |
| Benign secondary hypertension in child | Hyperalphalipoproteinemia | Atherosclerosis of coronary artery of transplanted heart, unspecified vessel or lesion type, unspecified whether angina present |
| Benign secondary hypertension in pediatric patient | Hyperalphalipoproteinemia type 1 | Atherosclerosis of coronary artery with angina pectoris |
| Benign secondary hypertension related to oral contraceptive use | Hyperalphalipoproteinemia type 2 | Atherosclerosis of coronary artery with angina pectoris and documented spasm |
| Benign systolic hypertension | Hyperapobetalipoproteinemia | Atherosclerosis of coronary artery with angina pectoris and documented spasm, unspecified vessel or lesion type, unspecified whether native or transplanted heart |
| Bleeding in brain due to blood pressure disorder | Hyperbetalipoproteinemia | Atherosclerosis of coronary artery with angina pectoris, unspecified vessel or lesion type, unspecified whether native or transplanted heart |
| BP (high blood pressure) | Hyperbetalipoproteinemia with prebetalipoproteinemia | Atherosclerosis of coronary artery with other form of angina pectoris |
| Brachydactyly and arterial hypertension syndrome | Hypercholesteremia | Atherosclerosis of coronary artery with other form of angina pectoris, unspecified vessel or lesion type, unspecified whether native or transplanted heart |
| Brachydactyly with hypertension | Hypercholesterinemic xanthomatosis | Atherosclerosis of coronary artery with refractory angina pectoris |
| Brachydactyly-arterial hypertension syndrome | Hypercholesterolemia | Atherosclerosis of coronary artery with refractory angina pectoris, unspecified vessel or lesion type, unspecified whether native or transplanted heart |
| Chronic benign essential hypertension in first trimester | Hypercholesterolemia due to cholesterol 7alpha-hydroxylase deficiency | Atherosclerosis of coronary artery with stable angina pectoris |
| Chronic benign essential hypertension in second trimester | Hypercholesterolemia with endogenous hyperglyceridemia | Atherosclerosis of coronary artery with stable angina pectoris, unspecified vessel or lesion type, unspecified whether native or transplanted heart |
| Chronic benign essential hypertension in third trimester | Hypercholesterolemia with hyperglyceridemia | Atherosclerosis of coronary artery with unstable angina pectoris |
| Chronic benign essential hypertension, antepartum | Hypercholesterolemia with hypertriglyceridemia | Atherosclerosis of coronary artery with unstable angina pectoris, unspecified vessel or lesion type, unspecified whether native or transplanted heart |
| Chronic hypertension | Hypercholesterolemia with LDL greater than 190 mg/dL | Atherosclerosis of coronary artery without angina pectoris |
| Chronic hypertension affecting pregnancy | Hypercholesterolemia without hypertriglyceridemia | Atherosclerosis of coronary artery without angina pectoris, unspecified vessel or lesion type, unspecified whether native or transplanted heart |
| Chronic hypertension complicating or reason for care during childbirth | Hypercholesterolemic xanthomatosis | Atherosclerosis of coronary artery without history of bypass graft |
| Chronic hypertension complicating or reason for care during pregnancy | Hyperchylomicronaemia | Atherosclerosis of coronary artery, angina presence unspecified, unspecified vessel or lesion type, unspecified whether native or transplanted heart |
| Chronic hypertension complicating or reason for care during pregnancy, first trimester | Hyperchylomicronemia | Atherosclerosis of coronary artery, unspecified vessel or lesion type, unspecified whether angina present, unspecified whether native or transplanted heart |
| Chronic hypertension complicating or reason for care during pregnancy, second trimester | Hyperchylomicronemia, familial | Atherosclerosis of left anterior descending (LAD) artery |
| Chronic hypertension complicating or reason for care during pregnancy, third trimester | Hyperglyceridemia | Atherosclerosis of left circumflex artery |
| Chronic hypertension complicating or reason for care during pregnancy, unspecified trimester | Hyperglyceridemia, pure | Atherosclerosis of native coronary artery |
| Chronic hypertension during pregnancy | Hyperlipemia | Atherosclerosis of native coronary artery of native heart |
| Chronic hypertension during pregnancy, antepartum | Hyperlipemia, fat-induced | Atherosclerosis of native coronary artery of native heart with angina pectoris |
| Chronic hypertension in obstetric context | Hyperlipemia, idiopathic familial | Atherosclerosis of native coronary artery of native heart with angina pectoris with documented spasm |
| Chronic hypertension in obstetric context in first trimester | Hyperlipemia, mixed | Atherosclerosis of native coronary artery of native heart with other form of angina pectoris |
| Chronic hypertension in obstetric context in second trimester | Hyperlipemia, retention | Atherosclerosis of native coronary artery of native heart with refractory angina pectoris |
| Chronic hypertension in obstetric context in third trimester | Hyperlipidaemia with target low density lipoprotein (LDL) cholesterol less than 1.4 mmol/L | Atherosclerosis of native coronary artery of native heart with stable angina pectoris |
| Chronic hypertension in obstetric context, antepartum | Hyperlipidaemia with target low density lipoprotein (LDL) cholesterol less than 1.8 mmol/L | Atherosclerosis of native coronary artery of native heart with unstable angina pectoris |
| Chronic hypertension in obstetric context, first trimester | Hyperlipidaemia with target low density lipoprotein (LDL) cholesterol less than 2.0 mmol/L | Atherosclerosis of native coronary artery of native heart without angina pectoris |
| Chronic hypertension in obstetric context, second trimester | Hyperlipidaemia with target low density lipoprotein (LDL) cholesterol less than 2.6 mmol/L | Atherosclerosis of native coronary artery of native heart, angina presence unspecified |
| Chronic hypertension in obstetric context, third trimester | Hyperlipidaemia with target low density lipoprotein (LDL) cholesterol less than 3.0 mmol/L | Atherosclerosis of native coronary artery of native heart, unspecified whether angina present |
| Chronic hypertension in obstetric context, unspecified trimester | Hyperlipidaemia, unspecified | Atherosclerosis of native coronary artery of transplanted heart |
| Chronic hypertension in pregnancy | Hyperlipidemia | Atherosclerosis of native coronary artery of transplanted heart with angina pectoris |
| Chronic hypertension in pregnancy, first trimester | Hyperlipidemia associated with type 2 diabetes mellitus | Atherosclerosis of native coronary artery of transplanted heart with angina pectoris with documented spasm |
| Chronic hypertension in pregnancy, second trimester | Hyperlipidemia due to dietary fat intake | Atherosclerosis of native coronary artery of transplanted heart with other forms of angina pectoris |
| Chronic hypertension in pregnancy, third trimester | Hyperlipidemia due to type 1 diabetes mellitus | Atherosclerosis of native coronary artery of transplanted heart with refractory angina pectoris |
| Chronic hypertension in pregnancy, unspecified trimester | Hyperlipidemia due to type 2 diabetes mellitus | Atherosclerosis of native coronary artery of transplanted heart with stable angina pectoris |
| Chronic hypertension not affecting current episode of care | Hyperlipidemia LDL goal <100 | Atherosclerosis of native coronary artery of transplanted heart with unspecified angina pectoris |
| Chronic hypertension with exacerbation during pregnancy | Hyperlipidemia LDL goal <130 | Atherosclerosis of native coronary artery of transplanted heart with unstable angina |
| Chronic hypertension with exacerbation during pregnancy in first trimester | Hyperlipidemia LDL goal <160 | Atherosclerosis of native coronary artery of transplanted heart without angina pectoris |
| Chronic hypertension with exacerbation during pregnancy in first trimester, antepartum | Hyperlipidemia LDL goal <50 | Atherosclerosis of native coronary artery of transplanted heart, angina presence unspecified |
| Chronic hypertension with exacerbation during pregnancy in second trimester | Hyperlipidemia LDL goal <55 | Atherosclerosis of native coronary artery of transplanted heart, unspecified whether angina present |
| Chronic hypertension with exacerbation during pregnancy in second trimester, antepartum | Hyperlipidemia LDL goal <70 | Atherosclerosis of native coronary artery with angina pectoris |
| Chronic hypertension with exacerbation during pregnancy in third trimester | Hyperlipidemia type II | Atherosclerosis of native coronary artery with angina pectoris with documented spasm |
| Chronic hypertension with exacerbation during pregnancy in third trimester, antepartum | Hyperlipidemia type III | Atherosclerosis of native coronary artery with angina pectoris with documented spasm, unspecified whether native or transplanted heart |
| Chronic hypertension with exacerbation during pregnancy, antepartum | Hyperlipidemia type IV | Atherosclerosis of native coronary artery with angina pectoris, unspecified whether native or transplanted heart |
| Chronic hypertension with exacerbation during pregnancy, antepartum, first trimester | Hyperlipidemia with lipid deposition in skin | Atherosclerosis of native coronary artery with other form of angina pectoris |
| Chronic hypertension with exacerbation during pregnancy, antepartum, second trimester | Hyperlipidemia with low HDL | Atherosclerosis of native coronary artery with other form of angina pectoris, unspecified whether native or transplanted heart |
| Chronic hypertension with exacerbation during pregnancy, antepartum, third trimester | Hyperlipidemia with target LDL less than 100 | Atherosclerosis of native coronary artery with refractory angina pectoris |
| Chronic hypertension with exacerbation during pregnancy, antepartum, unspecified trimester | Hyperlipidemia with target LDL less than 130 | Atherosclerosis of native coronary artery with refractory angina pectoris, unspecified whether native or transplanted heart |
| Chronic hypertension with exacerbation during pregnancy, first trimester | Hyperlipidemia with target LDL less than 160 | Atherosclerosis of native coronary artery with stable angina pectoris |
| Chronic hypertension with exacerbation during pregnancy, second trimester | Hyperlipidemia with target LDL less than 70 | Atherosclerosis of native coronary artery with stable angina pectoris, unspecified whether native or transplanted heart |
| Chronic hypertension with exacerbation during pregnancy, third trimester | Hyperlipidemia with target low density lipoprotein (LDL) cholesterol less than 100 mg/dL | Atherosclerosis of native coronary artery with unstable angina pectoris |
| Chronic hypertension with exacerbation during pregnancy, unspecified trimester | Hyperlipidemia with target low density lipoprotein (LDL) cholesterol less than 130 mg/dL | Atherosclerosis of native coronary artery with unstable angina pectoris, unspecified whether native or transplanted heart |
| Chronic hypertension with superimposed pre-eclampsia | Hyperlipidemia with target low density lipoprotein (LDL) cholesterol less than 160 mg/dL | Atherosclerosis of native coronary artery without angina pectoris |
| Chronic hypertension with superimposed preeclampsia | Hyperlipidemia with target low density lipoprotein (LDL) cholesterol less than 50 mg/dL | Atherosclerosis of native coronary artery without angina pectoris, unspecified whether native or transplanted heart |
| Chronic kidney disease with malignant hypertension | Hyperlipidemia with target low density lipoprotein (LDL) cholesterol less than 55 mg/dL | Atherosclerosis of native coronary artery, angina presence unspecified, unspecified whether native or transplanted heart |
| Chronic kidney disease with malignant hypertension, stage 1-4 or unspecified chronic kidney disease | Hyperlipidemia with target low density lipoprotein (LDL) cholesterol less than 70 mg/dL | Atherosclerosis of native coronary artery, unspecified whether angina present, unspecified whether native or transplanted heart |
| Chronic kidney disease with malignant hypertension, stage 5 chronic kidney disease or end stage renal disease (CMS/HCC) | Hyperlipidemia, acquired | Atherosclerosis of native heart with refractory angina pectoris (CMS/HCC) |
| Chronic pre-existing hypertension during pregnancy | Hyperlipidemia, familial, high LDL | Atherosclerosis of native heart with refractory angina pectoris, unspecified vessel or lesion type (CMS/HCC) |
| Chronic spont intraparenchymal hemorrhage assoc w/ hypertension | Hyperlipidemia, group A | Atherosclerosis of non-autologous biological coronary artery bypass graft with stable angina pectoris |
| Chronic spont subarachnoid hemorrhage assoc w/ hypertension | Hyperlipidemia, group B | Atherosclerosis of non-autologous coronary artery bypass graft |
| Chronic spontaneous intraparenchymal intracranial hemorrhage associated with hypertension | Hyperlipidemia, group D | Atherosclerosis of nonautologous biological coronary artery bypass graft |
| Chronic spontaneous intraventricular hemorrhage assoc w/ hypertension | Hyperlipidemia, mild | Atherosclerosis of nonautologous biological coronary artery bypass graft with angina pectoris |
| Chronic spontaneous intraventricular hemorrhage associated with hypertension | Hyperlipidemia, mixed | Atherosclerosis of nonautologous biological coronary artery bypass graft with angina pectoris with documented spasm |
| Chronic spontaneous subarachnoid intracranial hemorrhage associated with hypertension | Hyperlipidemia, type IIb | Atherosclerosis of nonautologous biological coronary artery bypass graft with other forms of angina pectoris |
| Combined pre-existing hypertension and pre-eclampsia affecting childbirth | Hyperlipidemia, unspecified | Atherosclerosis of nonautologous biological coronary artery bypass graft with refractory angina pectoris |
| Combined pre-existing hypertension and pre-eclampsia affecting puerperium | Hyperlipidemia, unspecified hyperlipidemia | Atherosclerosis of nonautologous biological coronary artery bypass graft with unstable angina pectoris |
| Combined systolic and diastolic hypertension | Hyperlipidemia, unspecified hyperlipidemia type | Atherosclerosis of nonautologous biological coronary artery bypass graft, angina presence unspecified |
| Coronary sinus hypertension as complication of procedure | Hyperlipoproteinemia | Atherosclerosis of nonautologous biological coronary artery bypass graft, unspecified whether angina present |
| Cyclosporine-induced hypertension | Hyperlipoproteinemia type 1B | Atherosclerosis of nonautologous biological coronary artery bypass graft(s) with angina pectoris with documented spasm |
| Dia hypertension | Hyperlipoproteinemia type 1D | Atherosclerosis of nonautologous biological coronary artery bypass graft(s) with other forms of angina pectoris |
| Diabetes mellitus with coincident hypertension | Hyperlipoproteinemia type 2 | Atherosclerosis of nonautologous biological coronary artery bypass graft(s) with refractory angina pectoris |
| Diastolic hypertension | Hyperlipoproteinemia type 2 with hearing loss | Atherosclerosis of nonautologous biological coronary artery bypass graft(s) with unspecified angina pectoris |
| Eclampsia added to pre-existing hypertension | Hyperlipoproteinemia type I | Atherosclerosis of nonautologous biological coronary artery bypass graft(s) with unstable angina pectoris |
| Eclampsia superimposed on pre-existing hypertension, delivered | Hyperlipoproteinemia type II | Atherosclerosis of nonbiological coronary artery bypass graft |
| Eclampsia superimposed on pre-existing hypertension, delivered, with postpartum complication | Hyperlipoproteinemia type III | Atherosclerosis of other coronary artery bypass graft |
| Eclampsia w/ old hyp-del w/ postpartum complication | Hyperlipoproteinemia type IV | Atherosclerosis of other coronary artery bypass graft with angina pectoris |
| Eclampsia w/ old hyper-antepart | Hyperlipoproteinemia type V | Atherosclerosis of other coronary artery bypass graft with angina pectoris with documented spasm |
| Eclampsia w/ old hyper-postpart | Hyperlipoproteinemia, acquired | Atherosclerosis of other coronary artery bypass graft with other form of angina pectoris |
| Eclampsia w/ old hyperten-deliv | Hyperlipoproteinemia, Fredrickson type I | Atherosclerosis of other coronary artery bypass graft with refractory angina pectoris |
| Eclampsia w/ old hyperten-unsp | Hyperlipoproteinemia, Fredrickson type III | Atherosclerosis of other coronary artery bypass graft with stable angina pectoris |
| Eclampsia with pre-existing hypertension | Hyperlipoproteinemia, Fredrickson type IV | Atherosclerosis of other coronary artery bypass graft with unstable angina pectoris |
| Eclampsia with pre-existing hypertension during childbirth | Hyperlipoproteinemia, Fredrickson type V | Atherosclerosis of other coronary artery bypass graft without angina pectoris |
| Elevated blood pressure affecting pregnancy, antepartum | Hyperprebetalipoproteinemia | Atherosclerosis of other coronary artery bypass graft, angina presence unspecified |
| Elevated blood pressure complicating pregnancy, antepartum | Hyperprebetalipoproteinemia with chylomicronemia | Atherosclerosis of other coronary artery bypass graft, unspecified whether angina present |
| Elevated blood pressure reading in office with diagnosis of hypertension | Hypertriglyceridemia | Atherosclerosis of other coronary artery bypass graft(s) with angina pectoris with documented spasm |
| Elevated blood pressure reading in office with white coat syndrome, with diagnosis of hypertension | Hypertriglyceridemia associated with mutation in APOA5 gene | Atherosclerosis of other coronary artery bypass graft(s) with other forms of angina pectoris |
| Elevated blood pressure reading with diagnosis of hypertension | Hypertriglyceridemia associated with mutation in CREB3L3 gene | Atherosclerosis of other coronary artery bypass graft(s) with refractory angina pectoris |
| Elevated blood pressure, situational | Hypertriglyceridemia without hypercholesterolemia | Atherosclerosis of other coronary artery bypass graft(s) with unspecified angina pectoris |
| Elevated heart rate with elevated blood pressure and diagnosis of hypertension | Hypertriglyceridemia, essential | Atherosclerosis of other coronary artery bypass graft(s) with unstable angina pectoris |
| Elevated systolic blood pressure reading with diagnosis of hypertension | Hypertriglyceridemia, familial | Atherosclerosis of right coronary artery |
| Encounter for dietary treatment of hypertension | Hypertriglyceridemia, sporadic | Atherosclerosis of transplanted heart with refractory angina pectoris (CMS/HCC) |
| Encounter for follow-up for hypertension | Hypocholesteremia | Atherosclerosis of transplanted heart with refractory angina pectoris, unspecified vessel or lesion type (CMS/HCC) |
| Encounter for management of hypertension | Hypocholesterolemia | Atherosclerotic coronary vascular disease |
| End stage renal disease with hypertension (CMS/HCC) | Hypolipidemia | Atherosclerotic heart disease |
| Endocrinal hypertension | Hypomagnesemia, hypertension, and hypercholesterolemia associated with mutation in MT-TI gene | Atherosclerotic heart disease native coronary artery w/angina pectoris |
| Endocrine benign hypertension | Immunosuppressive-induced hyperlipidemia | Atherosclerotic heart disease of artery bypass graft |
| Endocrine hypertension | Isolated hypercholesterolemia | Atherosclerotic heart disease of autologous vein bypass graft |
| EPH (edema, proteinuria and hypertension of pregnancy) | Isolated hypertriglyceridemia | Atherosclerotic heart disease of coronary artery of transplanted heart |
| EPH (edema, proteinuria and hypertension of pregnancy), second trimester | Latent autoimmune diabetes mellitus in adult (LADA) with hyperlipidemia | Atherosclerotic heart disease of native coronary artery |
| EPH (edema, proteinuria and hypertension of pregnancy), third trimester | LDL (low density lipoprotein receptor disorder) | Atherosclerotic heart disease of native coronary artery with angina pectoris |
| EPH (edema, proteinuria and hypertension of pregnancy), unspecified trimester | LDL receptor disorder | Atherosclerotic heart disease of native coronary artery with angina pectoris with documented spasm |
| Essen hyperten preg-unsp | LDLR-related hypercholesterolemia, autosomal dominant | Atherosclerotic heart disease of native coronary artery with other form of angina pectoris |
| Essential (primary) hypertension | Lipidemia | Atherosclerotic heart disease of native coronary artery with other forms of angina pectoris |
| Essential hypertension | Lipoprotein lipase deficiency, familial | Atherosclerotic heart disease of native coronary artery with refractory angina pectoris |
| Essential hypertension affecting pregnancy | Low plasma apolipoprotein C-II | Atherosclerotic heart disease of native coronary artery with unspecified angina pectoris |
| Essential hypertension affecting pregnancy in first trimester | Low plasma apolipoprotein C2 | Atherosclerotic heart disease of native coronary artery with unstable angina pectoris |
| Essential hypertension affecting pregnancy in first trimester, antepartum | Low-density-lipoid-type (LDL) hyperlipoproteinemia | Atherosclerotic heart disease of native coronary artery without angina pectoris |
| Essential hypertension affecting pregnancy in second trimester | LPL-related familial combined hyperlipidemia | Atherosclerotic heart disease of nonautologous biological bypass graft |
| Essential hypertension affecting pregnancy in second trimester, antepartum | LRP1 gene mutation | Atherosclerotic heart disease of unspecified type of bypass graft |
| Essential hypertension affecting pregnancy in third trimester | Mild hypercholesterolemia | Atherosclerotic heart disease of unspecified type of vessel, native or graft |
| Essential hypertension affecting pregnancy in third trimester, antepartum | Mild hyperlipidemia | Atherosclerotic heart disease, of autologous bypass graft |
| Essential hypertension affecting pregnancy, antepartum | Mixed diabetic hyperlipidemia associated with type 1 diabetes mellitus | Atherosclerotic heart disease, of native coronary artery |
| Essential hypertension affecting pregnancy, antepartum, first trimester | Mixed diabetic hyperlipidemia associated with type 2 diabetes mellitus | Atherosclerotic heart disease, of nonautologous bypass graft |
| Essential hypertension affecting pregnancy, antepartum, second trimester | Mixed hypercholesterolemia and hypertriglyceridemia | Atherosclerotic heart disease, of unspecified vessel |
| Essential hypertension affecting pregnancy, antepartum, third trimester | Mixed hyperglyceridemia | Athscl autologous artery CABG w ang pctrs w documented spasm |
| Essential hypertension affecting pregnancy, antepartum, unspecified trimester | Mixed hyperlipidaemia | Athscl autologous artery CABG w oth angina pectoris |
| Essential hypertension antepartum | Mixed hyperlipidemia | Athscl autologous artery CABG w unsp angina pectoris |
| Essential hypertension antepartum, first trimester | Mixed hyperlipidemia due to type 1 diabetes mellitus | Athscl autologous artery CABG w unstable angina pectoris |
| Essential hypertension antepartum, second trimester | Mixed hyperlipidemia due to type 2 diabetes mellitus | Athscl autologous vein CABG w ang pctrs w documented spasm (CMS/HCC) |
| Essential hypertension antepartum, third trimester | Mixed hyperlipidemia with apolipoprotein E2 variant | Athscl autologous vein CABG w oth angina pectoris (CMS/HCC) |
| Essential hypertension antepartum, unspecified trimester | Mixed hyperlipidemia with apolipoprotein E3 variant | Athscl autologous vein CABG w unsp angina pectoris (CMS/HCC) |
| Essential hypertension associated with mutation in AGTR1 gene | Mixed hyperlipidemia with apolipoprotein E4 variant | Athscl autologous vein CABG w unstable angina pectoris (CMS/HCC) |
| Essential hypertension associated with mutation in PTGIS gene | Mixed hypertriglyceridemia | Athscl bypass of cor art of transplanted heart w/o ang pctrs |
| Essential hypertension complicating pregnancy | Moderate mixed hyperlipidemia not requiring statin therapy | Athscl bypass of cor art of txplt heart w ang pctrs w spasm (CMS/HCC) |
| Essential hypertension complicating pregnancy, first trimester | Multiple-type hyperlipidemia | Athscl bypass of cor art of txplt heart w oth ang pctrs (CMS/HCC) |
| Essential hypertension complicating pregnancy, second trimester | Mutation of LDLR gene | Athscl bypass of cor art of txplt heart w unsp ang pctrs (CMS/HCC) |
| Essential hypertension complicating pregnancy, third trimester | On treatment for hyperlipidemia | Athscl bypass of cor art of txplt heart w unstable angina (CMS/HCC) |
| Essential hypertension complicating pregnancy, unspecified trimester | Other and unspecified hyperlipidemia | Athscl CABG, unsp, w angina pectoris w documented spasm (CMS/HCC) |
| Essential hypertension during pregnancy | Other hyperlipidaemia | Athscl heart disease of native cor art w ang pctrs w spasm (CMS/HCC) |
| Essential hypertension during pregnancy in first trimester | Other hyperlipidemia | Athscl heart disease of native cor art w oth ang pctrs (CMS/HCC) |
| Essential hypertension during pregnancy in first trimester, antepartum | PCSK9-related hypercholesterolemia, autosomal dominant | Athscl heart disease of native cor art w unsp ang pctrs (CMS/HCC) |
| Essential hypertension during pregnancy in second trimester | Polygenic combined type IIb hyperlipidemia | Athscl heart disease of native cor art w unstable ang pctrs (CMS/HCC) |
| Essential hypertension during pregnancy in second trimester, antepartum | Polygenic hypercholesterolemia | Athscl heart disease of native coronary artery w/o ang pctrs |
| Essential hypertension during pregnancy in third trimester | Post-transplant hyperlipidemia | Athscl native cor art of transplanted heart w oth ang pctrs (CMS/HCC) |
| Essential hypertension during pregnancy in third trimester, antepartum | Predisposition to hypertriglyceridemia | Athscl native cor art of transplanted heart w unsp ang pctrs (CMS/HCC) |
| Essential hypertension during pregnancy, antepartum | Primary acquired chylomicronemia | Athscl native cor art of transplanted heart w/o ang pctrs |
| Essential hypertension during pregnancy, antepartum, first trimester | Primary chylomicronemia | Athscl native cor art of txplt heart w ang pctrs w spasm (CMS/HCC) |
| Essential hypertension during pregnancy, antepartum, second trimester | Primary combined hyperlipidemia | Athscl native cor art of txplt heart w unstable angina (CMS/HCC) |
| Essential hypertension during pregnancy, antepartum, third trimester | Primary dysbetalipoproteinemia | Athscl nonaut biological CABG w ang pctrs w documented spasm (CMS/HCC) |
| Essential hypertension during pregnancy, antepartum, unspecified trimester | Primary hypercholesterolemia | Athscl nonautologous biological CABG w oth angina pectoris (CMS/HCC) |
| Essential hypertension during pregnancy, delivered | Primary hyperchylomicronemia | Athscl nonautologous biological CABG w unsp angina pectoris (CMS/HCC) |
| Essential hypertension in child | Primary hypertriglyceridemia | Athscl nonautologous biological CABG w unstable ang pctrs (CMS/HCC) |
| Essential hypertension in first trimester | Pure hypercholesterolaemia | Autosomal dominant coronary artery disease associated with mutation in LRP6 gene |
| Essential hypertension in obstetric context | Pure hypercholesterolemia | Autosomal dominant coronary artery disease type 1 associated with mutation in MEF2A gene |
| Essential hypertension in obstetric context in first trimester | Pure hypercholesterolemia with target low density lipoprotein (LDL) cholesterol less than 130 mg/dL | Autosomal dominant coronary artery disease type 2 |
| Essential hypertension in obstetric context in second trimester | Pure hypercholesterolemia with target low density lipoprotein (LDL) cholesterol less than 160 mg/dL | Autosomal dominant coronary artery disease type 2 associated with mutation of LRP6 gene |
| Essential hypertension in obstetric context in third trimester | Pure hypercholesterolemia with target low density lipoprotein (LDL) cholesterol less than 70 mg/dL | Blockage of coronary artery bypass graft |
| Essential hypertension in obstetric context, antepartum | Pure hypercholesterolemia with target low density lipoprotein (LDL) less than 130 | Blockage of coronary artery bypass graft, initial encounter |
| Essential hypertension in obstetric context, first trimester | Pure hypercholesterolemia with target low density lipoprotein (LDL) less than 160 | Blockage of coronary artery bypass graft, sequela |
| Essential hypertension in obstetric context, second trimester | Pure hypercholesterolemia with target low density lipoprotein (LDL) less than 70 | Blockage of coronary artery bypass graft, subsequent encounter |
| Essential hypertension in obstetric context, third trimester | Pure hypercholesterolemia, unspecified | CAD (coronary artery disease) |
| Essential hypertension in obstetric context, unspecified trimester | Pure hyperglyceridaemia | CAD (CORONARY ARTERY DISEASE) OF ARTERY BYPASS GRAFT |
| Essential hypertension in pediatric patient | Pure hyperglyceridemia | CAD (coronary artery disease) of bypass graft |
| Essential hypertension in postpartum patient | Pure hypertriglyceridemia | CAD (coronary artery disease), autologous vein bypass graft |
| Essential hypertension in pregnancy | Remnant hyperlipidemia | CAD (coronary artery disease), bypass graft transplanted heart |
| Essential hypertension in pregnancy, delivered | Remnant hyperlipoproteinemia | CAD (coronary artery disease), native artery transplanted heart |
| Essential hypertension in third trimester, antepartum | Secondary combined hyperlipidemia | CAD (coronary artery disease), native coronary artery |
| Essential hypertension with goal blood pressure less than 130/80 | Secondary diabetes mellitus with hypercholesterolemia | CAD (coronary artery disease), nonautologous biological bypass graft |
| Essential hypertension with goal blood pressure less than 130/85 | Secondary diabetes mellitus with hyperlipidemia | CAD (coronary atherosclerotic disease) |
| Essential hypertension with goal blood pressure less than 140/90 | Secondary hypercholesterolemia | CAD (coronary atherosclerotic disease) without angina pectoris |
| Essential hypertension with goal blood pressure less than 150/90 | Secondary hyperlipidemia | CAD in native artery |
| Essential hypertension with high renin level | Secondary hypertriglyceridemia | CAD involving native coronary artery without angina pectoris |
| Essential hypertension-del with p/p | Sirolimus-induced hyperlipidemia | CAD of autologous arterial graft |
| Essential hypertension-postpartum | Sitosterolemia | CAD of autologous artery bypass graft without angina |
| Essential hypertension, benign | Sitosterolemia associated with mutation in ABCG5 gene | CAD of autologous bypass graft |
| Essential hypertension, malignant | Sitosterolemia associated with mutation in ABCG8 gene | CAD of autologous vein bypass graft without angina |
| Exertional hypertension | Sitosterolemia type 1 | CAD S/P percutaneous coronary angioplasty |
| Familial hypertensive hypokalemia disorder | Sitosterolemia type 2 | CAD without angina pectoris |
| Genetic essential hypertension | Sitosterolemia with xanthomatosis | CAD, multiple vessel |
| Gest h/t antepartum | Sitosterolemic xanthomatosis | Calcific coronary arteriosclerosis |
| Gest h/t deliv w compl | Steroid-induced hyperlipidemia | CHD (coronary heart disease) |
| Gest h/t delivered | Substance-induced hyperlipidemia | Chest pain due to CAD (CMS/HCC) |
| Gest h/t postpartum | Thrombophilia due to elevated lipoprotein A | Chest pain due to coronary artery disease |
| Gest h/t unspecified | Thrombophilia, elevated lipoprotein A, confirmed | Chronic coronary artery disease |
| Gestational (pregnancy-induced) hypertension | Transient infantile hypertriglyceridemia | Chronic diastolic heart failure secondary to coronary artery disease |
| Gestational (pregnancy-induced) hypertension without significant proteinuria | Transient infantile hypertriglyceridemia with hepatosteatosis | Clogged artery (heart) |
| Gestational (pregnancy-induced) hypertension without significant proteinuria, complicating childbirth | Type 1 diabetes mellitus with hypercholesterolemia | Congenital coronary artery sclerosis |
| Gestational (pregnancy-induced) hypertension without significant proteinuria, complicating the puerperium | Type 1 diabetes mellitus with hyperlipidemia | Cor athrscl-uns vessel |
| Gestational (pregnancy-induced) hypertension without significant proteinuria, first trimester | Type 1.5 diabetes with hyperlipidemia | Coronary arteriosclerosis |
| Gestational (pregnancy-induced) hypertension without significant proteinuria, second trimester | Type 2 diabetes mellitus with hypercholesterolemia | Coronary arteriosclerosis after coronary artery bypass grafting |
| Gestational (pregnancy-induced) hypertension without significant proteinuria, third trimester | Type 2 diabetes mellitus with hyperlipidemia | Coronary arteriosclerosis after percutaneous transluminal coronary angioplasty (PTCA) |
| Gestational (pregnancy-induced) hypertension without significant proteinuria, unspecified trimester | Type 2 diabetes mellitus with hypertriglyceridemia | Coronary arteriosclerosis due to lipid rich plaque |
| Gestational (pregnancy-induced) hypertension, antepartum condition or complication | Type I hyperlipoproteinemia | Coronary arteriosclerosis in native artery |
| Gestational (pregnancy-induced) hypertension, delivered, with mention of postpartum complication | Type II hyperlipoproteinemia | Coronary arteriosclerosis in patient with history of previous myocardial infarction |
| Gestational (pregnancy-induced) hypertension, delivered, with or without mention of antepartum condition | Type III hyperlipoproteinemia | Coronary arteriosclerosis of native coronary artery of transplanted heart |
| Gestational (pregnancy-induced) hypertension, postpartum condition or complication | Type IV hyperlipidemia | Coronary arteriosclerosis of transplanted heart |
| Gestational (pregnancy-induced) hypertension, unspecified as to episode of care, or not applicable | USF1-related familial combined hyperlipidemia | Coronary artery arteriosclerosis |
| Gestational HTN | Very low density lipoprotinemia | Coronary artery disease (CAD) excluded |
| Gestational htn w/o significant proteinuria, first trimester | Very-low-density-lipoid-type (VLDL) hyperlipoproteinemia | Coronary artery disease |
| Gestational htn w/o significant proteinuria, third trimester | Xanthoma due to primary chylomicronemia | Coronary artery disease due to calcified coronary lesion |
| Gestational htn w/o significant proteinuria, unsp trimester | Xanthoma due to primary combined hyperlipidemia | Coronary artery disease due to lipid rich plaque |
| Gestational HTN, first trimester | Xanthoma due to primary hypercholesterolemia | Coronary artery disease due to type 1 diabetes mellitus |
| Gestational HTN, second trimester | Xanthoma due to primary hypertriglyceridemia | Coronary artery disease due to type 2 diabetes mellitus |
| Gestational HTN, third trimester | Xanthoma due to secondary hyperlipidemia | Coronary artery disease excluded |
| Gestational HTN, unspecified trimester |  | Coronary artery disease in mother affecting pregnancy in first trimester, antepartum |
| Gestational hypertension |  | Coronary artery disease involving autologous artery coronary bypass graft |
| Gestational hypertension affecting eighth pregnancy |  | Coronary artery disease involving autologous artery coronary bypass graft with angina pectoris |
| Gestational hypertension affecting fifth pregnancy |  | Coronary artery disease involving autologous artery coronary bypass graft with angina pectoris with documented spasm |
| Gestational hypertension affecting first pregnancy |  | Coronary artery disease involving autologous artery coronary bypass graft with other forms of angina pectoris |
| Gestational hypertension affecting fourth pregnancy |  | Coronary artery disease involving autologous artery coronary bypass graft with refractory angina pectoris |
| Gestational hypertension affecting ninth pregnancy |  | Coronary artery disease involving autologous artery coronary bypass graft with unstable angina pectoris |
| Gestational hypertension affecting second pregnancy |  | Coronary artery disease involving autologous artery coronary bypass graft without angina pectoris |
| Gestational hypertension affecting seventh pregnancy |  | Coronary artery disease involving autologous artery coronary bypass graft, angina presence unspecified |
| Gestational hypertension affecting sixth pregnancy |  | Coronary artery disease involving autologous artery coronary bypass graft, unspecified whether angina present |
| Gestational hypertension affecting tenth pregnancy |  | Coronary artery disease involving autologous vein bypass graft |
| Gestational hypertension affecting third pregnancy |  | Coronary artery disease involving autologous vein bypass graft, angina presence unspecified |
| Gestational hypertension associated with abortive pregnancy |  | Coronary artery disease involving autologous vein bypass graft, unspecified whether angina present |
| Gestational hypertension w/o significant proteinuria in 2nd trimester |  | Coronary artery disease involving autologous vein coronary bypass graft with angina pectoris |
| Gestational hypertension w/o significant proteinuria in 3rd trimester |  | Coronary artery disease involving autologous vein coronary bypass graft with angina pectoris with documented spasm |
| Gestational hypertension with significant proteinuria, delivered |  | Coronary artery disease involving autologous vein coronary bypass graft with other forms of angina pectoris |
| Gestational hypertension with significant proteinuria, postpartum |  | Coronary artery disease involving autologous vein coronary bypass graft with refractory angina pectoris |
| Gestational hypertension without significant proteinuria |  | Coronary artery disease involving autologous vein coronary bypass graft with unstable angina pectoris |
| Gestational hypertension without significant proteinuria during pregnancy in first trimester, antepartum |  | Coronary artery disease involving autologous vein coronary bypass graft without angina pectoris |
| Gestational hypertension without significant proteinuria during pregnancy in second trimester, antepartum |  | Coronary artery disease involving bypass graft of transplanted heart |
| Gestational hypertension without significant proteinuria during pregnancy in third trimester, antepartum |  | Coronary artery disease involving bypass graft of transplanted heart with angina pectoris |
| Gestational hypertension without significant proteinuria in first trimester |  | Coronary artery disease involving bypass graft of transplanted heart with angina pectoris with documented spasm |
| Gestational hypertension without significant proteinuria in second trimester |  | Coronary artery disease involving bypass graft of transplanted heart with other forms of angina pectoris |
| Gestational hypertension without significant proteinuria in third trimester |  | Coronary artery disease involving bypass graft of transplanted heart with unstable angina pectoris |
| Gestational hypertension without significant proteinuria, affecting childbirth |  | Coronary artery disease involving bypass graft of transplanted heart without angina pectoris |
| Gestational hypertension without significant proteinuria, affecting puerperium |  | Coronary artery disease involving bypass graft of transplanted heart, angina presence unspecified |
| Gestational hypertension without significant proteinuria, antepartum |  | Coronary artery disease involving bypass graft of transplanted heart, unspecified whether angina present |
| Gestational hypertension without significant proteinuria, antepartum, first trimester |  | Coronary artery disease involving coronary bypass graft |
| Gestational hypertension without significant proteinuria, antepartum, second trimester |  | Coronary artery disease involving coronary bypass graft of native heart |
| Gestational hypertension without significant proteinuria, antepartum, third trimester |  | Coronary artery disease involving coronary bypass graft of native heart with angina pectoris |
| Gestational hypertension without significant proteinuria, antepartum, unspecified trimester |  | Coronary artery disease involving coronary bypass graft of native heart with angina pectoris with documented spasm |
| Gestational hypertension without significant proteinuria, first trimester |  | Coronary artery disease involving coronary bypass graft of native heart with other forms of angina pectoris |
| Gestational hypertension without significant proteinuria, postpartum |  | Coronary artery disease involving coronary bypass graft of native heart with refractory angina pectoris |
| Gestational hypertension without significant proteinuria, second trimester |  | Coronary artery disease involving coronary bypass graft of native heart with unstable angina pectoris |
| Gestational hypertension without significant proteinuria, third trimester |  | Coronary artery disease involving coronary bypass graft of native heart without angina pectoris |
| Gestational hypertension without significant proteinuria, unspecified trimester |  | Coronary artery disease involving coronary bypass graft of native heart, angina presence unspecified |
| Gestational hypertension, antepartum |  | Coronary artery disease involving coronary bypass graft of native heart, unspecified whether angina present |
| Gestational hypertension, first trimester |  | Coronary artery disease involving coronary bypass graft of transplanted heart with refractory angina pectoris |
| Gestational hypertension, second trimester |  | Coronary artery disease involving coronary bypass graft with angina pectoris |
| Gestational hypertension, third trimester |  | Coronary artery disease involving coronary bypass graft with angina pectoris with documented spasm |
| Gestational hypertension, unspecified trimester |  | Coronary artery disease involving coronary bypass graft with angina pectoris with documented spasm, unspecified whether native or transplanted heart |
| Gestatnl htn w/o significant proteinuria, second trimester |  | Coronary artery disease involving coronary bypass graft with angina pectoris, unspecified whether native or transplanted heart |
| Gestosis, second trimester |  | Coronary artery disease involving coronary bypass graft with other forms of angina pectoris |
| Gestosis, third trimester |  | Coronary artery disease involving coronary bypass graft with other forms of angina pectoris, unspecified whether native or transplanted heart |
| Gestosis, unspecified trimester |  | Coronary artery disease involving coronary bypass graft with refractory angina pectoris |
| Goldblatt hypertension |  | Coronary artery disease involving coronary bypass graft with refractory angina pectoris, unspecified whether native or transplanted heart |
| Goldblatt's, hypertension |  | Coronary artery disease involving coronary bypass graft with unstable angina pectoris |
| HBP (high blood pressure) |  | Coronary artery disease involving coronary bypass graft with unstable angina pectoris, unspecified whether native or transplanted heart |
| HELLP (hemolytic anemia/elev liver enzymes/low platelets in pregnancy) |  | Coronary artery disease involving coronary bypass graft without angina pectoris |
| HELLP (hemolytic anemia/elev liver enzymes/low platelets in pregnancy), antepartum |  | Coronary artery disease involving coronary bypass graft without angina pectoris, unspecified whether native or transplanted heart |
| HELLP (hemolytic anemia/elev liver enzymes/low platelets in pregnancy), second trimester |  | Coronary artery disease involving coronary bypass graft, angina presence unspecified, unspecified whether native or transplanted heart |
| HELLP (hemolytic anemia/elev liver enzymes/low platelets in pregnancy), third trimester |  | Coronary artery disease involving coronary bypass graft, unspecified whether angina present, unspecified whether native or transplanted heart |
| HELLP (hemolytic anemia/elev liver enzymes/low platelets in pregnancy), unspecified trimester |  | Coronary artery disease involving left main coronary artery |
| HELLP syndrome (HELLP), second trimester |  | Coronary artery disease involving native artery of transplanted heart |
| HELLP syndrome (HELLP), third trimester |  | Coronary artery disease involving native artery of transplanted heart with angina pectoris |
| HELLP syndrome (HELLP), unspecified trimester |  | Coronary artery disease involving native artery of transplanted heart with angina pectoris with documented spasm |
| HELLP syndrome |  | Coronary artery disease involving native artery of transplanted heart with other forms of angina pectoris |
| HELLP syndrome delivered |  | Coronary artery disease involving native artery of transplanted heart with unstable angina pectoris |
| HELLP syndrome in first trimester |  | Coronary artery disease involving native artery of transplanted heart without angina pectoris |
| HELLP syndrome in second trimester |  | Coronary artery disease involving native artery of transplanted heart, angina presence unspecified |
| HELLP syndrome in third trimester |  | Coronary artery disease involving native artery of transplanted heart, unspecified whether angina present |
| HELLP syndrome, antepartum |  | Coronary artery disease involving native coronary artery |
| HELLP syndrome, antepartum condition or complication |  | Coronary artery disease involving native coronary artery of native heart |
| HELLP syndrome, complicating childbirth |  | Coronary artery disease involving native coronary artery of native heart with angina pectoris |
| HELLP syndrome, complicating the puerperium |  | Coronary artery disease involving native coronary artery of native heart with angina pectoris with documented spasm |
| HELLP syndrome, delivered, current hospitalization |  | Coronary artery disease involving native coronary artery of native heart with other form of angina pectoris |
| HELLP syndrome, delivered, with mention of postpartum complication |  | Coronary artery disease involving native coronary artery of native heart with refractory angina pectoris |
| HELLP syndrome, delivered, with or without mention of antepartum condition |  | Coronary artery disease involving native coronary artery of native heart with unstable angina pectoris |
| HELLP syndrome, postpartum condition or complication |  | Coronary artery disease involving native coronary artery of native heart without angina pectoris |
| HELLP syndrome, second trimester |  | Coronary artery disease involving native coronary artery of native heart, angina presence unspecified |
| HELLP syndrome, third trimester |  | Coronary artery disease involving native coronary artery of native heart, unspecified whether angina present |
| HELLP syndrome, unspecified as to episode of care, or not applicable |  | Coronary artery disease involving native coronary artery of transplanted heart with refractory angina pectoris |
| HELLP syndrome, unspecified trimester |  | Coronary artery disease involving native coronary artery with angina pectoris |
| Hemolysis, elevated liver enzymes, and low platelet (HELLP) syndrome |  | Coronary artery disease involving native coronary artery with angina pectoris with documented spasm |
| Hemolysis, elevated liver enzymes, and low platelet (HELLP) syndrome druing pregnancy, delivered |  | Coronary artery disease involving native coronary artery with angina pectoris with documented spasm, unspecified whether native or transplanted heart |
| Hemolysis, elevated liver enzymes, and low platelet (HELLP) syndrome during pregnancy |  | Coronary artery disease involving native coronary artery with angina pectoris, unspecified whether native or transplanted heart |
| Hemolysis, elevated liver enzymes, and low platelet (HELLP) syndrome during pregnancy in third trimester |  | Coronary artery disease involving native coronary artery with other form of angina pectoris, unspecified whether native or transplanted heart |
| Hemolysis, elevated liver enzymes, and low platelet (HELLP) syndrome during pregnancy, antepartum |  | Coronary artery disease involving native coronary artery with other forms of angina pectoris |
| Hemolysis, elevated liver enzymes, and low platelet (HELLP) syndrome during pregnancy, delivered |  | Coronary artery disease involving native coronary artery with other forms of angina pectoris, unspecified whether native or transplanted heart |
| Hemolysis, elevated liver enzymes, and low platelet (HELLP) syndrome during pregnancy, delivered, with postpartum complication |  | Coronary artery disease involving native coronary artery with refractory angina pectoris |
| Hemolysis, elevated liver enzymes, and low platelet (HELLP) syndrome during pregnancy, postpartum |  | Coronary artery disease involving native coronary artery with refractory angina pectoris, unspecified whether native or transplanted heart |
| Hemolysis, elevated liver enzymes, and low platelet (HELLP) syndrome during pregnancy, second trimester |  | Coronary artery disease involving native coronary artery with unstable angina pectoris |
| Hemolysis, elevated liver enzymes, and low platelet (HELLP) syndrome during pregnancy, third trimester |  | Coronary artery disease involving native coronary artery with unstable angina pectoris, unspecified whether native or transplanted heart |
| Hemolysis, elevated liver enzymes, and low platelet (HELLP) syndrome during pregnancy, unspecified trimester |  | Coronary artery disease involving native coronary artery without angina pectoris |
| Hemolysis, elevated liver enzymes, and low platelet (HELLP) syndrome in pregnancy |  | Coronary artery disease involving native coronary artery without angina pectoris, unspecified whether native or transplanted heart |
| Hemolysis, elevated liver enzymes, and low platelet (HELLP) syndrome in pregnancy in third trimester |  | Coronary artery disease involving native coronary artery, angina presence unspecified, unspecified whether native or transplanted heart |
| Hemolysis, elevated liver enzymes, and low platelet (HELLP) syndrome in pregnancy, second trimester |  | Coronary artery disease involving native coronary artery, unspecified whether angina present, unspecified whether native or transplanted heart |
| Hemolysis, elevated liver enzymes, and low platelet (HELLP) syndrome in pregnancy, third trimester |  | Coronary artery disease involving native heart |
| Hemolysis, elevated liver enzymes, and low platelet (HELLP) syndrome in pregnancy, unspecified trimester |  | Coronary artery disease involving native heart with angina pectoris |
| Hemolysis, elevated liver enzymes, and low platelet (HELLP) syndrome in third trimester |  | Coronary artery disease involving native heart with angina pectoris and documented spasm |
| Hemolysis, elevated liver enzymes, and low platelet (HELLP) syndrome, antepartum |  | Coronary artery disease involving native heart with angina pectoris and documented spasm, unspecified vessel or lesion type |
| Hemolysis, elevated liver enzymes, and low platelet (HELLP) syndrome, postpartum |  | Coronary artery disease involving native heart with angina pectoris, unspecified vessel or lesion type |
| Hemolysis, elevated liver enzymes, and low platelet count (HELLP) syndrome in first trimester |  | Coronary artery disease involving native heart with other form of angina pectoris |
| Hemolysis, elevated liver enzymes, and low platelet count (HELLP) syndrome in second trimester |  | Coronary artery disease involving native heart with other form of angina pectoris, unspecified vessel or lesion type |
| Hemolysis, elevated liver enzymes, and low platelet count (HELLP) syndrome, delivered, current hospitalization |  | Coronary artery disease involving native heart with refractory angina pectoris |
| Hemolysis, elevated liver enzymes, and low platelets (HELLP) syndrome affecting childbirth |  | Coronary artery disease involving native heart with refractory angina pectoris, unspecified vessel or lesion type |
| Hemolysis, elevated liver enzymes, and low platelets (HELLP) syndrome affecting puerperium |  | Coronary artery disease involving native heart with unstable angina pectoris |
| Hemolysis, elevated liver enzymes, and low platelets (HELLP) syndrome with abortive outcome |  | Coronary artery disease involving native heart with unstable angina pectoris, unspecified vessel or lesion type |
| Hemolytic anemia, elevated liver enzymes, low platelets in pregnancy |  | Coronary artery disease involving native heart without angina pectoris |
| Hemolytic anemia, elevated liver enzymes, low platelets in pregnancy, second trimester |  | Coronary artery disease involving native heart without angina pectoris, unspecified vessel or lesion type |
| Hemolytic anemia, elevated liver enzymes, low platelets in pregnancy, third trimester |  | Coronary artery disease involving native heart, angina presence unspecified, unspecified vessel or lesion type |
| Hemolytic anemia, elevated liver enzymes, low platelets in pregnancy, unspecified trimester |  | Coronary artery disease involving native heart, unspecified vessel or lesion type, unspecified whether angina present |
| High blood pressure |  | Coronary artery disease involving nonautologous biological coronary bypass graft |
| High blood pressure affecting pregnancy, antepartum |  | Coronary artery disease involving nonautologous biological coronary bypass graft with angina pectoris |
| High blood pressure associated with diabetes |  | Coronary artery disease involving nonautologous biological coronary bypass graft with angina pectoris with documented spasm |
| High blood pressure determined by examination |  | Coronary artery disease involving nonautologous biological coronary bypass graft with other forms of angina pectoris |
| High blood pressure disorder |  | Coronary artery disease involving nonautologous biological coronary bypass graft with refractory angina pectoris |
| High blood pressure due to drug |  | Coronary artery disease involving nonautologous biological coronary bypass graft with unstable angina pectoris |
| High blood pressure due to drug, accidental or unintentional, initial encounter |  | Coronary artery disease involving nonautologous biological coronary bypass graft without angina pectoris |
| High blood pressure due to drug, accidental or unintentional, sequela |  | Coronary artery disease involving nonautologous biological coronary bypass graft, angina presence unspecified |
| High blood pressure due to drug, accidental or unintentional, subsequent encounter |  | Coronary artery disease involving nonautologous biological coronary bypass graft, unspecified whether angina present |
| High blood pressure due to drug, assault, initial encounter |  | Coronary artery disease involving other coronary artery bypass graft |
| High blood pressure due to drug, assault, sequela |  | Coronary artery disease involving other coronary artery bypass graft with angina pectoris |
| High blood pressure due to drug, assault, subsequent encounter |  | Coronary artery disease involving other coronary artery bypass graft with angina pectoris with documented spasm |
| High blood pressure due to drug, intentional self-harm, initial encounter |  | Coronary artery disease involving other coronary artery bypass graft with other forms of angina pectoris |
| High blood pressure due to drug, intentional self-harm, sequela |  | Coronary artery disease involving other coronary artery bypass graft with refractory angina pectoris |
| High blood pressure due to drug, intentional self-harm, subsequent encounter |  | Coronary artery disease involving other coronary artery bypass graft with unspecified angina pectoris |
| High blood pressure due to drug, undetermined intent, initial encounter |  | Coronary artery disease involving other coronary artery bypass graft with unstable angina pectoris |
| High blood pressure due to drug, undetermined intent, sequela |  | Coronary artery disease involving other coronary artery bypass graft without angina pectoris |
| High blood pressure due to drug, undetermined intent, subsequent encounter |  | Coronary artery disease involving other coronary artery bypass graft, unspecified whether angina present |
| High blood pressure due to oral contraceptive |  | Coronary artery disease involving transplanted heart |
| High blood pressure due to overproduction of aldosterone (CMS/HCC) |  | Coronary artery disease involving transplanted heart with angina pectoris |
| High blood pressure with chronic kidney disease |  | Coronary artery disease involving transplanted heart with angina pectoris and documented spasm |
| High blood pressure with chronic kidney disease, stage 1 through stage 4 or unspecified chronic kidney disease |  | Coronary artery disease involving transplanted heart with angina pectoris and documented spasm, unspecified vessel or lesion type |
| High blood pressure with chronic kidney disease, stage 1-4 or unspecified chronic kidney disease |  | Coronary artery disease involving transplanted heart with angina pectoris, unspecified vessel or lesion type |
| High blood pressure with chronic kidney disease, stage 5 chronic kidney disease or end stage renal disease |  | Coronary artery disease involving transplanted heart with other form of angina pectoris |
| High-renin essential hypertension |  | Coronary artery disease involving transplanted heart with other form of angina pectoris, unspecified vessel or lesion type |
| Hormonal hypertension |  | Coronary artery disease involving transplanted heart with refractory angina pectoris |
| HTN (hypertension) |  | Coronary artery disease involving transplanted heart with refractory angina pectoris, unspecified vessel or lesion type |
| HTN (hypertension) with goal to be determined |  | Coronary artery disease involving transplanted heart with unstable angina pectoris |
| HTN (hypertension), benign |  | Coronary artery disease involving transplanted heart with unstable angina pectoris, unspecified vessel or lesion type |
| HTN (hypertension), malignant |  | Coronary artery disease involving transplanted heart without angina pectoris |
| HTN complicating peripregnancy, antepartum |  | Coronary artery disease involving transplanted heart without angina pectoris, unspecified vessel or lesion type |
| HTN complicating peripregnancy, antepartum, first trimester |  | Coronary artery disease involving transplanted heart, angina presence unspecified, unspecified vessel or lesion type |
| HTN complicating peripregnancy, antepartum, second trimester |  | Coronary artery disease involving transplanted heart, unspecified vessel or lesion type, unspecified whether angina present |
| HTN complicating peripregnancy, antepartum, third trimester |  | Coronary artery disease of autologous bypass graft with stable angina pectoris |
| HTN complicating peripregnancy, antepartum, unspecified trimester |  | Coronary artery disease of autologous vein bypass graft with stable angina pectoris |
| HTN in pregnancy, chronic |  | Coronary artery disease of bypass graft of native heart with stable angina pectoris |
| HTN in pregnancy, chronic, first trimester |  | Coronary artery disease of bypass graft of transplanted heart with stable angina pectoris |
| HTN in pregnancy, chronic, second trimester |  | Coronary artery disease of bypass graft with stable angina pectoris |
| HTN in pregnancy, chronic, third trimester |  | Coronary artery disease of bypass graft with stable angina pectoris, unspecified whether native or transplanted heart |
| HTN in pregnancy, chronic, unspecified trimester |  | Coronary artery disease of native artery of native heart with stable angina pectoris |
| HTN, age 0-18 |  | Coronary artery disease of native artery of transplanted heart with stable angina pectoris |
| HTN, goal below 130/80 |  | Coronary artery disease of native artery with stable angina pectoris |
| HTN, goal below 140/80 |  | Coronary artery disease of native artery with stable angina pectoris, unspecified whether native or transplanted heart |
| HTN, goal below 140/90 |  | Coronary artery disease of native heart with stable angina pectoris |
| HTN, goal below 150/90 |  | Coronary artery disease of native heart with stable angina pectoris, unspecified vessel or lesion type |
| HTN, goal to be determined |  | Coronary artery disease of non-autologous biological bypass graft with stable angina pectoris |
| HTN, goal: symptom mgmt only |  | Coronary artery disease of other bypass graft with stable angina pectoris |
| HTN, kidney transplant related |  | Coronary artery disease of transplanted heart |
| HTN, white coat |  | Coronary artery disease of transplanted heart with stable angina pectoris |
| HVD (hypertensive vascular disease) |  | Coronary artery disease of transplanted heart with stable angina pectoris, unspecified vessel or lesion type |
| Hypergency in pregnancy, benign essential |  | Coronary artery disease status post coronary stent insertion |
| Hypergency in pregnancy, essential |  | Coronary artery disease with angina pectoris |
| Hyperpiesia |  | Coronary artery disease with angina pectoris with documented spasm |
| Hyperpiesis |  | Coronary artery disease with angina pectoris with documented spasm, unspecified vessel or lesion type, unspecified whether native or transplanted heart |
| Hyperten preg NOS-unspec |  | Coronary artery disease with angina pectoris, unspecified vessel or lesion type, unspecified whether native or transplanted heart |
| Hypertens NOS-del w/ postpartum complication |  | Coronary artery disease with cardiac symptoms |
| Hypertension |  | Coronary artery disease with cardiac symptoms and history of coronary revascularization |
| Hypertension affecting pregnancy |  | Coronary artery disease with exertional angina |
| Hypertension affecting pregnancy in first trimester |  | Coronary artery disease with history of coronary revascularization |
| Hypertension affecting pregnancy in first trimester, antepartum |  | Coronary artery disease with history of incomplete coronary revascularization |
| Hypertension affecting pregnancy in second trimester |  | Coronary artery disease with history of myocardial infarction without history of CABG |
| Hypertension affecting pregnancy in second trimester, antepartum |  | Coronary artery disease with history of percutaneous transluminal angioplasty (PTCA) |
| Hypertension affecting pregnancy in third trimester |  | Coronary artery disease with hx of myocardial infarct w/o hx of CABG |
| Hypertension affecting pregnancy in third trimester, antepartum |  | Coronary artery disease with other form of angina pectoris |
| Hypertension affecting pregnancy, antepartum |  | Coronary artery disease with other form of angina pectoris, unspecified vessel or lesion type, unspecified whether native or transplanted heart |
| Hypertension affecting pregnancy, antepartum, first trimester |  | Coronary artery disease with recent acute coronary syndrome |
| Hypertension affecting pregnancy, antepartum, second trimester |  | Coronary artery disease with recent acute coronary syndrome and history of coronary revascularization |
| Hypertension affecting pregnancy, antepartum, third trimester |  | Coronary artery disease with recent acute coronary syndrome and incomplete coronary revascularization |
| Hypertension affecting pregnancy, antepartum, unspecified trimester |  | Coronary artery disease with refractory angina pectoris |
| Hypertension affecting pregnancy, delivered, current hospitalization |  | Coronary artery disease with refractory angina pectoris, unspecified vessel or lesion type, unspecified whether native or transplanted heart |
| Hypertension affecting pregnancy, first trimester |  | Coronary artery disease with stable angina pectoris |
| Hypertension affecting pregnancy, second trimester |  | Coronary artery disease with stable angina pectoris, unspecified vessel or lesion type, unspecified whether native or transplanted heart |
| Hypertension affecting pregnancy, third trimester |  | Coronary artery disease with unstable angina pectoris |
| Hypertension affecting pregnancy, unspecified trimester |  | Coronary artery disease with unstable angina pectoris, unspecified vessel or lesion type, unspecified whether native or transplanted heart |
| Hypertension after donor nephrectomy requiring medication |  | Coronary artery disease without angina pectoris |
| Hypertension as manifestation of blood transfusion reaction |  | Coronary artery disease without angina pectoris, unspecified vessel or lesion type, unspecified whether native or transplanted heart |
| Hypertension as manifestation of blood transfusion reaction, initial encounter |  | Coronary artery disease, angina presence unspecified, unspecified vessel or lesion type, unspecified whether native or transplanted heart |
| Hypertension as manifestation of blood transfusion reaction, sequela |  | Coronary artery disease, non-occlusive |
| Hypertension as manifestation of blood transfusion reaction, subsequent encounter |  | Coronary artery disease, occlusive |
| Hypertension associated with chronic kidney disease due to type 1 diabetes mellitus |  | Coronary artery disease, unspecified vessel or lesion type, unspecified whether angina present, unspecified whether native or transplanted heart |
| Hypertension associated with chronic kidney disease due to type 2 diabetes mellitus |  | Coronary artery sclerosis |
| Hypertension associated with diabetes |  | Coronary atherosclerosis |
| Hypertension associated with stage 2 chronic kidney disease due to type 1 diabetes mellitus |  | Coronary atherosclerosis due to calcified coronary lesion (CODE) |
| Hypertension associated with stage 2 chronic kidney disease due to type 2 diabetes mellitus |  | Coronary atherosclerosis due to calcified coronary lesion |
| Hypertension associated with stage 3 chronic kidney disease due to type 1 diabetes mellitus |  | Coronary atherosclerosis due to calcified coronary lesion of native artery |
| Hypertension associated with stage 3 chronic kidney disease due to type 2 diabetes mellitus |  | Coronary atherosclerosis due to lipid rich plaque (CODE) |
| Hypertension associated with stage 3a chronic kidney disease due to type 1 diabetes mellitus |  | Coronary atherosclerosis due to lipid rich plaque |
| Hypertension associated with stage 3a chronic kidney disease due to type 2 diabetes mellitus |  | Coronary atherosclerosis due to severely calcified coronary lesion |
| Hypertension associated with stage 3b chronic kidney disease due to type 1 diabetes mellitus |  | Coronary atherosclerosis of artery bypass graft |
| Hypertension associated with stage 3b chronic kidney disease due to type 2 diabetes mellitus |  | Coronary atherosclerosis of artery of transplanted heart |
| Hypertension associated with stage 4 chronic kidney disease due to type 1 diabetes mellitus |  | Coronary atherosclerosis of autologous artery bypass graft |
| Hypertension associated with stage 4 chronic kidney disease due to type 2 diabetes mellitus |  | Coronary atherosclerosis of autologous artery bypass graft without angina |
| Hypertension associated with stage 5 chronic kidney disease due to type 1 diabetes mellitus |  | Coronary atherosclerosis of autologous bypass graft |
| Hypertension associated with stage 5 chronic kidney disease due to type 2 diabetes mellitus |  | Coronary atherosclerosis of autologous vein bypass graft |
| Hypertension associated with transplantation |  | Coronary atherosclerosis of autologous vein bypass graft without angina |
| Hypertension associated with type 1 diabetes mellitus |  | Coronary atherosclerosis of bypass artery of transplanted heart |
| Hypertension associated with type 2 diabetes mellitus |  | Coronary atherosclerosis of bypass graft |
| Hypertension complicating childbirth |  | Coronary atherosclerosis of bypass graft of transplanted heart |
| Hypertension complicating diabetes |  | Coronary atherosclerosis of bypass vein of transplanted heart |
| Hypertension complicating pregnancy |  | Coronary atherosclerosis of coronary artery of transplanted heart |
| Hypertension complicating pregnancy in first trimester |  | Coronary atherosclerosis of internal mammary artery bypass graft |
| Hypertension complicating pregnancy in second trimester |  | Coronary atherosclerosis of native artery of transplanted heart |
| Hypertension complicating pregnancy in third trimester |  | Coronary atherosclerosis of native coronary artery |
| Hypertension complicating pregnancy, childbirth and puerperium with baby delivered and postnatal complication |  | Coronary atherosclerosis of native coronary artery of transplanted heart |
| Hypertension complicating pregnancy, childbirth, and the puerperium |  | Coronary atherosclerosis of native coronary vessel |
| Hypertension complicating pregnancy, childbirth, or puerperium |  | Coronary atherosclerosis of nonautologous biological bypass graft |
| Hypertension complicating pregnancy, delivered-current hospitalization |  | Coronary atherosclerosis of transplanted heart |
| Hypertension complicating pregnancy, delivered, current hospitalization |  | Coronary atherosclerosis of unspecified type of bypass graft |
| Hypertension complicating pregnancy, first trimester |  | Coronary atherosclerosis of unspecified type of vessel, native or graft |
| Hypertension complicating pregnancy, second trimester |  | Coronary atherosclerosis of vein bypass graft |
| Hypertension complicating pregnancy, third trimester |  | Coronary atherosclerosis transplanted heart |
| Hypertension complicating pregnancy, unspecified trimester |  | Coronary disease |
| Hypertension complications |  | Coronary heart disease |
| Hypertension due to arteriovenous shunt |  | Coronary ostial sclerosis |
| Hypertension due to AV shunt |  | Coronary sclerosis |
| Hypertension due to catecholamine excess |  | Diastolic heart failure secondary to coronary artery disease |
| Hypertension due to compression of parenchyma of kidney |  | Diastolic heart failure secondary to coronary artery disease, acute |
| Hypertension due to compression of renal parenchyma |  | Diastolic heart failure secondary to coronary artery disease, acute on chronic |
| Hypertension due to congenital adrenal hyperplasia |  | Diastolic heart failure secondary to coronary artery disease, chronic |
| Hypertension due to drug |  | Diastolic heart failure secondary to coronary artery disease, unspecified failure chronicity |
| Hypertension due to end stage renal disease caused by type 1 diabetes mellitus, on dialysis |  | Double vessel coronary artery disease |
| Hypertension due to end stage renal disease caused by type 2 diabetes mellitus, on dialysis |  | Failed CABG (coronary artery bypass graft) |
| Hypertension due to end stage renal disease on dialysis |  | Failed CABG (coronary artery bypass graft), initial encounter |
| Hypertension due to endocrine disorder |  | Failed CABG (coronary artery bypass graft), sequela |
| Hypertension due to gain-of-function mutation in mineralocorticoid receptor gene |  | Failed CABG (coronary artery bypass graft), subsequent encounter |
| Hypertension due to hormonal excess |  | Failed coronary artery bypass graft |
| Hypertension due to intracranial pressure |  | Failed coronary artery bypass graft, initial encounter |
| Hypertension due to kidney disease complicating or reason for care during puerperium |  | Failed coronary artery bypass graft, sequela |
| Hypertension due to kidney disease during pregnancy, antepartum |  | Failed coronary artery bypass graft, subsequent encounter |
| Hypertension due to kidney transplant |  | Hardening of bypass graft of coronary artery |
| Hypertension due to obstruction of aortic arch |  | Hardening of the arteries of the heart |
| Hypertension due to reduced activity of kallikrein-kinin system |  | Heart failure, diastolic, due to CAD |
| Hypertension due to surgical care |  | Heart failure, diastolic, due to CAD, acute |
| Hypertension during pregnancy |  | Heart failure, diastolic, due to CAD, acute on chronic |
| Hypertension during pregnancy in first trimester |  | Heart failure, diastolic, due to CAD, chronic |
| Hypertension during pregnancy in first trimester, antepartum |  | Heart failure, diastolic, due to CAD, unspecified failure chronicity |
| Hypertension during pregnancy in first trimester, unspecified hypertension in pregnancy |  | Heart failure, systolic, due to CAD |
| Hypertension during pregnancy in first trimester, unspecified hypertension in pregnancy type |  | Ischemic heart disease due to obstructive coronary atherosclerosis |
| Hypertension during pregnancy in second trimester |  | Left main coronary artery disease |
| Hypertension during pregnancy in second trimester, antepartum |  | Maternal coronary artery disease affecting pregnancy in second trimester, antepartum |
| Hypertension during pregnancy in second trimester, unspecified hypertension in pregnancy |  | Maternal coronary artery disease affecting pregnancy in third trimester, antepartum |
| Hypertension during pregnancy in second trimester, unspecified hypertension in pregnancy type |  | Maternal coronary artery disease affecting pregnancy, antepartum |
| Hypertension during pregnancy in third trimester |  | Maternal coronary artery disease affecting pregnancy, antepartum, first trimester |
| Hypertension during pregnancy in third trimester, antepartum |  | Maternal coronary artery disease affecting pregnancy, antepartum, second trimester |
| Hypertension during pregnancy in third trimester, unspecified hypertension in pregnancy |  | Maternal coronary artery disease affecting pregnancy, antepartum, third trimester |
| Hypertension during pregnancy in third trimester, unspecified hypertension in pregnancy type |  | Maternal coronary artery disease affecting pregnancy, antepartum, unspecified trimester |
| Hypertension during pregnancy, antepartum |  | Maternal coronary artery disease complicating pregnancy |
| Hypertension during pregnancy, antepartum, first trimester |  | Maternal coronary artery disease complicating pregnancy in first trimester |
| Hypertension during pregnancy, antepartum, second trimester |  | Maternal coronary artery disease complicating pregnancy in second trimester |
| Hypertension during pregnancy, antepartum, third trimester |  | Maternal coronary artery disease complicating pregnancy in third trimester |
| Hypertension during pregnancy, antepartum, unspecified hypertension in pregnancy type |  | Maternal coronary artery disease complicating pregnancy, antepartum |
| Hypertension during pregnancy, antepartum, unspecified trimester |  | Maternal coronary artery disease complicating pregnancy, first trimester |
| Hypertension during pregnancy, first trimester |  | Maternal coronary artery disease complicating pregnancy, second trimester |
| Hypertension during pregnancy, second trimester |  | Maternal coronary artery disease complicating pregnancy, third trimester |
| Hypertension during pregnancy, third trimester |  | Maternal coronary artery disease complicating pregnancy, unspecified trimester |
| Hypertension during pregnancy, unspecified hypertension in pregnancy, unspecified trimester |  | MEF2A-related coronary artery disease 1, autosomal dominant |
| Hypertension during pregnancy, unspecified trimester |  | Mild CAD |
| Hypertension during puerperium with pulmonary edema |  | Mild CAD without angina pectoris |
| Hypertension following surgery |  | Mild coronary artery disease |
| Hypertension goal BP (blood pressure) < 130/80 |  | Mild coronary artery disease without angina pectoris |
| Hypertension goal BP (blood pressure) < 140/80 |  | Multiple vessel coronary artery disease |
| Hypertension goal BP (blood pressure) < 140/90 |  | Myocardial disease (with mention of arteriosclerosis) |
| Hypertension goal BP (blood pressure) < 150/90 |  | Non-occlusive coronary artery disease |
| Hypertension in antepartum pregnancy |  | Non-occlusive coronary artery disease requiring drug therapy |
| Hypertension in antepartum pregnancy, first trimester |  | Nonobstructive atherosclerosis of coronary artery |
| Hypertension in antepartum pregnancy, second trimester |  | Nonocclusive coronary atherosclerosis of native coronary artery |
| Hypertension in antepartum pregnancy, third trimester |  | Obliterative coronary artery disease |
| Hypertension in antepartum pregnancy, unspecified trimester |  | Occlusive coronary artery disease |
| Hypertension in child |  | Occlusive coronary artery disease requiring drug therapy |
| Hypertension in child age 0-18 |  | Pre-clinical coronary artery disease |
| Hypertension in chronic kidney disease due to type 1 diabetes mellitus |  | Preclinical coronary artery disease |
| Hypertension in chronic kidney disease due to type 2 diabetes mellitus |  | Premature coronary artery disease |
| Hypertension in delivered pregnancy |  | Presence of stent in coronary artery in patient with coronary artery disease |
| Hypertension in obstetric context |  | Radiation-induced coronary artery disease |
| Hypertension in obstetric context in first trimester |  | Recurrent coronary arteriosclerosis after percutaneous transluminal coronary angioplasty |
| Hypertension in obstetric context in second trimester |  | Recurrent coronary arteriosclerosis following PTCA |
| Hypertension in obstetric context in third trimester |  | Significant disease of coronary bypass graft |
| Hypertension in obstetric context, antepartum |  | SILENT ISCHEMIA |
| Hypertension in obstetric context, first trimester |  | Single vessel coronary artery disease |
| Hypertension in obstetric context, second trimester |  | Single vessel coronary disease |
| Hypertension in obstetric context, third trimester |  | Small vessel coronary artery disease |
| Hypertension in obstetric context, unspecified trimester |  | ST elevation (STEMI) myocardial infarction involving left main coronary artery |
| Hypertension in pregnancy |  | ST elevation myocardial infarction (STEMI) involving left main coronary artery with complication |
| Hypertension in pregnancy, antepartum |  | ST elevation myocardial infarction involving left main coronary artery |
| Hypertension in pregnancy, antepartum, first trimester |  | STEMI involving left main coronary artery |
| Hypertension in pregnancy, antepartum, second trimester |  | Systolic heart failure secondary to coronary artery disease |
| Hypertension in pregnancy, antepartum, third trimester |  | Triple vessel coronary artery disease |
| Hypertension in pregnancy, antepartum, unspecified trimester |  | Triple vessel disease of the heart |
| Hypertension in pregnancy, benign essential, delivered |  | Two-vessel coronary artery disease |
| Hypertension in pregnancy, delivered |  | Under observation for suspected coronary artery disease |
| Hypertension in pregnancy, delivered with postpartum condition |  | Unstable angina due to arteriosclerosis of coronary artery bypass graft |
| Hypertension in pregnancy, essential |  | Unstable angina pectoris due to coronary arteriosclerosis |
| Hypertension in pregnancy, essential, antepartum |  |  |
| Hypertension in pregnancy, essential, antepartum, first trimester |  |  |
| Hypertension in pregnancy, essential, antepartum, second trimester |  |  |
| Hypertension in pregnancy, essential, antepartum, third trimester |  |  |
| Hypertension in pregnancy, essential, antepartum, unspecified trimester |  |  |
| Hypertension in pregnancy, essential, delivered |  |  |
| Hypertension in pregnancy, essential, delivered w postpartum condition |  |  |
| Hypertension in pregnancy, essential, postpartum condition |  |  |
| Hypertension in pregnancy, first trimester |  |  |
| Hypertension in pregnancy, postpartum condition |  |  |
| Hypertension in pregnancy, pre-eclampsia |  |  |
| Hypertension in pregnancy, pre-eclampsia, delivered |  |  |
| Hypertension in pregnancy, pre-eclampsia, severe |  |  |
| Hypertension in pregnancy, pre-eclampsia, severe, antepartum |  |  |
| Hypertension in pregnancy, pre-eclampsia, severe, delivered |  |  |
| Hypertension in pregnancy, pre-eclampsia, severe, delivered/postpartum |  |  |
| Hypertension in pregnancy, pre-eclampsia, severe, postpartum condition |  |  |
| Hypertension in pregnancy, pre-existing |  |  |
| Hypertension in pregnancy, pre-existing, antepartum |  |  |
| Hypertension in pregnancy, pre-existing, antepartum, first trimester |  |  |
| Hypertension in pregnancy, pre-existing, antepartum, second trimester |  |  |
| Hypertension in pregnancy, pre-existing, antepartum, third trimester |  |  |
| Hypertension in pregnancy, pre-existing, antepartum, unspecified trimester |  |  |
| Hypertension in pregnancy, pre-existing, delivered w postpartum condit |  |  |
| Hypertension in pregnancy, pre-existing, postpartum complication |  |  |
| Hypertension in pregnancy, pre-existing, with delivery |  |  |
| Hypertension in pregnancy, preeclampsia |  |  |
| Hypertension in pregnancy, preeclampsia, delivered |  |  |
| Hypertension in pregnancy, preeclampsia, second trimester |  |  |
| Hypertension in pregnancy, preeclampsia, severe |  |  |
| Hypertension in pregnancy, preeclampsia, severe, antepartum |  |  |
| Hypertension in pregnancy, preeclampsia, severe, antepartum, second trimester |  |  |
| Hypertension in pregnancy, preeclampsia, severe, antepartum, third trimester |  |  |
| Hypertension in pregnancy, preeclampsia, severe, antepartum, unspecified trimester |  |  |
| Hypertension in pregnancy, preeclampsia, severe, delivered |  |  |
| Hypertension in pregnancy, preeclampsia, severe, delivered/postpartum |  |  |
| Hypertension in pregnancy, preeclampsia, severe, postpartum condition |  |  |
| Hypertension in pregnancy, preeclampsia, severe, second trimester |  |  |
| Hypertension in pregnancy, preeclampsia, severe, third trimester |  |  |
| Hypertension in pregnancy, preeclampsia, severe, unspecified trimester |  |  |
| Hypertension in pregnancy, preeclampsia, third trimester |  |  |
| Hypertension in pregnancy, preeclampsia, unspecified trimester |  |  |
| Hypertension in pregnancy, preeclampsia/eclampsia/prior HTN/deliv |  |  |
| Hypertension in pregnancy, preeclampsia/eclampsia/prior HTN/deliv/post |  |  |
| Hypertension in pregnancy, preeclampsia/eclampsia/prior HTN/postp cond |  |  |
| Hypertension in pregnancy, renal |  |  |
| Hypertension in pregnancy, renal, antepartum |  |  |
| Hypertension in pregnancy, renal, antepartum, first trimester |  |  |
| Hypertension in pregnancy, renal, antepartum, second trimester |  |  |
| Hypertension in pregnancy, renal, antepartum, third trimester |  |  |
| Hypertension in pregnancy, renal, antepartum, unspecified trimester |  |  |
| Hypertension in pregnancy, renal, delivered |  |  |
| Hypertension in pregnancy, renal, delivered with postpartum condition |  |  |
| Hypertension in pregnancy, second trimester |  |  |
| Hypertension in pregnancy, third trimester |  |  |
| Hypertension in pregnancy, transient |  |  |
| Hypertension in pregnancy, transient, antepartum |  |  |
| Hypertension in pregnancy, transient, antepartum, first trimester |  |  |
| Hypertension in pregnancy, transient, antepartum, second trimester |  |  |
| Hypertension in pregnancy, transient, antepartum, third trimester |  |  |
| Hypertension in pregnancy, transient, antepartum, unspecified trimester |  |  |
| Hypertension in pregnancy, transient, delivered |  |  |
| Hypertension in pregnancy, transient, delivered w postpartum condition |  |  |
| Hypertension in pregnancy, transient, delivered with postpartum complication |  |  |
| Hypertension in pregnancy, transient, first trimester |  |  |
| Hypertension in pregnancy, transient, postpartum |  |  |
| Hypertension in pregnancy, transient, postpartum condition |  |  |
| Hypertension in pregnancy, transient, second trimester |  |  |
| Hypertension in pregnancy, transient, third trimester |  |  |
| Hypertension in pregnancy, transient, unspecified trimester |  |  |
| Hypertension in pregnancy, unspecified trimester |  |  |
| Hypertension in stage 2 chronic kidney disease due to type 2 diabetes mellitus |  |  |
| Hypertension in stage 3 chronic kidney disease due to type 2 diabetes mellitus |  |  |
| Hypertension in stage 4 chronic kidney disease due to type 2 diabetes mellitus |  |  |
| Hypertension in stage 5 chronic kidney disease due to type 2 diabetes mellitus |  |  |
| Hypertension not at goal |  |  |
| Hypertension of pregnancy, transient |  |  |
| Hypertension of pregnancy, transient, antepartum |  |  |
| Hypertension of pregnancy, transient, antepartum, first trimester |  |  |
| Hypertension of pregnancy, transient, antepartum, second trimester |  |  |
| Hypertension of pregnancy, transient, antepartum, third trimester |  |  |
| Hypertension of pregnancy, transient, antepartum, unspecified trimester |  |  |
| Hypertension of pregnancy, transient, delivered |  |  |
| Hypertension of pregnancy, transient, delivered w postpartum condition |  |  |
| Hypertension of pregnancy, transient, first trimester |  |  |
| Hypertension of pregnancy, transient, postpartum condition |  |  |
| Hypertension of pregnancy, transient, second trimester |  |  |
| Hypertension of pregnancy, transient, third trimester |  |  |
| Hypertension of pregnancy, transient, unspecified trimester |  |  |
| Hypertension requiring medication after live kidney donation |  |  |
| Hypertension resistant to drug therapy |  |  |
| Hypertension secondary to drug |  |  |
| Hypertension secondary to endocrine disorder with goal blood pressure less than 130/80 |  |  |
| Hypertension secondary to endocrine disorder with goal blood pressure less than 130/85 |  |  |
| Hypertension secondary to endocrine disorder with goal blood pressure less than 140/90 |  |  |
| Hypertension secondary to endocrine disorder with goal blood pressure less than 150/90 |  |  |
| Hypertension secondary to endocrine disorders |  |  |
| Hypertension secondary to endocrine disorders, benign or unspecified |  |  |
| Hypertension secondary to endocrine disorders, malignant |  |  |
| Hypertension secondary to kidney disease affecting pregnancy |  |  |
| Hypertension secondary to other kidney disorders |  |  |
| Hypertension secondary to other renal disorders (CODE) |  |  |
| Hypertension secondary to other renal disorders |  |  |
| Hypertension secondary to other renal disorders, benign or unspecified |  |  |
| Hypertension secondary to other renal disorders, malignant |  |  |
| Hypertension secondary to renal disease affecting pregnancy in first trimester |  |  |
| Hypertension secondary to renal disease affecting pregnancy in second trimester |  |  |
| Hypertension secondary to renal disease affecting pregnancy in third trimester |  |  |
| Hypertension secondary to renal disease affecting pregnancy, antepartum |  |  |
| Hypertension secondary to renal disease affecting pregnancy, childbirth, or puerperium |  |  |
| Hypertension secondary to renal disease complicating childbirth |  |  |
| Hypertension secondary to renal disease complicating or reason for care during puerperium |  |  |
| Hypertension secondary to renal disease complicating pregnancy |  |  |
| Hypertension secondary to renal disease complicating pregnancy, first trimester |  |  |
| Hypertension secondary to renal disease complicating pregnancy, second trimester |  |  |
| Hypertension secondary to renal disease complicating pregnancy, third trimester |  |  |
| Hypertension secondary to renal disease complicating pregnancy, unspecified trimester |  |  |
| Hypertension secondary to renal disease complicating puerperium |  |  |
| Hypertension secondary to renal disease during pregnancy in second trimester, antepartum |  |  |
| Hypertension secondary to renal disease during pregnancy in third trimester, antepartum |  |  |
| Hypertension secondary to renal disease during pregnancy, antepartum |  |  |
| Hypertension secondary to renal disease during pregnancy, antepartum, first trimester |  |  |
| Hypertension secondary to renal disease during pregnancy, antepartum, second trimester |  |  |
| Hypertension secondary to renal disease during pregnancy, antepartum, third trimester |  |  |
| Hypertension secondary to renal disease during pregnancy, antepartum, unspecified trimester |  |  |
| Hypertension secondary to renal disease in first trimester |  |  |
| Hypertension secondary to renal disease in first trimester, antepartum |  |  |
| Hypertension secondary to renal disease in obstetric context |  |  |
| Hypertension secondary to renal disease in pregnancy |  |  |
| Hypertension secondary to renal disease in pregnancy, antepartum |  |  |
| Hypertension secondary to renal disease in pregnancy, antepartum, first trimester |  |  |
| Hypertension secondary to renal disease in pregnancy, antepartum, second trimester |  |  |
| Hypertension secondary to renal disease in pregnancy, antepartum, third trimester |  |  |
| Hypertension secondary to renal disease in pregnancy, antepartum, unspecified trimester |  |  |
| Hypertension secondary to renal disease in pregnancy, deliver/postpart |  |  |
| Hypertension secondary to renal disease in pregnancy, delivered |  |  |
| Hypertension secondary to renal disease in second trimester |  |  |
| Hypertension secondary to renal disease in third trimester |  |  |
| Hypertension secondary to renal disease, antepartum |  |  |
| Hypertension secondary to renal disease, antepartum, first trimester |  |  |
| Hypertension secondary to renal disease, antepartum, second trimester |  |  |
| Hypertension secondary to renal disease, antepartum, third trimester |  |  |
| Hypertension secondary to renal disease, antepartum, unspecified trimester |  |  |
| Hypertension secondary to renal disease, complicating pregnancy, childbirth, and the puerperium |  |  |
| Hypertension secondary to renal disease, complicating pregnancy, childbirth, and the puerperium, unspecified as to episode of care |  |  |
| Hypertension secondary to renal disease, postpartum |  |  |
| Hypertension secondary to renal disease, with delivery |  |  |
| Hypertension secondary to renal disease, with delivery, with current postpartum complication |  |  |
| Hypertension secondary to renal disease, with delivery, with mention of postpartum complication |  |  |
| Hypertension with albuminuria |  |  |
| Hypertension with baby delivered with postpartum complication |  |  |
| Hypertension with congestive heart failure and renal failure |  |  |
| Hypertension with delivery |  |  |
| Hypertension with fluid overload |  |  |
| Hypertension with goal blood pressure less than 120/80 |  |  |
| Hypertension with goal blood pressure less than 130/80 |  |  |
| Hypertension with goal blood pressure less than 130/85 |  |  |
| Hypertension with goal blood pressure less than 140/80 |  |  |
| Hypertension with goal blood pressure less than 140/90 |  |  |
| Hypertension with goal of symptom management only |  |  |
| Hypertension with goal to be determined |  |  |
| Hypertension with heart disease |  |  |
| Hypertension with hypervolemia |  |  |
| Hypertension with impaired renal function |  |  |
| Hypertension with intolerance to multiple antihypertensive drugs |  |  |
| Hypertension with mineralocorticoid excess |  |  |
| Hypertension with normal renal function |  |  |
| Hypertension with renal disease |  |  |
| Hypertension with target blood pressure goal under 150/90 |  |  |
| Hypertension with target organ involvement |  |  |
| Hypertension without albuminuria or edema in obstetric context |  |  |
| Hypertension without albuminuria or edema in obstetric context, antepartum |  |  |
| Hypertension without albuminuria or edema in the obstetric context |  |  |
| Hypertension without albuminuria or edema in the obstetric context, first trimester |  |  |
| Hypertension without albuminuria or edema in the obstetric context, second trimester |  |  |
| Hypertension without albuminuria or edema in the obstetric context, third trimester |  |  |
| Hypertension without albuminuria or edema in the obstetric context, unspecified trimester |  |  |
| Hypertension without cardiac signs or symptoms |  |  |
| Hypertension-nephrosis syndrome due to secondary diabetes |  |  |
| Hypertension, accelerated |  |  |
| Hypertension, accelerated, with diastolic congestive heart failure, NYHA class 1 |  |  |
| Hypertension, accelerated, with systolic CHF, NYHA class 4 |  |  |
| Hypertension, benign |  |  |
| Hypertension, benign essential, age 0-18 |  |  |
| Hypertension, benign essential, goal below 140/90 |  |  |
| Hypertension, benign renovascular |  |  |
| Hypertension, benign, secondary |  |  |
| Hypertension, diastolic |  |  |
| Hypertension, diastolic, benign |  |  |
| Hypertension, diastolic, labile |  |  |
| Hypertension, diastolic, secondary |  |  |
| Hypertension, diastolic, sustained |  |  |
| Hypertension, essential |  |  |
| Hypertension, essential, benign |  |  |
| Hypertension, essential, complicating pregnancy |  |  |
| Hypertension, essential, complicating pregnancy, first trimester |  |  |
| Hypertension, essential, complicating pregnancy, second trimester |  |  |
| Hypertension, essential, complicating pregnancy, third trimester |  |  |
| Hypertension, essential, complicating pregnancy, unspecified trimester |  |  |
| Hypertension, essential, malignant, with systolic CHF, NYHA class 1 |  |  |
| Hypertension, goal below 140/80 |  |  |
| Hypertension, goal below 140/90 |  |  |
| Hypertension, goal below 150/90 |  |  |
| Hypertension, goal to be determined |  |  |
| Hypertension, Goldblatt |  |  |
| Hypertension, isolated systolic |  |  |
| Hypertension, malignant |  |  |
| Hypertension, malignant renovascular |  |  |
| Hypertension, malignant, secondary |  |  |
| Hypertension, malignant, with acute intensive management |  |  |
| Hypertension, pediatric |  |  |
| Hypertension, postpartum condition or complication |  |  |
| Hypertension, pre-transplant |  |  |
| Hypertension, renal |  |  |
| Hypertension, renal, stage 1-4 or unspecified chronic kidney disease |  |  |
| Hypertension, renal, stage 5 chronic kidney disease or end stage renal disease |  |  |
| Hypertension, renovascular |  |  |
| Hypertension, renovascular, malignant, with acute intensive management |  |  |
| Hypertension, renovascular, severity unknown |  |  |
| Hypertension, secondary |  |  |
| Hypertension, surgical complication |  |  |
| Hypertension, uncontrolled |  |  |
| Hypertension, unspecified type |  |  |
| Hypertension, with delivery, with current postpartum complication |  |  |
| Hypertensive arteriosclerotic cardiovascular disease |  |  |
| Hypertensive congenital adrenal hyperplasia |  |  |
| Hypertensive crisis |  |  |
| Hypertensive crisis without congestive heart failure |  |  |
| Hypertensive crisis, unspecified |  |  |
| Hypertensive disease |  |  |
| Hypertensive disease arising during pregnancy |  |  |
| Hypertensive disorder |  |  |
| Hypertensive emergency |  |  |
| Hypertensive emergency without congestive heart failure |  |  |
| Hypertensive emergency, no CHF |  |  |
| Hypertensive renal sclerosis with hypertension |  |  |
| Hypertensive renal sclerosis with hypertension, stage 1-4 or unspecified chronic kidney disease |  |  |
| Hypertensive renal sclerosis with hypertension, stage 5 chronic kidney disease or end stage renal disease (CMS/HCC) |  |  |
| Hypertensive urgency |  |  |
| Hypertensive urgency, malignant |  |  |
| Hypertensive vascular degeneration |  |  |
| Hypertensive vascular disease |  |  |
| Hypertensive vascular disease during pregnancy, antepartum |  |  |
| Hypomagnesemia, hypertension, and hypercholesterolemia associated with mutation in MT-TI gene |  |  |
| Idiopathic hypertension |  |  |
| Immunosuppressive-induced high blood pressure |  |  |
| Impending eclampsia |  |  |
| Impending eclampsia in second trimester |  |  |
| Impending eclampsia in third trimester |  |  |
| Impending eclampsia, antepartum |  |  |
| Impending eclampsia, second trimester |  |  |
| Impending eclampsia, third trimester |  |  |
| Impending eclampsia, unspecified trimester |  |  |
| Increased blood pressure and edema during pregnancy |  |  |
| Intermittent hypertension |  |  |
| Intracranial hemorrhage, spontaneous intraparenchymal, associated with hypertension, acute |  |  |
| Intracranial hemorrhage, spontaneous intraparenchymal, associated with hypertension, in utero |  |  |
| Intracranial hemorrhage, spontaneous intraparenchymal, associated with hypertension, perinatal |  |  |
| Intracranial hemorrhage, spontaneous intraparenchymal, associated with hypertension, presumed perinatal |  |  |
| Intracranial hemorrhage, spontaneous intraventricular, associated with hypertension, acute |  |  |
| Intracranial hemorrhage, spontaneous intraventricular, associated with hypertension, chronic |  |  |
| Intracranial hemorrhage, spontaneous intraventricular, associated with hypertension, in utero |  |  |
| Intracranial hemorrhage, spontaneous intraventricular, associated with hypertension, perinatal |  |  |
| Intracranial hemorrhage, spontaneous intraventricular, associated with hypertension, presumed perinatal |  |  |
| Intracranial hemorrhage, spontaneous subarachnoid, associated with hypertension, acute |  |  |
| Intracranial hemorrhage, spontaneous subarachnoid, associated with hypertension, chronic |  |  |
| Intracranial hemorrhage, spontaneous subarachnoid, associated with hypertension, in utero |  |  |
| Intracranial hemorrhage, spontaneous subarachnoid, associated with hypertension, perinatal |  |  |
| Intracranial hemorrhage, spontaneous subarachnoid, associated with hypertension, presumed perinatal |  |  |
| Kidney disease with benign hypertension |  |  |
| Labile diastolic hypertension |  |  |
| Labile essential hypertension |  |  |
| Labile hypertension |  |  |
| Labile hypertension due to clinical environment |  |  |
| Labile secondary hypertension |  |  |
| Late postpartum pre-eclampsia with posterior leukoencephalopathy syndrome |  |  |
| Late postpartum pre-eclampsia with posterior reversible encephalopathy syndrome |  |  |
| Late postpartum preeclampsia with posterior leukoencephalopathy syndrome |  |  |
| Late postpartum preeclampsia with posterior reversible encephalopathy syndrome |  |  |
| Low-renin essential hypertension |  |  |
| Malignant diastolic hypertension with diastolic congestive heart failure, NYHA class 2 |  |  |
| Malignant diastolic hypertension without heart disease |  |  |
| Malignant essential hypertension |  |  |
| Malignant essential hypertension with CHF w/o chronic kidney disease |  |  |
| Malignant essential hypertension with CHF with renal disease |  |  |
| Malignant essential hypertension with CHF without renal disease |  |  |
| Malignant essential hypertension with CHF, NYHA class 2 |  |  |
| Malignant essential hypertension with combined systolic and diastolic CHF, NYHA class 2 |  |  |
| Malignant essential hypertension with combined systolic and diastolic NYHA class 2 congestive heart failure |  |  |
| Malignant essential hypertension with congestive heart failure with chronic kidney disease |  |  |
| Malignant essential hypertension with congestive heart failure with combined systolic and diastolic dysfunction |  |  |
| Malignant essential hypertension with congestive heart failure with preserved left ventricular function |  |  |
| Malignant essential hypertension with congestive heart failure with reduced left ventricular function |  |  |
| Malignant essential hypertension with congestive heart failure with renal disease |  |  |
| Malignant essential hypertension with congestive heart failure without chronic kidney disease |  |  |
| Malignant essential hypertension with congestive heart failure without renal disease |  |  |
| Malignant essential hypertension with congestive heart failure, NYHA class 2 |  |  |
| Malignant essential hypertension with diastolic congestive heart failure, NYHA class 2 |  |  |
| Malignant essential hypertension with NYHA class 4 systolic heart failure |  |  |
| Malignant essential hypertension with systolic CHF, NYHA class 4 |  |  |
| Malignant essential hypertension with systolic congestive heart failure, NYHA class 1 |  |  |
| Malignant HTN with heart disease, w/o CHF, w/o chronic kidney disease |  |  |
| Malignant HTN with heart disease, w/o CHF, with chronic kidney disease |  |  |
| Malignant hypertension |  |  |
| Malignant hypertension affecting pregnancy |  |  |
| Malignant hypertension affecting pregnancy in first trimester |  |  |
| Malignant hypertension affecting pregnancy in first trimester, antepartum |  |  |
| Malignant hypertension affecting pregnancy in second trimester |  |  |
| Malignant hypertension affecting pregnancy in second trimester, antepartum |  |  |
| Malignant hypertension affecting pregnancy in third trimester |  |  |
| Malignant hypertension affecting pregnancy in third trimester, antepartum |  |  |
| Malignant hypertension affecting pregnancy, antepartum |  |  |
| Malignant hypertension affecting pregnancy, antepartum, first trimester |  |  |
| Malignant hypertension affecting pregnancy, antepartum, second trimester |  |  |
| Malignant hypertension affecting pregnancy, antepartum, third trimester |  |  |
| Malignant hypertension affecting pregnancy, antepartum, unspecified trimester |  |  |
| Malignant hypertension complicating childbirth |  |  |
| Malignant hypertension complicating pregnancy |  |  |
| Malignant hypertension complicating pregnancy, first trimester |  |  |
| Malignant hypertension complicating pregnancy, second trimester |  |  |
| Malignant hypertension complicating pregnancy, third trimester |  |  |
| Malignant hypertension complicating pregnancy, unspecified trimester |  |  |
| Malignant hypertension complicating the puerperium |  |  |
| Malignant hypertension due to endocrine disorder |  |  |
| Malignant hypertension due to increased intracranial pressure |  |  |
| Malignant hypertension due to polycystic kidney |  |  |
| Malignant hypertension due to primary aldosteronism (CMS/HCC) |  |  |
| Malignant hypertension in obstetric context |  |  |
| Malignant hypertension in obstetric context in first trimester |  |  |
| Malignant hypertension in obstetric context in second trimester |  |  |
| Malignant hypertension in obstetric context in third trimester |  |  |
| Malignant hypertension in obstetric context, antepartum |  |  |
| Malignant hypertension in obstetric context, first trimester |  |  |
| Malignant hypertension in obstetric context, second trimester |  |  |
| Malignant hypertension in obstetric context, third trimester |  |  |
| Malignant hypertension in obstetric context, unspecified trimester |  |  |
| Malignant hypertension requiring acute intensive management |  |  |
| Malignant hypertension w/o heart disease, w/o chronic kidney disease |  |  |
| Malignant hypertension w/o heart disease, with chronic kidney disease |  |  |
| Malignant hypertension w/o heart disease, with chronic kidney disease, stage 1-4 or unspecified chronic kidney disease |  |  |
| Malignant hypertension w/o heart disease, with chronic kidney disease, stage 5 chronic kidney disease or end stage renal disease (CMS/HCC) |  |  |
| Malignant hypertension with chronic kidney disease |  |  |
| Malignant hypertension with chronic kidney disease, stage 1 through stage 4 or unspecified chronic kidney disease |  |  |
| Malignant hypertension with chronic kidney disease, stage 1-4 or unspecified chronic kidney disease |  |  |
| Malignant hypertension with chronic kidney disease, stage 5 chronic kidney disease or end stage renal disease |  |  |
| Malignant hypertension with chronic renal disease, stage V or ESRD |  |  |
| Malignant hypertension with heart disease, w/o CHF, w/o renal disease |  |  |
| Malignant hypertension with heart disease, without congestive heart failure, with chronic kidney disease |  |  |
| Malignant hypertension with heart disease, without congestive heart failure, without chronic kidney disease |  |  |
| Malignant hypertension with heart disease, without congestive heart failure, without renal disease |  |  |
| Malignant hypertension with heart failure and chronic kidney dis stage V |  |  |
| Malignant hypertension with heart failure and chronic kidney disease stage I |  |  |
| Malignant hypertension with heart failure and chronic kidney disease stage V |  |  |
| Malignant hypertension with heart failure and stage 2 chronic kidney disease |  |  |
| Malignant hypertension with heart failure and stage 3 chronic kidney disease |  |  |
| Malignant hypertension with heart failure and stage V renal dis |  |  |
| Malignant hypertension with renal disease and congestive heart failure |  |  |
| Malignant hypertension with renal failure and congestive heart failure |  |  |
| Malignant hypertension without cardiac or renal disease |  |  |
| Malignant hypertension without heart disease |  |  |
| Malignant hypertension without heart disease, with chronic kidney disease |  |  |
| Malignant hypertension without heart disease, with chronic kidney disease, stage 1 through stage 4 or unspecified chronic kidney disease |  |  |
| Malignant hypertension without heart disease, with chronic kidney disease, stage 1-4 or unspecified chronic kidney disease |  |  |
| Malignant hypertension without heart disease, with chronic kidney disease, stage 5 chronic kidney disease or end stage renal disease |  |  |
| Malignant hypertension without heart disease, with renal disease |  |  |
| Malignant hypertension without heart disease, with renal disease, stage 1 through stage 4 or unspecified chronic kidney disease |  |  |
| Malignant hypertension without heart disease, with renal disease, stage 1-4 or unspecified chronic kidney disease |  |  |
| Malignant hypertension without heart disease, with renal disease, stage 5 chronic kidney disease or end stage renal disease |  |  |
| Malignant hypertension without heart disease, without chronic kidney disease |  |  |
| Malignant hypertension without renal or heart disease |  |  |
| Malignant hypertension, endocrine |  |  |
| Malignant hypertension, heart failure & chronic kidney dis stage I |  |  |
| Malignant hypertension, heart failure & chronic kidney dis stage II |  |  |
| Malignant hypertension, heart failure & chronic kidney disease stage I |  |  |
| Malignant hypertension, heart failure, and stage III chronic kidney disease |  |  |
| Malignant hypertensive urgency |  |  |
| Malignant reactive hypertension |  |  |
| Malignant renal hypertension, non-vascular |  |  |
| Malignant renal hypertension, non-vascular, stage 1 through stage 4 or unspecified chronic kidney disease |  |  |
| Malignant renal hypertension, non-vascular, stage 1-4 or unspecified chronic kidney disease |  |  |
| Malignant renal hypertension, non-vascular, stage 5 chronic kidney disease or end stage renal disease |  |  |
| Malignant renovascular hypertension |  |  |
| Malignant renovascular hypertension requiring acute intensive management |  |  |
| Malignant secondary hypertension |  |  |
| Malignant secondary hypertension due to renal artery stenosis |  |  |
| Malignant secondary hypertension due to renal disease |  |  |
| Malignant secondary renovascular hypertension |  |  |
| Malignant systolic hypertension with CHF and chronic kidney disease |  |  |
| Malignant systolic hypertension with CHF and chronic kidney disease, stage 1-4 or unspecified chronic kidney disease |  |  |
| Malignant systolic hypertension with CHF and chronic kidney disease, stage 5 chronic kidney disease or end stage renal disease |  |  |
| Malignant systolic hypertension with congestive heart failure and chronic kidney disease |  |  |
| Malignant systolic hypertension with congestive heart failure and chronic kidney disease, stage 1 through stage 4 or unspecified chronic kidney disease |  |  |
| Malignant systolic hypertension with congestive heart failure and chronic kidney disease, stage 1-4 or unspecified chronic kidney disease |  |  |
| Malignant systolic hypertension with congestive heart failure and chronic kidney disease, stage 5 chronic kidney disease or end stage renal disease |  |  |
| Malignant systolic hypertension with heart disease, without congestive heart failure, without chronic kidney disease |  |  |
| Malignant systolic hypertension without heart disease |  |  |
| Malignant systolic hypertension, with congestive heart failure, with combined systolic and diastolic dysfunction |  |  |
| Malignant systolic hypertension, with diastolic congestive heart failure, with preserved left ventricular function |  |  |
| Maternal chronic hypertension |  |  |
| Maternal chronic hypertension in first trimester |  |  |
| Maternal chronic hypertension in second trimester |  |  |
| Maternal chronic hypertension in third trimester |  |  |
| Maternal chronic hypertension, first trimester |  |  |
| Maternal chronic hypertension, second trimester |  |  |
| Maternal chronic hypertension, third trimester |  |  |
| Maternal chronic hypertension, unspecified trimester |  |  |
| Maternal hypertension |  |  |
| Maternal hypertension affecting childbirth |  |  |
| Maternal hypertension affecting puerperium |  |  |
| Maternal hypertension during pregnancy |  |  |
| Maternal hypertension in first trimester |  |  |
| Maternal hypertension in first trimester, antepartum |  |  |
| Maternal hypertension in second trimester |  |  |
| Maternal hypertension in second trimester, antepartum |  |  |
| Maternal hypertension in third trimester |  |  |
| Maternal hypertension in third trimester, antepartum |  |  |
| Maternal hypertension secondary to renal disease, antepartum |  |  |
| Maternal hypertension secondary to renal disease, antepartum, first trimester |  |  |
| Maternal hypertension secondary to renal disease, antepartum, second trimester |  |  |
| Maternal hypertension secondary to renal disease, antepartum, third trimester |  |  |
| Maternal hypertension secondary to renal disease, antepartum, unspecified trimester |  |  |
| Maternal hypertension syndrome |  |  |
| Maternal hypertension syndrome, first trimester |  |  |
| Maternal hypertension syndrome, second trimester |  |  |
| Maternal hypertension syndrome, third trimester |  |  |
| Maternal hypertension syndrome, unspecified trimester |  |  |
| Maternal hypertension with abortive outcome |  |  |
| Maternal hypertension, antepartum |  |  |
| Maternal hypertension, antepartum, first trimester |  |  |
| Maternal hypertension, antepartum, second trimester |  |  |
| Maternal hypertension, antepartum, third trimester |  |  |
| Maternal hypertension, antepartum, unspecified trimester |  |  |
| Maternal hypertension, delivered, with postpartum complication |  |  |
| Maternal hypertension, first trimester |  |  |
| Maternal hypertension, second trimester |  |  |
| Maternal hypertension, third trimester |  |  |
| Maternal hypertension, unspecified trimester |  |  |
| Maternal toxemia |  |  |
| Maternal toxemia, second trimester |  |  |
| Maternal toxemia, third trimester |  |  |
| Maternal toxemia, unspecified trimester |  |  |
| Mild essential hypertension |  |  |
| Mild HTN |  |  |
| Mild hypertension |  |  |
| Mild or unspecified pre-eclampsia |  |  |
| Mild or unspecified pre-eclampsia, antepartum |  |  |
| Mild or unspecified pre-eclampsia, postpartum condition or complication |  |  |
| Mild or unspecified pre-eclampsia, unspecified as to episode of care |  |  |
| Mild or unspecified pre-eclampsia, with delivery |  |  |
| Mild or unspecified pre-eclampsia, with delivery, with current postpartum complication |  |  |
| Mild pre-eclampsia |  |  |
| Mild pre-eclampsia affecting eighth pregnancy |  |  |
| Mild pre-eclampsia affecting fifth pregnancy |  |  |
| Mild pre-eclampsia affecting first pregnancy |  |  |
| Mild pre-eclampsia affecting fourth pregnancy |  |  |
| Mild pre-eclampsia affecting ninth pregnancy |  |  |
| Mild pre-eclampsia affecting second pregnancy |  |  |
| Mild pre-eclampsia affecting seventh pregnancy |  |  |
| Mild pre-eclampsia affecting sixth pregnancy |  |  |
| Mild pre-eclampsia affecting tenth pregnancy |  |  |
| Mild pre-eclampsia affecting third pregnancy |  |  |
| Mild pre-eclampsia delivered |  |  |
| Mild pre-eclampsia in second trimester |  |  |
| Mild pre-eclampsia in second trimester, antepartum |  |  |
| Mild pre-eclampsia in third trimester |  |  |
| Mild pre-eclampsia in third trimester, antepartum |  |  |
| Mild pre-eclampsia with delivery with postpartum complication |  |  |
| Mild pre-eclampsia, antepartum |  |  |
| Mild pre-eclampsia, antepartum, second trimester |  |  |
| Mild pre-eclampsia, antepartum, third trimester |  |  |
| Mild pre-eclampsia, antepartum, unspecified trimester |  |  |
| Mild pre-eclampsia, delivered, current hospitalization |  |  |
| Mild pre-eclampsia, postpartum |  |  |
| Mild pre-eclampsia, second trimester |  |  |
| Mild pre-eclampsia, third trimester |  |  |
| Mild pre-eclampsia, unspecified trimester |  |  |
| Mild pre-eclamptic toxemia |  |  |
| Mild pre-eclamptic toxemia, second trimester |  |  |
| Mild pre-eclamptic toxemia, third trimester |  |  |
| Mild pre-eclamptic toxemia, unspecified trimester |  |  |
| Mild preeclamp-del w/ postpartum complication |  |  |
| Mild preeclampsia |  |  |
| Mild preeclampsia delivered |  |  |
| Mild preeclampsia, second trimester |  |  |
| Mild preeclampsia, third trimester |  |  |
| Mild preeclampsia, unspecified trimester |  |  |
| Mild proteinuric hypertension of pregnancy |  |  |
| Mild proteinuric hypertension of pregnancy, second trimester |  |  |
| Mild proteinuric hypertension of pregnancy, third trimester |  |  |
| Mild proteinuric hypertension of pregnancy, unspecified trimester |  |  |
| Mild secondary hypertension |  |  |
| Mild systolic hypertension |  |  |
| Mild to moderate pre-eclampsia |  |  |
| Mild to moderate pre-eclampsia affecting childbirth |  |  |
| Mild to moderate pre-eclampsia complicating puerperium |  |  |
| Mild to moderate pre-eclampsia in second trimester |  |  |
| Mild to moderate pre-eclampsia in third trimester |  |  |
| Mild to moderate pre-eclampsia with abortive outcome |  |  |
| Mild to moderate pre-eclampsia, antepartum |  |  |
| Mild to moderate pre-eclampsia, antepartum condition or complication |  |  |
| Mild to moderate pre-eclampsia, complicating childbirth |  |  |
| Mild to moderate pre-eclampsia, complicating the puerperium |  |  |
| Mild to moderate pre-eclampsia, delivered |  |  |
| Mild to moderate pre-eclampsia, delivered, with mention of postpartum complication |  |  |
| Mild to moderate pre-eclampsia, delivered, with or without mention of antepartum condition |  |  |
| Mild to moderate pre-eclampsia, delivered, with postpartum complication |  |  |
| Mild to moderate pre-eclampsia, postpartum |  |  |
| Mild to moderate pre-eclampsia, postpartum condition or complication |  |  |
| Mild to moderate pre-eclampsia, second trimester |  |  |
| Mild to moderate pre-eclampsia, third trimester |  |  |
| Mild to moderate pre-eclampsia, unspecified as to episode of care, or not applicable |  |  |
| Mild to moderate pre-eclampsia, unspecified trimester |  |  |
| Mild/moderate preeclampsia/del w comp |  |  |
| Mild/moderate preeclampsia/delivered |  |  |
| Mild/moderate preeclampsia/unspec |  |  |
| Mild/NOS preeclamp-postpartum complication |  |  |
| Mildly increased blood pressure and edema during pregnancy |  |  |
| Moderate diastolic hypertension |  |  |
| Moderate essential hypertension |  |  |
| Moderate hypertension |  |  |
| Moderate pre-eclampsia |  |  |
| Moderate pre-eclampsia in second trimester |  |  |
| Moderate pre-eclampsia in third trimester |  |  |
| Moderate pre-eclampsia, antepartum |  |  |
| Moderate pre-eclampsia, second trimester |  |  |
| Moderate pre-eclampsia, third trimester |  |  |
| Moderate pre-eclampsia, unspecified trimester |  |  |
| Moderate secondary hypertension |  |  |
| Muromonab-CD3-induced hypertension |  |  |
| Neonatal hypertension |  |  |
| Neoral-induced hypertension |  |  |
| Neurogenic hypertension |  |  |
| Nocturnal hypertension |  |  |
| Non-proteinuric hypertension of pregnancy |  |  |
| Non-vascular benign renal hypertension |  |  |
| Obesity, diabetes, and hypertension syndrome |  |  |
| Office hypertension |  |  |
| Old hyperten preg-unspec |  |  |
| Old hyperten-deliv w/ postpartum complication |  |  |
| On treatment for hypertension |  |  |
| Other complication of benign secondary hypertension |  |  |
| Other complication of malignant secondary hypertension |  |  |
| Other pre-existing hypertension affecting pregnancy, childbirth, or puerperium |  |  |
| Other pre-existing hypertension complicating pregnancy, childbirth, and the puerperium |  |  |
| Other pre-existing hypertension complicating pregnancy, childbirth, and the puerperium, unspecified as to episode of care |  |  |
| Other pre-existing hypertension, antepartum |  |  |
| Other pre-existing hypertension, postpartum condition or complication |  |  |
| Other pre-existing hypertension, with delivery |  |  |
| Other pre-existing hypertension, with delivery, with current postpartum complication |  |  |
| Other secondary hypertension |  |  |
| Other secondary hypertension, benign |  |  |
| Other secondary hypertension, benign or unspecified |  |  |
| Other secondary hypertension, malignant |  |  |
| Other secondary hypertension, unspecified |  |  |
| Other secondary malignant hypertension |  |  |
| Page kidney |  |  |
| Parenchymal renal hypertension |  |  |
| Parenchymal renal hypertension, stage 1 through stage 4 or unspecified chronic kidney disease |  |  |
| Parenchymal renal hypertension, stage 1-4 or unspecified chronic kidney disease |  |  |
| Parenchymal renal hypertension, stage 5 chronic kidney disease or end stage renal disease |  |  |
| Paroxysmal hypertension |  |  |
| PE (pre-eclampsia) |  |  |
| Pe (pre-eclampsia), second trimester |  |  |
| Pe (pre-eclampsia), third trimester |  |  |
| Pe (pre-eclampsia), unspecified trimester |  |  |
| Pediatric hypertension |  |  |
| Perioperative hypertension |  |  |
| PET (mild pre-eclamptic toxemia) |  |  |
| Pet (mild pre-eclamptic toxemia), second trimester |  |  |
| Pet (mild pre-eclamptic toxemia), third trimester |  |  |
| Pet (mild pre-eclamptic toxemia), unspecified trimester |  |  |
| PET (pre-eclamptic toxemia) |  |  |
| Pet (pre-eclamptic toxemia), second trimester |  |  |
| Pet (pre-eclamptic toxemia), third trimester |  |  |
| Pet (pre-eclamptic toxemia), unspecified trimester |  |  |
| PET (severe pre-eclamptic toxemia) |  |  |
| Pet (severe pre-eclamptic toxemia), second trimester |  |  |
| Pet (severe pre-eclamptic toxemia), third trimester |  |  |
| Pet (severe pre-eclamptic toxemia), unspecified trimester |  |  |
| PIH (pregnancy induced hypertension) |  |  |
| PIH (pregnancy induced hypertension), antepartum |  |  |
| PIH (pregnancy induced hypertension), first trimester |  |  |
| PIH (pregnancy induced hypertension), second trimester |  |  |
| PIH (pregnancy induced hypertension), third trimester |  |  |
| PIH (pregnancy induced hypertension), unspecified trimester |  |  |
| Postoperative hypertension |  |  |
| Postpartum essential hypertension during pregnancy, delivered |  |  |
| Postpartum hypertension |  |  |
| Postpartum pre-eclampsia with posterior leukoencephalopathy syndrome |  |  |
| Postpartum pre-eclampsia with posterior reversible encephalopathy syndrome |  |  |
| Postpartum preeclampsia with posterior leukoencephalopathy syndrome |  |  |
| Postprocedural hypertension |  |  |
| Pre-eclampsia |  |  |
| Pre-eclampsia added to pre-existing hypertension |  |  |
| Pre-eclampsia affecting childbirth |  |  |
| Pre-eclampsia affecting pregnancy with pre-existing hypertension, delivered, current hospitalization |  |  |
| Pre-eclampsia affecting pregnancy, antepartum |  |  |
| Pre-eclampsia affecting puerperium |  |  |
| Pre-eclampsia complicating hypertension |  |  |
| Pre-eclampsia complicating pregnancy, with pre-existing hypertension, with delivery, current hospitalisation |  |  |
| Pre-eclampsia complicating pregnancy, with pre-existing hypertension, with delivery, current hospitalization |  |  |
| Pre-eclampsia during pregnancy in second trimester, antepartum |  |  |
| Pre-eclampsia during pregnancy in third trimester, antepartum |  |  |
| Pre-eclampsia in postpartum period |  |  |
| Pre-eclampsia in puerperium |  |  |
| Pre-eclampsia in second trimester |  |  |
| Pre-eclampsia in third trimester |  |  |
| Pre-eclampsia or eclampsia superimposed on pre-existing hypertension |  |  |
| Pre-eclampsia or eclampsia superimposed on pre-existing hypertension, antepartum |  |  |
| Pre-eclampsia or eclampsia superimposed on pre-existing hypertension, complicating pregnancy, childbirth, or the puerperium |  |  |
| Pre-eclampsia or eclampsia superimposed on pre-existing hypertension, complicating pregnancy, childbirth, or the puerperium, unspecified as to episode of care |  |  |
| Pre-eclampsia or eclampsia superimposed on pre-existing hypertension, postpartum |  |  |
| Pre-eclampsia or eclampsia superimposed on pre-existing hypertension, with delivery |  |  |
| Pre-eclampsia or eclampsia superimposed on pre-existing hypertension, with delivery, with current postpartum complication |  |  |
| Pre-eclampsia or eclampsia superimposed on pre-existing hypertension, with delivery, with mention of postpartum complication |  |  |
| Pre-eclampsia or eclampsia with pre-existing hypertension with postnatal complication |  |  |
| Pre-eclampsia superimposed on chronic hypertension |  |  |
| Pre-eclampsia superimposed on chronic hypertension with abortive outcome |  |  |
| Pre-eclampsia superimposed on chronic hypertension, antepartum |  |  |
| Pre-eclampsia superimposed on chronic hypertension, antepartum condition or complication |  |  |
| Pre-eclampsia superimposed on chronic hypertension, delivered |  |  |
| Pre-eclampsia superimposed on chronic hypertension, delivered, with mention of postpartum complication |  |  |
| Pre-eclampsia superimposed on chronic hypertension, delivered, with or without mention of antepartum condition |  |  |
| Pre-eclampsia superimposed on chronic hypertension, delivered, with postpartum complication |  |  |
| Pre-eclampsia superimposed on chronic hypertension, postpartum |  |  |
| Pre-eclampsia superimposed on chronic hypertension, postpartum condition or complication |  |  |
| Pre-eclampsia superimposed on chronic hypertension, unspecified as to episode of care, or not applicable |  |  |
| Pre-eclampsia with abortive outcome |  |  |
| Pre-eclampsia, antepartum |  |  |
| Pre-eclampsia, antepartum, second trimester |  |  |
| Pre-eclampsia, antepartum, third trimester |  |  |
| Pre-eclampsia, antepartum, unspecified trimester |  |  |
| Pre-eclampsia, delivered |  |  |
| Pre-eclampsia, delivered with postpartum condition |  |  |
| Pre-eclampsia, delivered, with postpartum complication |  |  |
| Pre-eclampsia, mild |  |  |
| Pre-eclampsia, mild to moderate, second trimester |  |  |
| Pre-eclampsia, mild to moderate, third trimester |  |  |
| Pre-eclampsia, mild, antepartum |  |  |
| Pre-eclampsia, mild, antepartum, second trimester |  |  |
| Pre-eclampsia, mild, antepartum, third trimester |  |  |
| Pre-eclampsia, mild, antepartum, unspecified trimester |  |  |
| Pre-eclampsia, mild, delivered |  |  |
| Pre-eclampsia, mild, delivered with postpartum complication |  |  |
| Pre-eclampsia, mild, postpartum |  |  |
| Pre-eclampsia, mild, second trimester |  |  |
| Pre-eclampsia, mild, third trimester |  |  |
| Pre-eclampsia, mild, unspecified trimester |  |  |
| Pre-eclampsia, postpartum |  |  |
| Pre-eclampsia, second trimester |  |  |
| Pre-eclampsia, severe |  |  |
| Pre-eclampsia, severe, antepartum |  |  |
| Pre-eclampsia, severe, antepartum, second trimester |  |  |
| Pre-eclampsia, severe, antepartum, third trimester |  |  |
| Pre-eclampsia, severe, antepartum, unspecified trimester |  |  |
| Pre-eclampsia, severe, delivered |  |  |
| Pre-eclampsia, severe, delivered with postpartum condition |  |  |
| Pre-eclampsia, severe, postpartum condition |  |  |
| Pre-eclampsia, severe, second trimester |  |  |
| Pre-eclampsia, severe, third trimester |  |  |
| Pre-eclampsia, severe, unspecified trimester |  |  |
| Pre-eclampsia, severe, with delivery |  |  |
| Pre-eclampsia, third trimester |  |  |
| Pre-eclampsia, unspecified |  |  |
| Pre-eclampsia, unspecified trimester |  |  |
| Pre-eclampsia, unspecified, antepartum condition or complication |  |  |
| Pre-eclampsia, unspecified, delivered, with mention of postpartum complication |  |  |
| Pre-eclampsia, unspecified, delivered, with or without mention of antepartum condition |  |  |
| Pre-eclampsia, unspecified, postpartum condition or complication |  |  |
| Pre-eclampsia, unspecified, unspecified as to episode of care, or not applicable |  |  |
| Pre-eclampsia/eclampsia with prior hypertension, antepartum |  |  |
| Pre-eclampsia/eclampsia with prior hypertension, delivered |  |  |
| Pre-eclampsia/eclampsia with prior hypertension, delivered/postpartum |  |  |
| Pre-eclampsia/eclampsia with prior hypertension, postpartum condition |  |  |
| Pre-eclamptic toxemia |  |  |
| Pre-eclamptic toxemia, second trimester |  |  |
| Pre-eclamptic toxemia, third trimester |  |  |
| Pre-eclamptic toxemia, unspecified trimester |  |  |
| Pre-exist hyp heart and chr kidney dis comp preg, first tri |  |  |
| Pre-exist hyp heart and chr kidney dis comp preg, second tri |  |  |
| Pre-exist hyp heart and chr kidney dis comp preg, third tri |  |  |
| Pre-exist hyp heart and chr kidney dis comp preg, unsp tri |  |  |
| Pre-exist hyp heart and chr kidney disease comp the puerp |  |  |
| Pre-exist hyp heart and chronic kidney disease comp chldbrth |  |  |
| Pre-exist hyp heart disease comp pregnancy, first trimester |  |  |
| Pre-exist hyp heart disease comp pregnancy, second trimester |  |  |
| Pre-exist hyp heart disease comp pregnancy, third trimester |  |  |
| Pre-exist hyp heart disease comp pregnancy, unsp trimester |  |  |
| Pre-existing essential htn comp pregnancy, first trimester |  |  |
| Pre-existing essential htn comp pregnancy, second trimester |  |  |
| Pre-existing essential htn comp pregnancy, third trimester |  |  |
| Pre-existing essential htn comp pregnancy, unsp trimester |  |  |
| Pre-existing essential hypertension affecting pregnancy |  |  |
| Pre-existing essential hypertension affecting pregnancy in first trimester |  |  |
| Pre-existing essential hypertension affecting pregnancy in second trimester |  |  |
| Pre-existing essential hypertension affecting pregnancy in third trimester |  |  |
| Pre-existing essential hypertension affecting pregnancy, antepartum |  |  |
| Pre-existing essential hypertension affecting pregnancy, first trimester |  |  |
| Pre-existing essential hypertension affecting pregnancy, second trimester |  |  |
| Pre-existing essential hypertension affecting pregnancy, third trimester |  |  |
| Pre-existing essential hypertension affecting pregnancy, unspecified trimester |  |  |
| Pre-existing essential hypertension complicating childbirth |  |  |
| Pre-existing essential hypertension complicating pregnancy |  |  |
| Pre-existing essential hypertension complicating pregnancy in first trimester |  |  |
| Pre-existing essential hypertension complicating pregnancy in second trimester |  |  |
| Pre-existing essential hypertension complicating pregnancy in third trimester |  |  |
| Pre-existing essential hypertension complicating pregnancy, childbirth and the puerperium |  |  |
| Pre-existing essential hypertension complicating pregnancy, childbirth and the puerperium, antepartum condition or complication |  |  |
| Pre-existing essential hypertension complicating pregnancy, childbirth and the puerperium, delivered, with mention of postpartum complication |  |  |
| Pre-existing essential hypertension complicating pregnancy, childbirth and the puerperium, delivered, with or without mention of antepartum condition |  |  |
| Pre-existing essential hypertension complicating pregnancy, childbirth and the puerperium, postpartum condition or complication |  |  |
| Pre-existing essential hypertension complicating pregnancy, childbirth and the puerperium, unspecified as to episode of care, or not applicable |  |  |
| Pre-existing essential hypertension complicating pregnancy, first trimester |  |  |
| Pre-existing essential hypertension complicating pregnancy, second trimester |  |  |
| Pre-existing essential hypertension complicating pregnancy, third trimester |  |  |
| Pre-existing essential hypertension complicating pregnancy, unspecified trimester |  |  |
| Pre-existing essential hypertension complicating puerperium |  |  |
| Pre-existing essential hypertension complicating the puerperium |  |  |
| Pre-existing essential hypertension during childbirth |  |  |
| Pre-existing essential hypertension during pregnancy |  |  |
| Pre-existing essential hypertension during pregnancy in first trimester |  |  |
| Pre-existing essential hypertension during pregnancy in second trimester |  |  |
| Pre-existing essential hypertension during pregnancy in third trimester |  |  |
| Pre-existing essential hypertension during pregnancy, antepartum |  |  |
| Pre-existing essential hypertension during pregnancy, unspecified trimester |  |  |
| Pre-existing essential hypertension during puerperium |  |  |
| Pre-existing essential hypertension, postpartum |  |  |
| Pre-existing hyp chronic kidney disease comp childbirth |  |  |
| Pre-existing hyp chronic kidney disease comp the puerperium |  |  |
| Pre-existing hypertension affecting pregnancy |  |  |
| Pre-existing hypertension affecting pregnancy in first trimester |  |  |
| Pre-existing hypertension affecting pregnancy in first trimester, antepartum |  |  |
| Pre-existing hypertension affecting pregnancy in second trimester |  |  |
| Pre-existing hypertension affecting pregnancy in second trimester, antepartum |  |  |
| Pre-existing hypertension affecting pregnancy in third trimester |  |  |
| Pre-existing hypertension affecting pregnancy in third trimester, antepartum |  |  |
| Pre-existing hypertension affecting pregnancy, antepartum |  |  |
| Pre-existing hypertension affecting pregnancy, delivered, current hospitalization |  |  |
| Pre-existing hypertension complicating childbirth |  |  |
| Pre-existing hypertension complicating or reason for care during pregnancy |  |  |
| Pre-existing hypertension complicating or reason for care during pregnancy in first trimester |  |  |
| Pre-existing hypertension complicating or reason for care during pregnancy in second trimester |  |  |
| Pre-existing hypertension complicating or reason for care during pregnancy in third trimester |  |  |
| Pre-existing hypertension complicating pregnancy |  |  |
| Pre-existing hypertension complicating pregnancy in first trimester |  |  |
| Pre-existing hypertension complicating pregnancy in second trimester |  |  |
| Pre-existing hypertension complicating pregnancy in third trimester |  |  |
| Pre-existing hypertension complicating pregnancy, childbirth and the puerperium |  |  |
| Pre-existing hypertension complicating pregnancy, first trimester |  |  |
| Pre-existing hypertension complicating pregnancy, second trimester |  |  |
| Pre-existing hypertension complicating pregnancy, third trimester |  |  |
| Pre-existing hypertension complicating pregnancy, unspecified trimester |  |  |
| Pre-existing hypertension complicating puerperium |  |  |
| Pre-existing hypertension complicating the puerperium |  |  |
| Pre-existing hypertension during pregnancy |  |  |
| Pre-existing hypertension during pregnancy in first trimester |  |  |
| Pre-existing hypertension during pregnancy in first trimester, unspecified pre-existing hypertension |  |  |
| Pre-existing hypertension during pregnancy in first trimester, unspecified pre-existing hypertension type |  |  |
| Pre-existing hypertension during pregnancy in second trimester |  |  |
| Pre-existing hypertension during pregnancy in second trimester, unspecified pre-existing hypertension |  |  |
| Pre-existing hypertension during pregnancy in second trimester, unspecified pre-existing hypertension type |  |  |
| Pre-existing hypertension during pregnancy in third trimester |  |  |
| Pre-existing hypertension during pregnancy in third trimester, unspecified pre-existing hypertension |  |  |
| Pre-existing hypertension during pregnancy in third trimester, unspecified pre-existing hypertension type |  |  |
| Pre-existing hypertension during pregnancy with postpartum complication |  |  |
| Pre-existing hypertension during pregnancy, antepartum |  |  |
| Pre-existing hypertension during pregnancy, antepartum, unspecified pre-existing hypertension type |  |  |
| Pre-existing hypertension during pregnancy, unspecified pre-existing hypertension, unspecified trimester |  |  |
| Pre-existing hypertension during puerperium |  |  |
| Pre-existing hypertension in first trimester, antepartum |  |  |
| Pre-existing hypertension in obstetric context |  |  |
| Pre-existing hypertension in obstetric context in first trimester |  |  |
| Pre-existing hypertension in obstetric context in second trimester |  |  |
| Pre-existing hypertension in obstetric context in third trimester |  |  |
| Pre-existing hypertension in obstetric context, first trimester |  |  |
| Pre-existing hypertension in obstetric context, second trimester |  |  |
| Pre-existing hypertension in obstetric context, third trimester |  |  |
| Pre-existing hypertension in obstetric context, unspecified trimester |  |  |
| Pre-existing hypertension in pregnancy with postpartum complication |  |  |
| Pre-existing hypertension in pregnancy, childbirth and the puerperium |  |  |
| Pre-existing hypertension in pregnancy, childbirth, or puerperium |  |  |
| Pre-existing hypertension in second trimester, antepartum |  |  |
| Pre-existing hypertension in third trimester, antepartum |  |  |
| Pre-existing hypertension with pre-eclampsia in first trimester |  |  |
| Pre-existing hypertension with pre-eclampsia in second trimester |  |  |
| Pre-existing hypertension with pre-eclampsia in third trimester |  |  |
| Pre-existing hypertension with pre-eclampsia, antepartum |  |  |
| Pre-existing hypertension with pre-eclampsia, complicating childbirth |  |  |
| Pre-existing hypertension with pre-eclampsia, complicating the puerperium |  |  |
| Pre-existing hypertension with pre-eclampsia, first trimester (CODE) |  |  |
| Pre-existing hypertension with pre-eclampsia, first trimester |  |  |
| Pre-existing hypertension with pre-eclampsia, second trimester (CODE) |  |  |
| Pre-existing hypertension with pre-eclampsia, second trimester |  |  |
| Pre-existing hypertension with pre-eclampsia, third trimester (CODE) |  |  |
| Pre-existing hypertension with pre-eclampsia, third trimester |  |  |
| Pre-existing hypertension with pre-eclampsia, unspecified trimester |  |  |
| Pre-existing hypertension, antepartum |  |  |
| Pre-existing hypertension, antepartum, first trimester |  |  |
| Pre-existing hypertension, antepartum, second trimester |  |  |
| Pre-existing hypertension, antepartum, third trimester |  |  |
| Pre-existing hypertension, antepartum, unspecified trimester |  |  |
| Pre-existing hypertension, complicating or reason for care during pregnancy |  |  |
| Pre-existing hypertension, complicating or reason for care during puerperium |  |  |
| Pre-existing hypertension, with delivery |  |  |
| Pre-existing hypertension, with delivery, with current postpartum complication |  |  |
| Pre-existing hypertensive chronic kidney disease complic childbirth |  |  |
| Pre-existing hypertensive chronic kidney disease complic puerperium |  |  |
| Pre-existing hypertensive chronic kidney disease complicating childbirth (CODE) |  |  |
| Pre-existing hypertensive chronic kidney disease complicating childbirth |  |  |
| Pre-existing hypertensive chronic kidney disease complicating puerperium |  |  |
| Pre-existing hypertensive chronic kidney disease complicating the puerperium (CODE) |  |  |
| Pre-existing hypertensive chronic kidney disease complicating the puerperium |  |  |
| Pre-existing hypertensive disorder with superimposed proteinuria |  |  |
| Pre-existing hypertensive disorder with superimposed proteinuria during pregnancy |  |  |
| Pre-existing hypertensive disorder with superimposed proteinuria during pregnancy in first trimester, antepartum |  |  |
| Pre-existing hypertensive disorder with superimposed proteinuria during pregnancy in second trimester, antepartum |  |  |
| Pre-existing hypertensive disorder with superimposed proteinuria during pregnancy in third trimester, antepartum |  |  |
| Pre-existing hypertensive disorder with superimposed proteinuria during pregnancy, antepartum |  |  |
| Pre-existing hypertensive disorder with superimposed proteinuria in first trimester |  |  |
| Pre-existing hypertensive disorder with superimposed proteinuria in obstetric context |  |  |
| Pre-existing hypertensive disorder with superimposed proteinuria in obstetric context in first trimester |  |  |
| Pre-existing hypertensive disorder with superimposed proteinuria in obstetric context in second trimester |  |  |
| Pre-existing hypertensive disorder with superimposed proteinuria in obstetric context in third trimester |  |  |
| Pre-existing hypertensive disorder with superimposed proteinuria in obstetric context, antepartum |  |  |
| Pre-existing hypertensive disorder with superimposed proteinuria in obstetric context, first trimester |  |  |
| Pre-existing hypertensive disorder with superimposed proteinuria in obstetric context, second trimester |  |  |
| Pre-existing hypertensive disorder with superimposed proteinuria in obstetric context, third trimester |  |  |
| Pre-existing hypertensive disorder with superimposed proteinuria in obstetric context, unspecified trimester |  |  |
| Pre-existing hypertensive disorder with superimposed proteinuria in second trimester |  |  |
| Pre-existing hypertensive disorder with superimposed proteinuria in third trimester |  |  |
| Pre-existing hypertensive disorder with superimposed proteinuria, antepartum |  |  |
| Pre-existing hypertensive disorder with superimposed proteinuria, antepartum condition or complication |  |  |
| Pre-existing hypertensive disorder with superimposed proteinuria, antepartum condition or complication, first trimester |  |  |
| Pre-existing hypertensive disorder with superimposed proteinuria, antepartum condition or complication, second trimester |  |  |
| Pre-existing hypertensive disorder with superimposed proteinuria, antepartum condition or complication, third trimester |  |  |
| Pre-existing hypertensive disorder with superimposed proteinuria, antepartum condition or complication, unspecified trimester |  |  |
| Pre-existing hypertensive disorder with superimposed proteinuria, antepartum, first trimester |  |  |
| Pre-existing hypertensive disorder with superimposed proteinuria, antepartum, second trimester |  |  |
| Pre-existing hypertensive disorder with superimposed proteinuria, antepartum, third trimester |  |  |
| Pre-existing hypertensive disorder with superimposed proteinuria, antepartum, unspecified trimester |  |  |
| Pre-existing hypertensive disorder with superimposed proteinuria, delivered |  |  |
| Pre-existing hypertensive disorder with superimposed proteinuria, postpartum |  |  |
| Pre-existing hypertensive disorder with superimposed proteinuria, postpartum condition or complication |  |  |
| Pre-existing hypertensive heart and chronic kidney disease affecting pregnancy |  |  |
| Pre-existing hypertensive heart and chronic kidney disease affecting pregnancy in first trimester |  |  |
| Pre-existing hypertensive heart and chronic kidney disease affecting pregnancy in first trimester, antepartum |  |  |
| Pre-existing hypertensive heart and chronic kidney disease affecting pregnancy in second trimester |  |  |
| Pre-existing hypertensive heart and chronic kidney disease affecting pregnancy in second trimester, antepartum |  |  |
| Pre-existing hypertensive heart and chronic kidney disease affecting pregnancy in third trimester |  |  |
| Pre-existing hypertensive heart and chronic kidney disease affecting pregnancy in third trimester, antepartum |  |  |
| Pre-existing hypertensive heart and chronic kidney disease affecting pregnancy, antepartum |  |  |
| Pre-existing hypertensive heart and chronic kidney disease affecting pregnancy, antepartum, first trimester |  |  |
| Pre-existing hypertensive heart and chronic kidney disease affecting pregnancy, antepartum, second trimester |  |  |
| Pre-existing hypertensive heart and chronic kidney disease affecting pregnancy, antepartum, third trimester |  |  |
| Pre-existing hypertensive heart and chronic kidney disease affecting pregnancy, antepartum, unspecified trimester |  |  |
| Pre-existing hypertensive heart and chronic kidney disease affecting pregnancy, first trimester |  |  |
| Pre-existing hypertensive heart and chronic kidney disease affecting pregnancy, second trimester |  |  |
| Pre-existing hypertensive heart and chronic kidney disease affecting pregnancy, third trimester |  |  |
| Pre-existing hypertensive heart and chronic kidney disease affecting pregnancy, unspecified trimester |  |  |
| Pre-existing hypertensive heart and chronic kidney disease complicating childbirth |  |  |
| Pre-existing hypertensive heart and chronic kidney disease complicating pregnancy |  |  |
| Pre-existing hypertensive heart and chronic kidney disease complicating pregnancy in first trimester |  |  |
| Pre-existing hypertensive heart and chronic kidney disease complicating pregnancy in second trimester |  |  |
| Pre-existing hypertensive heart and chronic kidney disease complicating pregnancy in third trimester |  |  |
| Pre-existing hypertensive heart and chronic kidney disease complicating pregnancy, first trimester (CODE) |  |  |
| Pre-existing hypertensive heart and chronic kidney disease complicating pregnancy, first trimester |  |  |
| Pre-existing hypertensive heart and chronic kidney disease complicating pregnancy, second trimester (CODE) |  |  |
| Pre-existing hypertensive heart and chronic kidney disease complicating pregnancy, second trimester |  |  |
| Pre-existing hypertensive heart and chronic kidney disease complicating pregnancy, third trimester (CODE) |  |  |
| Pre-existing hypertensive heart and chronic kidney disease complicating pregnancy, third trimester |  |  |
| Pre-existing hypertensive heart and chronic kidney disease complicating pregnancy, unspecified trimester |  |  |
| Pre-existing hypertensive heart and chronic kidney disease complicating puerperium |  |  |
| Pre-existing hypertensive heart and chronic kidney disease complicating the puerperium |  |  |
| Pre-existing hypertensive heart and chronic kidney disease in mother affecting childbirth |  |  |
| Pre-existing hypertensive heart and renal disease complicating pregnancy, childbirth and the puerperium |  |  |
| Pre-existing hypertensive heart and renal disease complicating pregnancy, childbirth and the puerperium, antepartum condition or complication |  |  |
| Pre-existing hypertensive heart and renal disease complicating pregnancy, childbirth and the puerperium, delivered, with mention of postpartum complication |  |  |
| Pre-existing hypertensive heart and renal disease complicating pregnancy, childbirth and the puerperium, delivered, with or without mention of antepartum condition |  |  |
| Pre-existing hypertensive heart and renal disease complicating pregnancy, childbirth and the puerperium, postpartum condition or complication |  |  |
| Pre-existing hypertensive heart and renal disease complicating pregnancy, childbirth and the puerperium, unspecified as to episode of care, or not applicable |  |  |
| Pre-existing hypertensive heart disease affecting pregnancy |  |  |
| Pre-existing hypertensive heart disease affecting pregnancy in first trimester |  |  |
| Pre-existing hypertensive heart disease affecting pregnancy in first trimester, antepartum |  |  |
| Pre-existing hypertensive heart disease affecting pregnancy in second trimester |  |  |
| Pre-existing hypertensive heart disease affecting pregnancy in second trimester, antepartum |  |  |
| Pre-existing hypertensive heart disease affecting pregnancy in third trimester |  |  |
| Pre-existing hypertensive heart disease affecting pregnancy in third trimester, antepartum |  |  |
| Pre-existing hypertensive heart disease affecting pregnancy, antepartum |  |  |
| Pre-existing hypertensive heart disease affecting pregnancy, antepartum, first trimester |  |  |
| Pre-existing hypertensive heart disease affecting pregnancy, antepartum, second trimester |  |  |
| Pre-existing hypertensive heart disease affecting pregnancy, antepartum, third trimester |  |  |
| Pre-existing hypertensive heart disease affecting pregnancy, antepartum, unspecified trimester |  |  |
| Pre-existing hypertensive heart disease affecting pregnancy, first trimester |  |  |
| Pre-existing hypertensive heart disease affecting pregnancy, second trimester |  |  |
| Pre-existing hypertensive heart disease affecting pregnancy, third trimester |  |  |
| Pre-existing hypertensive heart disease affecting pregnancy, unspecified trimester |  |  |
| Pre-existing hypertensive heart disease complicating childbirth (CODE) |  |  |
| Pre-existing hypertensive heart disease complicating childbirth |  |  |
| Pre-existing hypertensive heart disease complicating pregnancy |  |  |
| Pre-existing hypertensive heart disease complicating pregnancy in first trimester |  |  |
| Pre-existing hypertensive heart disease complicating pregnancy in second trimester |  |  |
| Pre-existing hypertensive heart disease complicating pregnancy in third trimester |  |  |
| Pre-existing hypertensive heart disease complicating pregnancy, childbirth and the puerperium |  |  |
| Pre-existing hypertensive heart disease complicating pregnancy, childbirth and the puerperium, antepartum condition or complication |  |  |
| Pre-existing hypertensive heart disease complicating pregnancy, childbirth and the puerperium, delivered, with mention of postpartum complication |  |  |
| Pre-existing hypertensive heart disease complicating pregnancy, childbirth and the puerperium, delivered, with or without mention of antepartum condition |  |  |
| Pre-existing hypertensive heart disease complicating pregnancy, childbirth and the puerperium, postpartum condition or complication |  |  |
| Pre-existing hypertensive heart disease complicating pregnancy, childbirth and the puerperium, unspecified as to episode of care, or not applicable |  |  |
| Pre-existing hypertensive heart disease complicating pregnancy, first trimester (CODE) |  |  |
| Pre-existing hypertensive heart disease complicating pregnancy, first trimester |  |  |
| Pre-existing hypertensive heart disease complicating pregnancy, second trimester (CODE) |  |  |
| Pre-existing hypertensive heart disease complicating pregnancy, second trimester |  |  |
| Pre-existing hypertensive heart disease complicating pregnancy, third trimester (CODE) |  |  |
| Pre-existing hypertensive heart disease complicating pregnancy, third trimester |  |  |
| Pre-existing hypertensive heart disease complicating pregnancy, unspecified trimester (CODE) |  |  |
| Pre-existing hypertensive heart disease complicating pregnancy, unspecified trimester |  |  |
| Pre-existing hypertensive heart disease complicating puerperium |  |  |
| Pre-existing hypertensive heart disease complicating the puerperium (CODE) |  |  |
| Pre-existing hypertensive heart disease complicating the puerperium |  |  |
| Pre-existing hypertensive heart disease in mother affecting childbirth |  |  |
| Pre-existing hypertensive kidney disease affecting pregnancy, antepartum |  |  |
| Pre-existing hypertensive renal disease complicating pregnancy, childbirth and the puerperium |  |  |
| Pre-existing hypertensive renal disease complicating pregnancy, childbirth and the puerperium, antepartum condition or complication |  |  |
| Pre-existing hypertensive renal disease complicating pregnancy, childbirth and the puerperium, delivered, with mention of postpartum complication |  |  |
| Pre-existing hypertensive renal disease complicating pregnancy, childbirth and the puerperium, delivered, with or without mention of antepartum condition |  |  |
| Pre-existing hypertensive renal disease complicating pregnancy, childbirth and the puerperium, postpartum condition or complication |  |  |
| Pre-existing hypertensive renal disease complicating pregnancy, childbirth and the puerperium, unspecified as to episode of care, or not applicable |  |  |
| Pre-existing secondary htn comp pregnancy, first trimester |  |  |
| Pre-existing secondary htn comp pregnancy, second trimester |  |  |
| Pre-existing secondary htn comp pregnancy, third trimester |  |  |
| Pre-existing secondary htn comp pregnancy, unsp trimester |  |  |
| Pre-existing secondary hypertension affecting pregnancy |  |  |
| Pre-existing secondary hypertension affecting pregnancy in first trimester |  |  |
| Pre-existing secondary hypertension affecting pregnancy in first trimester, antepartum |  |  |
| Pre-existing secondary hypertension affecting pregnancy in second trimester |  |  |
| Pre-existing secondary hypertension affecting pregnancy in second trimester, antepartum |  |  |
| Pre-existing secondary hypertension affecting pregnancy in third trimester |  |  |
| Pre-existing secondary hypertension affecting pregnancy in third trimester, antepartum |  |  |
| Pre-existing secondary hypertension affecting pregnancy, antepartum |  |  |
| Pre-existing secondary hypertension affecting pregnancy, antepartum, first trimester |  |  |
| Pre-existing secondary hypertension affecting pregnancy, antepartum, second trimester |  |  |
| Pre-existing secondary hypertension affecting pregnancy, antepartum, third trimester |  |  |
| Pre-existing secondary hypertension affecting pregnancy, antepartum, unspecified trimester |  |  |
| Pre-existing secondary hypertension affecting pregnancy, first trimester |  |  |
| Pre-existing secondary hypertension affecting pregnancy, second trimester |  |  |
| Pre-existing secondary hypertension affecting pregnancy, third trimester |  |  |
| Pre-existing secondary hypertension affecting pregnancy, unspecified trimester |  |  |
| Pre-existing secondary hypertension complicating childbirth (CODE) |  |  |
| Pre-existing secondary hypertension complicating childbirth |  |  |
| Pre-existing secondary hypertension complicating pregnancy |  |  |
| Pre-existing secondary hypertension complicating pregnancy in first trimester |  |  |
| Pre-existing secondary hypertension complicating pregnancy in second trimester |  |  |
| Pre-existing secondary hypertension complicating pregnancy in third trimester |  |  |
| Pre-existing secondary hypertension complicating pregnancy, childbirth and the puerperium |  |  |
| Pre-existing secondary hypertension complicating pregnancy, childbirth and the puerperium, antepartum condition or complication |  |  |
| Pre-existing secondary hypertension complicating pregnancy, childbirth and the puerperium, delivered, with mention of postpartum complication |  |  |
| Pre-existing secondary hypertension complicating pregnancy, childbirth and the puerperium, delivered, with or without mention of antepartum condition |  |  |
| Pre-existing secondary hypertension complicating pregnancy, childbirth and the puerperium, postpartum condition or complication |  |  |
| Pre-existing secondary hypertension complicating pregnancy, childbirth and the puerperium, unspecified as to episode of care, or not applicable |  |  |
| Pre-existing secondary hypertension complicating pregnancy, first trimester |  |  |
| Pre-existing secondary hypertension complicating pregnancy, second trimester |  |  |
| Pre-existing secondary hypertension complicating pregnancy, third trimester (CODE) |  |  |
| Pre-existing secondary hypertension complicating pregnancy, third trimester |  |  |
| Pre-existing secondary hypertension complicating pregnancy, unspecified trimester (CODE) |  |  |
| Pre-existing secondary hypertension complicating pregnancy, unspecified trimester |  |  |
| Pre-existing secondary hypertension complicating puerperium |  |  |
| Pre-existing secondary hypertension complicating the puerperium (CODE) |  |  |
| Pre-existing secondary hypertension complicating the puerperium |  |  |
| Pre-existing secondary hypertension during childbirth |  |  |
| Pre-existing secondary hypertension during pregnancy |  |  |
| Pre-existing secondary hypertension during pregnancy in first trimester |  |  |
| Pre-existing secondary hypertension during pregnancy in second trimester |  |  |
| Pre-existing secondary hypertension during pregnancy in third trimester |  |  |
| Pre-existing secondary hypertension during pregnancy, antepartum |  |  |
| Pre-existing secondary hypertension during pregnancy, unspecified trimester |  |  |
| Pre-existing secondary hypertension during puerperium |  |  |
| Pre-transplant hypertension |  |  |
| Preeclam superimp chronic h/t/antepartum |  |  |
| Preeclam superimp chronic h/t/postpartum |  |  |
| Preeclam superimp chronic h/tdel w compl |  |  |
| Preeclam superimp chronic hypert/unspec |  |  |
| Preeclamp superimp on chronic hypert/del |  |  |
| Preeclampsia |  |  |
| Preeclampsia complicating hypertension |  |  |
| Preeclampsia in postpartum period |  |  |
| Preeclampsia in postpartum period, second trimester |  |  |
| Preeclampsia in postpartum period, third trimester |  |  |
| Preeclampsia in postpartum period, unspecified trimester |  |  |
| Preeclampsia unspec delivered |  |  |
| Preeclampsia unspec unspec epis care |  |  |
| Preeclampsia unspeci del w comp |  |  |
| Preeclampsia, second trimester |  |  |
| Preeclampsia, severe |  |  |
| Preeclampsia, severe, second trimester |  |  |
| Preeclampsia, severe, third trimester |  |  |
| Preeclampsia, severe, unspecified trimester |  |  |
| Preeclampsia, third trimester |  |  |
| Preeclampsia, unspecified trimester |  |  |
| Preexisting hypertension complic pregnan, with deliv, curr hospitaliz |  |  |
| Preexisting hypertension complicating pregnancy in first trimester, antepartum |  |  |
| Preexisting hypertension complicating pregnancy in second trimester, antepartum |  |  |
| Preexisting hypertension complicating pregnancy in third trimester, antepartum |  |  |
| Preexisting hypertension complicating pregnancy, antepartum |  |  |
| Preexisting hypertension complicating pregnancy, antepartum, first trimester |  |  |
| Preexisting hypertension complicating pregnancy, antepartum, second trimester |  |  |
| Preexisting hypertension complicating pregnancy, antepartum, third trimester |  |  |
| Preexisting hypertension complicating pregnancy, antepartum, unspecified trimester |  |  |
| Preexisting hypertension complicating pregnancy, with delivery, current hospitalization |  |  |
| Pregnancy induced hypertension |  |  |
| Pregnancy induced hypertension, antepartum |  |  |
| Pregnancy induced hypertension, delivered, current hospitalization |  |  |
| Pregnancy induced hypertension, delivered, with postpartum complication |  |  |
| Pregnancy induced hypertension, first trimester |  |  |
| Pregnancy induced hypertension, postpartum |  |  |
| Pregnancy induced hypertension, second trimester |  |  |
| Pregnancy induced hypertension, third trimester |  |  |
| Pregnancy induced hypertension, unspecified trimester |  |  |
| Pregnancy toxemia |  |  |
| Pregnancy toxemia /hypertension |  |  |
| Pregnancy toxemia /hypertension, second trimester |  |  |
| Pregnancy toxemia /hypertension, third trimester |  |  |
| Pregnancy toxemia /hypertension, unspecified trimester |  |  |
| Pregnancy toxemia, second trimester |  |  |
| Pregnancy toxemia, third trimester |  |  |
| Pregnancy toxemia, unspecified trimester |  |  |
| Pregnancy with pre-existing hypertension |  |  |
| Pregnancy-induced benign hypertension |  |  |
| Pregnancy-induced benign hypertension in first trimester |  |  |
| Pregnancy-induced benign hypertension in second trimester |  |  |
| Pregnancy-induced benign hypertension in third trimester |  |  |
| Pregnancy-induced benign hypertension, antepartum |  |  |
| Pregnancy-induced benign hypertension, first trimester |  |  |
| Pregnancy-induced benign hypertension, second trimester |  |  |
| Pregnancy-induced benign hypertension, third trimester |  |  |
| Pregnancy-induced benign hypertension, unspecified trimester |  |  |
| Pregnancy-induced hypertension |  |  |
| Pregnancy-induced hypertension in first trimester |  |  |
| Pregnancy-induced hypertension in second trimester |  |  |
| Pregnancy-induced hypertension in third trimester |  |  |
| Pregnancy-induced hypertension with pulmonary edema |  |  |
| Pregnancy-induced hypertension, delivered |  |  |
| Pregnancy-induced hypertension, first trimester |  |  |
| Pregnancy-induced hypertension, second trimester |  |  |
| Pregnancy-induced hypertension, third trimester |  |  |
| Pregnancy-induced hypertension, unspecified trimester |  |  |
| Pregnancy-induced hypertensive state |  |  |
| Pregnancy-induced hypertensive state, first trimester |  |  |
| Pregnancy-induced hypertensive state, second trimester |  |  |
| Pregnancy-induced hypertensive state, third trimester |  |  |
| Pregnancy-induced hypertensive state, unspecified trimester |  |  |
| Primary hypertension |  |  |
| Proteinuric hypertension of pregnancy |  |  |
| Proteinuric hypertension of pregnancy in second trimester |  |  |
| Proteinuric hypertension of pregnancy in second trimester, antepartum |  |  |
| Proteinuric hypertension of pregnancy in third trimester |  |  |
| Proteinuric hypertension of pregnancy in third trimester, antepartum |  |  |
| Proteinuric hypertension of pregnancy, antepartum |  |  |
| Proteinuric hypertension of pregnancy, antepartum, second trimester |  |  |
| Proteinuric hypertension of pregnancy, antepartum, third trimester |  |  |
| Proteinuric hypertension of pregnancy, antepartum, unspecified trimester |  |  |
| Proteinuric hypertension of pregnancy, second trimester |  |  |
| Proteinuric hypertension of pregnancy, third trimester |  |  |
| Proteinuric hypertension of pregnancy, unspecified trimester |  |  |
| PTGIS-related essential hypertension |  |  |
| Reactive hypertension |  |  |
| Rebound hypertension |  |  |
| Renal arterial hypertension |  |  |
| Renal HTN-del w/postpartum complic |  |  |
| Renal hyperten-postpart |  |  |
| Renal hypertension |  |  |
| Renal hypertension affecting pregnancy |  |  |
| Renal hypertension affecting pregnancy in first trimester |  |  |
| Renal hypertension affecting pregnancy in first trimester, antepartum |  |  |
| Renal hypertension affecting pregnancy in second trimester |  |  |
| Renal hypertension affecting pregnancy in second trimester, antepartum |  |  |
| Renal hypertension affecting pregnancy in third trimester |  |  |
| Renal hypertension affecting pregnancy in third trimester, antepartum |  |  |
| Renal hypertension affecting pregnancy, antepartum |  |  |
| Renal hypertension affecting pregnancy, antepartum, first trimester |  |  |
| Renal hypertension affecting pregnancy, antepartum, second trimester |  |  |
| Renal hypertension affecting pregnancy, antepartum, third trimester |  |  |
| Renal hypertension affecting pregnancy, antepartum, unspecified trimester |  |  |
| Renal hypertension complicating pregnancy |  |  |
| Renal hypertension complicating pregnancy in first trimester |  |  |
| Renal hypertension complicating pregnancy in second trimester |  |  |
| Renal hypertension complicating pregnancy in third trimester |  |  |
| Renal hypertension complicating pregnancy, childbirth and puerperium with postnatal complication |  |  |
| Renal hypertension complicating pregnancy, first trimester |  |  |
| Renal hypertension complicating pregnancy, second trimester |  |  |
| Renal hypertension complicating pregnancy, third trimester |  |  |
| Renal hypertension complicating pregnancy, unspecified trimester |  |  |
| Renal hypertension during postpartum period |  |  |
| Renal hypertension during pregnancy, delivered |  |  |
| Renal hypertension in pregnancy, delivered |  |  |
| Renal hypertension, delivered, with postpartum complication |  |  |
| Renal hypertension, non-vascular |  |  |
| Renal hypertension, stage 5 chronic kidney disease or end stage renal disease |  |  |
| Renal sclerosis with hypertension |  |  |
| Renal sclerosis with hypertension, stage 1 through stage 4 or unspecified chronic kidney disease |  |  |
| Renal sclerosis with hypertension, stage 1-4 or unspecified chronic kidney disease |  |  |
| Renal sclerosis with hypertension, stage 5 chronic kidney disease or end stage renal disease |  |  |
| Renovascular hypertension |  |  |
| Renovascular hypertension in child |  |  |
| Renovascular hypertension in pediatric patient |  |  |
| Renovascular hypertension with goal blood pressure less than 130/80 |  |  |
| Renovascular hypertension with goal blood pressure less than 130/85 |  |  |
| Renovascular hypertension with goal blood pressure less than 140/90 |  |  |
| Renovascular hypertension with goal blood pressure less than 150/90 |  |  |
| Renovascular hypertension with unknown severity |  |  |
| Renovascular hypertension, benign or unspecified |  |  |
| Renovascular hypertension, malignant |  |  |
| Resistant hypertension |  |  |
| Secondary diabetes mellitus with hypertension and end stage renal disease on dialysis |  |  |
| Secondary diabetes mellitus with stage 1 chronic kidney disease and hypertension |  |  |
| Secondary diabetes mellitus with stage 2 chronic kidney disease and hypertension |  |  |
| Secondary diabetes mellitus with stage 3 chronic kidney disease and hypertension |  |  |
| Secondary diabetes mellitus with stage 4 chronic kidney disease and hypertension |  |  |
| Secondary diabetes mellitus with stage 5 chronic kidney disease and hypertension |  |  |
| Secondary diastolic hypertension |  |  |
| Secondary DM with CKD stage 1 and hypertension |  |  |
| Secondary DM with CKD stage 2 and hypertension |  |  |
| Secondary DM with CKD stage 3 and hypertension |  |  |
| Secondary DM with CKD stage 4 and hypertension |  |  |
| Secondary DM with CKD stage 5 and hypertension |  |  |
| Secondary DM with hypertension and ESRD on dialysis |  |  |
| Secondary hypertension |  |  |
| Secondary hypertension due to congenital heart disease |  |  |
| Secondary hypertension due to glomerulosclerosis |  |  |
| Secondary hypertension due to glomerulosclerosis, stage 1 through stage 4 or unspecified chronic kidney disease |  |  |
| Secondary hypertension due to glomerulosclerosis, stage 1-4 or unspecified chronic kidney disease |  |  |
| Secondary hypertension due to glomerulosclerosis, stage 5 chronic kidney disease or end stage renal disease |  |  |
| Secondary hypertension due to polycystic kidney |  |  |
| Secondary hypertension due to renal disease |  |  |
| Secondary hypertension due to renal tubular disorder |  |  |
| Secondary hypertension in child |  |  |
| Secondary hypertension in pediatric patient |  |  |
| Secondary hypertension with goal blood pressure less than 130/80 |  |  |
| Secondary hypertension with goal blood pressure less than 130/85 |  |  |
| Secondary hypertension with goal blood pressure less than 140/90 |  |  |
| Secondary hypertension with goal blood pressure less than 150/90 |  |  |
| Secondary hypertension, benign |  |  |
| Secondary hypertension, benign or unspecified |  |  |
| Secondary hypertension, malignant |  |  |
| Secondary hypertension, unspecified |  |  |
| Secondary hypertension, unspecified, malignant |  |  |
| Secondary hypertensive syndrome |  |  |
| Secondary malignant non-renal hypertension |  |  |
| Secondary renovascular hypertension, benign |  |  |
| Secondary renovascular hypertension, malignant |  |  |
| Secondary renovascular hypertension, unspecified |  |  |
| Sev preeclamp-del w/ postpartum complication |  |  |
| Severe diastolic hypertension |  |  |
| Severe essential hypertension |  |  |
| Severe hypertension |  |  |
| Severe hypertension affecting pregnancy |  |  |
| Severe hypertension affecting pregnancy in first trimester |  |  |
| Severe hypertension affecting pregnancy in first trimester, antepartum |  |  |
| Severe hypertension affecting pregnancy in second trimester |  |  |
| Severe hypertension affecting pregnancy in second trimester, antepartum |  |  |
| Severe hypertension affecting pregnancy in third trimester |  |  |
| Severe hypertension affecting pregnancy in third trimester, antepartum |  |  |
| Severe hypertension affecting pregnancy, antepartum |  |  |
| Severe hypertension affecting pregnancy, antepartum, first trimester |  |  |
| Severe hypertension affecting pregnancy, antepartum, second trimester |  |  |
| Severe hypertension affecting pregnancy, antepartum, third trimester |  |  |
| Severe hypertension affecting pregnancy, antepartum, unspecified trimester |  |  |
| Severe hypertension by National Institute for Health and Clinical Excellence 2011 criteria |  |  |
| Severe hypertension complicating childbirth |  |  |
| Severe hypertension complicating pregnancy |  |  |
| Severe hypertension complicating pregnancy, first trimester |  |  |
| Severe hypertension complicating pregnancy, second trimester |  |  |
| Severe hypertension complicating pregnancy, third trimester |  |  |
| Severe hypertension complicating pregnancy, unspecified trimester |  |  |
| Severe pre-eclampsia |  |  |
| Severe pre-eclampsia affecting 10th pregnancy |  |  |
| Severe pre-eclampsia affecting childbirth |  |  |
| Severe pre-eclampsia affecting eighth pregnancy |  |  |
| Severe pre-eclampsia affecting fifth pregnancy |  |  |
| Severe pre-eclampsia affecting first pregnancy |  |  |
| Severe pre-eclampsia affecting fourth pregnancy |  |  |
| Severe pre-eclampsia affecting ninth pregnancy |  |  |
| Severe pre-eclampsia affecting puerperium |  |  |
| Severe pre-eclampsia affecting second pregnancy |  |  |
| Severe pre-eclampsia affecting seventh pregnancy |  |  |
| Severe pre-eclampsia affecting sixth pregnancy |  |  |
| Severe pre-eclampsia affecting third pregnancy |  |  |
| Severe pre-eclampsia complicating childbirth |  |  |
| Severe pre-eclampsia in second trimester |  |  |
| Severe pre-eclampsia in second trimester, antepartum |  |  |
| Severe pre-eclampsia in third trimester |  |  |
| Severe pre-eclampsia in third trimester, antepartum |  |  |
| Severe pre-eclampsia with abortive outcome |  |  |
| Severe pre-eclampsia with postnatal complication |  |  |
| Severe pre-eclampsia, antepartum |  |  |
| Severe pre-eclampsia, antepartum condition or complication |  |  |
| Severe pre-eclampsia, antepartum, second trimester |  |  |
| Severe pre-eclampsia, antepartum, third trimester |  |  |
| Severe pre-eclampsia, antepartum, unspecified trimester |  |  |
| Severe pre-eclampsia, complicating the puerperium |  |  |
| Severe pre-eclampsia, delivered, current hospitalization |  |  |
| Severe pre-eclampsia, delivered, with mention of postpartum complication |  |  |
| Severe pre-eclampsia, delivered, with or without mention of antepartum condition |  |  |
| Severe pre-eclampsia, delivered, with postpartum complication |  |  |
| Severe pre-eclampsia, postpartum |  |  |
| Severe pre-eclampsia, postpartum condition or complication |  |  |
| Severe pre-eclampsia, second trimester |  |  |
| Severe pre-eclampsia, third trimester |  |  |
| Severe pre-eclampsia, unspecified as to episode of care |  |  |
| Severe pre-eclampsia, unspecified as to episode of care, or not applicable |  |  |
| Severe pre-eclampsia, unspecified trimester |  |  |
| Severe pre-eclampsia, with delivery |  |  |
| Severe pre-eclampsia, with delivery, with current postpartum complication |  |  |
| Severe pre-eclampsia, with delivery, with mention of postpartum complication |  |  |
| Severe pre-eclamptic toxemia |  |  |
| Severe pre-eclamptic toxemia, second trimester |  |  |
| Severe pre-eclamptic toxemia, third trimester |  |  |
| Severe pre-eclamptic toxemia, unspecified trimester |  |  |
| Severe preeclampsia |  |  |
| Severe preeclampsia del w comp |  |  |
| Severe preeclampsia delivered |  |  |
| Severe preeclampsia unspecified |  |  |
| Severe preeclampsia, second trimester |  |  |
| Severe preeclampsia, third trimester |  |  |
| Severe preeclampsia, unspecified trimester |  |  |
| Severe toxemia of pregnancy |  |  |
| Severe toxemia of pregnancy, second trimester |  |  |
| Severe toxemia of pregnancy, third trimester |  |  |
| Severe toxemia of pregnancy, unspecified trimester |  |  |
| Severe uncontrolled hypertension |  |  |
| Severely increased blood pressure and edema during pregnancy |  |  |
| Severely increased blood pressure and edema during pregnancy in second trimester |  |  |
| Severely increased blood pressure and edema during pregnancy in third trimester |  |  |
| Severely increased blood pressure and edema during pregnancy, antepartum |  |  |
| Severely increased blood pressure and edema during pregnancy, second trimester |  |  |
| Severely increased blood pressure and edema during pregnancy, third trimester |  |  |
| Severely increased blood pressure and edema during pregnancy, unspecified trimester |  |  |
| Severely increased blood pressure and swelling during pregnancy |  |  |
| Severely increased blood pressure and swelling during pregnancy in second trimester |  |  |
| Severely increased blood pressure and swelling during pregnancy in third trimester |  |  |
| Severely increased blood pressure and swelling during pregnancy, antepartum |  |  |
| Severely increased blood pressure and swelling during pregnancy, second trimester |  |  |
| Severely increased blood pressure and swelling during pregnancy, third trimester |  |  |
| Severely increased blood pressure and swelling during pregnancy, unspecified trimester |  |  |
| Situational hypertension |  |  |
| Spont in utero intraparenchymal hemorrhage assoc w/ hypertension |  |  |
| Spont in utero subarachnoid hemorrhage assoc w/ hypertension |  |  |
| Spont perinatal intraparenchymal hemorrhage assoc w/ hypertension |  |  |
| Spont perinatal subarachnoid hemorrhage assoc w/ hypertension |  |  |
| Spont presum perinat intraparenchymal hemorrhage assoc w/ hypertension |  |  |
| Spont presumed perinatal intraventricular hemorrhage assoc w/ hypertension |  |  |
| Spont presumed perinatal subarachnoid hemorrhage assoc w/ hypertension |  |  |
| Spontan presumed perinatal intraventric bleed assoc with hypertension |  |  |
| Spontaneous in utero intraparenchymal intracranial hemorrhage associated with hypertension |  |  |
| Spontaneous in utero intraventricular hemorrhage assoc w/ hypertension |  |  |
| Spontaneous in utero intraventricular hemorrhage associated with hypertension |  |  |
| Spontaneous in utero subarachnoid intracranial hemorrhage associated with hypertension |  |  |
| Spontaneous perinatal intraparenchymal intracranial hemorrhage associated with hypertension |  |  |
| Spontaneous perinatal intraventricular hemorrhage assoc/ hypertension |  |  |
| Spontaneous perinatal intraventricular hemorrhage associated with hypertension |  |  |
| Spontaneous perinatal subarachnoid intracranial hemorrhage associated with hypertension |  |  |
| Spontaneous presumed perinatal intraparenchymal intracranial hemorrhage associated with hypertension |  |  |
| Spontaneous presumed perinatal intraventricular hemorrhage associated with hypertension |  |  |
| Spontaneous presumed perinatal subarachnoid intracranial hemorrhage associated with hypertension |  |  |
| Stage 1 hypertension by National Institute for Health and Clinical Excellence 2011 criteria |  |  |
| Stage 1 hypertension by National Institute for Health and Clinical Excellence 2011 criteria with evidence of end organ damage |  |  |
| Stage 1 hypertension by National Institute for Health and Clinical Excellence 2011 criteria without evidence of end organ damage |  |  |
| Stage 2 hypertension by National Institute for Health and Clinical Excellence 2011 criteria |  |  |
| Supine hypertension |  |  |
| Supplementary code for hypertension |  |  |
| Sustained diastolic hypertension |  |  |
| Sys hypertension |  |  |
| Systemic hypertension |  |  |
| Systemic hypertension due to high intracranial pressure |  |  |
| Systemic primary arterial hypertension |  |  |
| Systolic and diastolic hypertension |  |  |
| Systolic essential hypertension |  |  |
| Systolic hypertension |  |  |
| Systolic hypertension in child |  |  |
| Systolic hypertension in pediatric patient |  |  |
| Systolic hypertension with cerebrovascular disease |  |  |
| Systolic hypertension, isolated |  |  |
| Systolic secondary hypertension |  |  |
| Tacrolimus-induced hypertension |  |  |
| Temporary high blood pressure |  |  |
| Toxemia in pregnancy |  |  |
| Toxemia in pregnancy, second trimester |  |  |
| Toxemia in pregnancy, third trimester |  |  |
| Toxemia in pregnancy, unspecified trimester |  |  |
| Toxemia of pregnancy |  |  |
| Toxemia of pregnancy in second trimester |  |  |
| Toxemia of pregnancy in second trimester, antepartum |  |  |
| Toxemia of pregnancy in third trimester |  |  |
| Toxemia of pregnancy in third trimester, antepartum |  |  |
| Toxemia of pregnancy, antepartum |  |  |
| Toxemia of pregnancy, antepartum, second trimester |  |  |
| Toxemia of pregnancy, antepartum, third trimester |  |  |
| Toxemia of pregnancy, antepartum, unspecified trimester |  |  |
| Toxemia of pregnancy, second trimester |  |  |
| Toxemia of pregnancy, third trimester |  |  |
| Toxemia of pregnancy, unspecified trimester |  |  |
| Trans hyperten-del w/ postpartum complication |  |  |
| Trans hyperten-postpart |  |  |
| Transient hypertension |  |  |
| Transient hypertension during pregnancy, postpartum |  |  |
| Transient hypertension in pregnancy, postpartum |  |  |
| Transient hypertension of pregnancy |  |  |
| Transient hypertension of pregnancy in first trimester |  |  |
| Transient hypertension of pregnancy in second trimester |  |  |
| Transient hypertension of pregnancy in third trimester |  |  |
| Transient hypertension of pregnancy with postnatal complication |  |  |
| Transient hypertension of pregnancy, antepartum |  |  |
| Transient hypertension of pregnancy, antepartum, first trimester |  |  |
| Transient hypertension of pregnancy, antepartum, second trimester |  |  |
| Transient hypertension of pregnancy, antepartum, third trimester |  |  |
| Transient hypertension of pregnancy, antepartum, unspecified trimester |  |  |
| Transient hypertension of pregnancy, first trimester |  |  |
| Transient hypertension of pregnancy, postpartum |  |  |
| Transient hypertension of pregnancy, postpartum condition or complication |  |  |
| Transient hypertension of pregnancy, second trimester |  |  |
| Transient hypertension of pregnancy, third trimester |  |  |
| Transient hypertension of pregnancy, unspecified as to episode of care |  |  |
| Transient hypertension of pregnancy, unspecified trimester |  |  |
| Transient hypertension of pregnancy, with delivery |  |  |
| Transient hypertension of pregnancy, with delivery, with current postpartum complication |  |  |
| Transient hypertension of pregnancy, with delivery, with mention of postpartum complication |  |  |
| Type 1 diabetes mellitus with hypertension and end stage renal disease on dialysis |  |  |
| Type 1 diabetes mellitus with stage 1 chronic kidney disease and hypertension |  |  |
| Type 1 diabetes mellitus with stage 2 chronic kidney disease and hypertension |  |  |
| Type 1 diabetes mellitus with stage 3 chronic kidney disease and hypertension |  |  |
| Type 1 diabetes mellitus with stage 4 chronic kidney disease and hypertension |  |  |
| Type 1 diabetes mellitus with stage 5 chronic kidney disease and hypertension |  |  |
| Type 1 DM with CKD stage 1 and hypertension |  |  |
| Type 1 DM with CKD stage 2 and hypertension |  |  |
| Type 1 DM with CKD stage 3 and hypertension |  |  |
| Type 1 DM with CKD stage 4 and hypertension |  |  |
| Type 1 DM with CKD stage 5 and hypertension |  |  |
| Type 1 DM with hypertension and ESRD on dialysis |  |  |
| Type 2 diabetes mellitus with hypertension and end stage renal disease not on dialysis |  |  |
| Type 2 diabetes mellitus with hypertension and end stage renal disease on dialysis |  |  |
| Type 2 diabetes mellitus with stage 1 chronic kidney disease and hypertension |  |  |
| Type 2 diabetes mellitus with stage 2 chronic kidney disease and hypertension |  |  |
| Type 2 diabetes mellitus with stage 3 chronic kidney disease and hypertension |  |  |
| Type 2 diabetes mellitus with stage 3a chronic kidney disease and hypertension |  |  |
| Type 2 diabetes mellitus with stage 3b chronic kidney disease and hypertension |  |  |
| Type 2 diabetes mellitus with stage 4 chronic kidney disease and hypertension |  |  |
| Type 2 diabetes mellitus with stage 5 chronic kidney disease and hypertension |  |  |
| Type 2 DM with CKD stage 1 and hypertension |  |  |
| Type 2 DM with CKD stage 2 and hypertension |  |  |
| Type 2 DM with CKD stage 3 and hypertension |  |  |
| Type 2 DM with CKD stage 4 and hypertension |  |  |
| Type 2 DM with CKD stage 5 and hypertension |  |  |
| Type 2 DM with hypertension and ESRD on dialysis |  |  |
| Uncomplicated hypertension |  |  |
| Uncontrolled hypertension |  |  |
| Uncontrolled hypertension as indication for native nephrectomy |  |  |
| Uncontrolled hypertension, stage 1 |  |  |
| Uncontrolled stage 2 hypertension |  |  |
| Unsp pre-existing htn comp pregnancy, first trimester |  |  |
| Unsp pre-existing htn comp pregnancy, second trimester |  |  |
| Unsp pre-existing htn comp pregnancy, third trimester |  |  |
| Unsp pre-existing htn comp pregnancy, unsp trimester |  |  |
| Unspec maternal hypertension/delivered |  |  |
| Unspecified essential hypertension |  |  |
| Unspecified hypertension antepartum |  |  |
| Unspecified hypertension complicating pregnancy, childbirth, or the puerperium |  |  |
| Unspecified hypertension complicating pregnancy, childbirth, or the puerperium, unspecified as to episode of care |  |  |
| Unspecified hypertension, postpartum condition or complication |  |  |
| Unspecified hypertension, with delivery |  |  |
| Unspecified hypertension, with delivery, with current postpartum complication |  |  |
| Unspecified maternal hypertension |  |  |
| Unspecified maternal hypertension, antepartum condition or complication |  |  |
| Unspecified maternal hypertension, complicating childbirth |  |  |
| Unspecified maternal hypertension, complicating the puerperium |  |  |
| Unspecified maternal hypertension, delivered, with mention of postpartum complication |  |  |
| Unspecified maternal hypertension, delivered, with or without mention of antepartum condition |  |  |
| Unspecified maternal hypertension, first trimester |  |  |
| Unspecified maternal hypertension, postpartum condition or complication |  |  |
| Unspecified maternal hypertension, second trimester |  |  |
| Unspecified maternal hypertension, third trimester |  |  |
| Unspecified maternal hypertension, unspecified as to episode of care, or not applicable |  |  |
| Unspecified maternal hypertension, unspecified trimester |  |  |
| Unspecified pre-eclampsia, complicating childbirth |  |  |
| Unspecified pre-eclampsia, complicating the puerperium |  |  |
| Unspecified pre-eclampsia, second trimester |  |  |
| Unspecified pre-eclampsia, third trimester |  |  |
| Unspecified pre-eclampsia, unspecified trimester |  |  |
| Unspecified pre-existing hypertension complicating childbirth |  |  |
| Unspecified pre-existing hypertension complicating pregnancy, childbirth and the puerperium |  |  |
| Unspecified pre-existing hypertension complicating pregnancy, childbirth and the puerperium, antepartum condition or complication |  |  |
| Unspecified pre-existing hypertension complicating pregnancy, childbirth and the puerperium, delivered, with mention of postpartum complication |  |  |
| Unspecified pre-existing hypertension complicating pregnancy, childbirth and the puerperium, delivered, with or without mention of antepartum condition |  |  |
| Unspecified pre-existing hypertension complicating pregnancy, childbirth and the puerperium, postpartum condition or complication |  |  |
| Unspecified pre-existing hypertension complicating pregnancy, childbirth and the puerperium, unspecified as to episode of care, or not applicable |  |  |
| Unspecified pre-existing hypertension complicating pregnancy, first trimester |  |  |
| Unspecified pre-existing hypertension complicating pregnancy, second trimester |  |  |
| Unspecified pre-existing hypertension complicating pregnancy, third trimester |  |  |
| Unspecified pre-existing hypertension complicating pregnancy, unspecified trimester |  |  |
| Unspecified pre-existing hypertension complicating the puerperium |  |  |
| Unspecified secondary hypertension, unspecified |  |  |
| Vascular hypertension |  |  |
| White coat hypertension |  |  |
| White coat syndrome with diagnosis of hypertension |  |  |
| White coat syndrome with high blood pressure but without hypertension |  |  |
| White coat syndrome with high blood pressure without hypertension |  |  |
| White coat syndrome with hypertension |  |  |
| White coat syndrome without diagnosis of hypertension |  |  |
| White coat syndrome without hypertension |  |  |
