## Supplementary material for "Use of AI-Related CPT Codes by Patient, Geographic, and Payer Characteristics in the U.S., 2021-2026": eTable 1

**eTable 1.** Current Procedural Terminology (CPT) codes associated with artificial intelligence.

| **CPT code** | **CPT description** | **AMA Category** | **AI use** | **Appendix S classification** | **Care stage** | **In effect** | **Examples of Devices** | **Cosmos name** | **Grouper** |
| --- | --- | --- | --- | --- | --- | --- | --- | --- | --- |
| 20985 | Computer-assisted surgical navigational procedure for musculoskeletal procedures, image-less | 1 | Optional | Assistive | Treatment | 1/1/08 | Stryker Mako System | CPTR-ASST SURGICAL NAVIGATION IMAGE-LESS 20985 |  |
| 31627 | Bronchoscopy, rigid or flexible, including fluoroscopic guidance, when performed with computer-assisted, image-guided navigation, including 3D reconstruction | 1 | Optional | Assistive | Treatment | 1/1/10 | LungVision, Monarch, Ion Endoluminal System | BRONCHOSCOPY W/CPTR-ASST IMAGE-GUIDED NAVIGATION 31627 |  |
| 61781 | Stereotactic computer-assisted (navigational) procedure; cranial, intradural | 1 | Optional | Assistive | Treatment | 7/1/11 | Medivis Cranial Navigation | STRTCTC CPTR ASSTD PX CRANIAL INTRADURAL 61781 |  |
| 61782 | Stereotactic computer-assisted (navigational) procedure; cranial, extradural | 1 | Optional | Assistive | Treatment | 7/1/11 | Acclarent Balloon Sinuplasty | STRTCTC CPTR ASSTD PX EXTRADURAL CRANIAL 61782 |  |
| 61783 | Stereotactic computer-assisted (navigational) procedure; spinal | 1 | Optional | Assistive | Treatment | 7/1/11 | Medivis Spine Navigation | STEREOTACTIC COMPUTER ASSISTED PX SPINAL 61783 |  |
| 75577 | Quantification and characterization of coronary atherosclerotic plaque to assess severity of coronary disease, derived from augmentative software analysis of the data set from a coronary computed tomographic angiography, with interpretation and report by a physician or other qualified health care professional | 1 | Required | Augmentative | Screening | 1/1/26 | cvi42, Plaque | Not in Cosmos |  |
| 75580 | Noninvasive estimate of coronary fractional flow reserve (FFR) derived from augmentative software analysis of the data set from a coronary computed tomography angiography, with interpretation and report by a physician or other qualified health care professional | 1 | Required | Augmentative | Diagnosis | 1/1/24 | Cleerly ISCHEMIA, HeartFlow FFR-CT Analysis | N-INVAS EST C FFR AUGMNT SW ALYS CTA I&R PHY/QHP 75580 | 75580 |
| 77067 | Screening mammography, bilateral (2-view study of each breast), including computer-aided detection (CAD) when performed | 1 | Optional | Assistive | Screening | 1/1/17 | SenoClaire, Senographe Pristina | SCREENING MAMMOGRAPHY BI 2-VIEW BREAST INC CAD 77067 |  |
| 77280 | Therapeutic radiology simulation-aided field setting; simple | 1 | Optional | Assistive | Treatment | 1/1/91 | Varian Eclipse, Elekta ONE | THER RAD SIMULAJ-AIDED FIELD SETTING SIMPLE 77280 | 77280,77285,77290 |
| 77285 | Therapeutic radiology simulation-aided field setting; intermediate (eg, tangent screen, Autoplot, arc perimeter, or single stimulus level automated test, such as Octopus 3 or 7 equivalent) | 1 | Optional | Assistive | Treatment | 1/1/91 | Varian Eclipse, Elekta ONE | THER RAD SIMULAJ-AIDED FIELD SETTING INTERMED 77285 | 77280,77285,77290 |
| 77290 | Therapeutic radiology simulation-aided field setting; complex | 1 | Optional | Assistive | Treatment | 1/1/91 | Varian Eclipse, Elekta ONE | THER RAD SIMULAJ-AIDED FIELD SETTING COMPLEX 77290 | 77280,77285,77290 |
| 81504 | Oncology (tissue of origin), microarray gene expression profiling of >2,000 genes, utilizing formalin-fixed paraffin-embedded tissue, algorithm reported as tissue similarity scores | 1 | Optional | Augmentative | Diagnosis | 1/1/24 | The Jackson Laboratory CUP-AI-Dx | ONCOLOGY TISSUE OF ORIGIN SIMILAR SCOR ALGORITHM 81054 |  |
| 85027 | Blood count; complete (CBC), automated (Hgb, Hct, RBC, WBC and platelet count) | 1 | Optional | Augmentative | Diagnosis | 1/1/99 | CellaVision, Vision Hema, EasyCell, Techcyte | BLOOD COUNT COMPLETE AUTOMATED 85027 |  |
| 88175 | Cytopathology, cervical or vaginal (the Bethesda System); automated thin layer preparation, screening by automated system under physician supervision | 1 | Optional | Autonomous | Screening | 1/1/07 | ThinPrep, Surepath, Genius Digital Diagnosis System, PAPNET | CYTP C/V AUTO THIN LYR PREPJ SCR MNL RESCR PHYS 88175 |  |
| 92081 | Visual field examination, unilateral or bilateral, with interpretation and report; limited examination (eg, tangent screen, Autoplot, arc perimeter, or single stimulus level automated test, such as Octopus 3 or 7 equivalent) | 1 | Optional | Assistive | Screening | 1/1/96 | VF2000, VF3 | LIMITED VISUAL FIELD XM UNI/BI I&R 92081 |  |
| 92082 | Visual field examination, unilateral or bilateral, with interpretation and report; intermediate examination (eg, tangent screen, Autoplot, arc perimeter, or single stimulus level automated test, such as Octopus 3 or 7 equivalent) | 1 | Optional | Assistive | Screening | 1/1/96 | VF2000, VF3 | INTERMEDIATE VISUAL FIELD XM UNI/BI I&R 92082 |  |
| 92083 | Visual field examination, unilateral or bilateral, with interpretation and report; extended examination | 1 | Optional | Assistive | Screening | 1/1/96 | VF2000, VF3 | EXTENDED VISUAL FIELD XM UNI/BI I&R 92083 |  |
| 92227 | Imaging of retina for detection or monitoring of disease; with remote clinical staff review and report, unilateral or bilateral | 1 | Optional | Augmentative | Diagnosis | 1/1/11 | LumineticsCore (IDx-DR), EyeArt | IMG RETINA DETCJ/MNTR DS REM CLIN STAFF UNI/BI 92227 | 92227-92228 |
| 92228 | Imaging of retina for detection or monitoring of disease; with remote clinical staff review and report, unilateral or bilateral; point-of-care autonomous analysis and report, unilateral or bilateral | 1 | Optional | Augmentative | Diagnosis | 1/1/11 | LumineticsCore (IDx-DR), EyeArt | IMG RETINA DETCJ/MNTR DS REM PHYS/QHP I&R UNI/BI 92228 | 92227-92228 |
| 92229 | Imaging of retina for detection or monitoring of disease; with remote clinical staff review and report, unilateral or bilateral; point-of-care autonomous analysis and report, unilateral or bilateral | 1 | Required | Autonomous | Screening | 1/1/21 | LumineticsCore (IDx-DR), AEYE-DS | IMG RETINA DETCJ/MNTR DS POC AUTON A/R UNI/BI 92229 |  |
| 93356 | Myocardial strain imaging using speckle tracking-derived assessment of myocardial mechanics (List separately in addition to codes for echocardiography imaging) | 1 | Optional | Augmentative | Diagnosis | 1/1/20 | Ultromics EchoGo | MYOCRD STRAIN IMG SPECKLE TRCK ASSMT MYOCRD MECH 93356 | 93356 |
| 95919 | Quantitative pupillometry with physician or other qualified health care professional interpretation and report, unilateral or bilateral | 1 | Optional | Augmentative | Diagnosis | 1/1/23 | NeurOptics NPi-100, Carrot Visual Diagnostics Platform | QUANTITATIVE PUILLOMETRY PHYS/QHP I&R UNI/BI 95919 |  |
| 0866T | Quantitative magnetic resonance image (MRI) analysis of the brain with comparison to prior magnetic resonance (MR) study(ies), including lesion identification, characterization, and quantification, with brain volume(s) quantification and/or severity score, when performed, data preparation and transmission, interpretation and report, obtained without diagnostic MRI examination of the brain not in the same session; add-on | 3 | Required | Augmentative | Diagnosis | 1/1/24 | icometrix icobrain | QUAN MRI ALYS BRAIN WITH DIAGNOSTIC MRI 0866T |  |
| 0054T | Computer-assisted musculoskeletal surgical navigational orthopedic procedure, with image-guidance based on fluoroscopic images | 3 | Optional | Assistive | Treatment | 1/1/09 | Stryker Mako System, OrthoGrid Hip AI, Radlink | CPTR-ASST MUSCSKEL NAVIGJ ORTHO FLUOR IMAGES 0054T |  |
| 0055T | Computer-assisted musculoskeletal surgical navigational orthopedic procedure, with image-guidance based on CT/MRI images (List separately in addition to code for primary procedure) | 3 | Optional | Assistive | Treatment | 1/1/09 | Stryker Mako System | CPTR-ASST MUSCKEL NAVIGJ ORTHO CT/MRI 0055T |  |
| 0399T | Myocardial strain imaging (quantitative assessment of myocardial mechanics using image-based analysis of local myocardial dynamics) | 3 | Optional | Augmentative | Diagnosis | 1/1/16 | Epsilon Imaging EchoInsight | MYOCARDIAL STRAIN IMAGING QUAN ASSMT 0399T | 93356 |
| 0501T | Noninvasive estimated coronary FFR derived from coronary CTA data using computation fluid dynamics physiologic simulation software analysis of functional data to assess the severity of CAD; data preparation and transmission, analysis of fluid dynamics and simulated maximal coronary hyperemia, generation of estimated FFR model, with anatomical data review in comparison with estimated FFR model to reconcile discordant data, interpretation and report | 3 | Required | Augmentative | Diagnosis | 1/1/18 | Cleerly ISCHEMIA, HeartFlow FFR-CT Analysis | COR FFR DERIVED CTA DATA ASSESS COR ART DISEASE 0501T | 75580 |
| 0502T | Noninvasive estimated coronary FFR derived from coronary CTA data using computation fluid dynamics physiologic simulation software analysis of functional data to assess the severity of CAD; data preparation and transmission | 3 | Required | Augmentative | Diagnosis | 1/1/18 | Cleerly ISCHEMIA, HeartFlow FFR-CT Analysis | COR FFR DERIVED CTA DATA PREP & TRANSMIS 0502T | 75580 |
| 0503T | Noninvasive estimated coronary FFR derived from coronary CTA data using computation fluid dynamics physiologic simulation software analysis of functional data to assess the severity of CAD; analysis of fluid dynamics and simulated maximal coronary hyperemia, generation of estimated FFR model | 3 | Required | Augmentative | Diagnosis | 1/1/18 | Cleerly ISCHEMIA, HeartFlow FFR-CT Analysis | COR FFR CTA DATA ALYS & GNRJ ESTIMATED FFR MODEL 0503T | 75580 |
| 0504T | Noninvasive estimated coronary FFR derived from coronary CTA data using computation fluid dynamics physiologic simulation software analysis of functional data to assess the severity of CAD; with anatomical data review in comparison with estimated FFR model to reconcile discordant data, interpretation and report | 3 | Required | Augmentative | Diagnosis | 1/1/18 | Cleerly ISCHEMIA, HeartFlow FFR-CT Analysis | COR FFR CTA DATA REVIEW W/INTERPJ & FINAL REPORT 0504T | 75580 |
| 0604T | Optical coherence tomography (OCT) of retina, remote, patient-initiated image capture and transmission to a remote surveillance center, unilateral or bilateral; initial device provision, set-up and patient education on use of equipment | 3 | Required | Augmentative | Diagnosis | 7/1/20 | Notal Vision Inc. Home OCT | REMOTE OCT RETINA 1ST DEV SET-UP & PT EDUCAJ 0604T | 0604T-0606T |
| 0605T | Optical coherence tomography (OCT) of retina, remote, patient-initiated image capture and transmission to a remote surveillance center, unilateral or bilateral; remote surveillance center technical support, data analyses and reports, with a minimum of 8 daily recordings, each 30 days | 3 | Required | Augmentative | Diagnosis | 7/1/20 | Notal Vision Inc. Home OCT | REM OCT RETINA TECHL SUPRT MIN 8 DLY REC EA 30D 0605T | 0604T-0606T |
| 0606T | Optical coherence tomography (OCT) of retina, remote, patient-initiated image capture and transmission to a remote surveillance center, unilateral or bilateral; review, interpretation and report by the prescribing physician or other qualified health care professional of remote surveillance center data analyses, each 30 days | 3 | Required | Augmentative | Diagnosis | 7/1/20 | Notal Vision Inc. Home OCT | REMOTE OCT RETINA REVIEW I&R PHYS/QHP EA 30 D 0606T | 0604T-0606T |
| 0615T | Automated analysis of binocular eye movements without spatial calibration, including disconjugacy, saccades, and pupillary dynamics for the assessment of concussion, with interpretation and report | 3 | Required | Autonomous | Diagnosis | 7/1/20 | EyeBOX | AUTO ALYS BINOCULAR EYE MVMT ASSMT CONCUSSION 0615T |  |
| 0623T | Automated quantification and characterization of coronary atherosclerotic plaque to assess severity of coronary disease, using data from coronary computed tomographic angiography; data preparation and transmission, computerized analysis of data, with review of computerized analysis output to reconcile discordant data, interpretation and report | 3 | Required | Augmentative | Screening | 1/1/21 | Cleerly Coronary Analysis, Elucid PlaqueIQ, HeartFlow Plaque Analysis | AUTO QUAN&CHARAC CORONARY ATHEROSCLEROTIC PLAQUE 0623T | 0623T-0626T |
| 0624T | Automated quantification and characterization of coronary atherosclerotic plaque to assess severity of coronary disease, using data from coronary computed tomographic angiography; data preparation and transmission, computerized analysis of data, with review of computerized analysis output to reconcile discordant data, data preparation and transmission | 3 | Required | Augmentative | Screening | 1/1/21 | Cleerly Coronary Analysis, Elucid PlaqueIQ, HeartFlow Plaque Analysis | AUTO QUAN&CHARAC CORONARY PLAQ COMPUTERIZED ALYS 0625T | 0623T-0626T |
| 0625T | Automated quantification and characterization of coronary atherosclerotic plaque to assess severity of coronary disease, using data from coronary computed tomographic angiography; data preparation and transmission, computerized analysis of data, with review of computerized analysis output to reconcile discordant data, computerized analysis of data from coronary computed tomographic angiography | 3 | Required | Augmentative | Screening | 1/1/21 | Cleerly Coronary Analysis, Elucid PlaqueIQ, HeartFlow Plaque Analysis | AUTO QUAN&CHARAC CORONARY PLAQ DATA PREP&TRNSMIS 0624T | 0623T-0626T |
| 0626T | Automated quantification and characterization of coronary atherosclerotic plaque to assess severity of coronary disease, using data from coronary computed tomographic angiography; data preparation and transmission, computerized analysis of data, with review of computerized analysis output to reconcile discordant data, review of computerized analysis output to reconcile discordant data, interpretation and report | 3 | Required | Augmentative | Screening | 1/1/21 | Cleerly Coronary Analysis, Elucid PlaqueIQ, HeartFlow Plaque Analysis | AUTO QUAN&CHARAC CORONARY PLAQ REV CPTR ALYS I&R 0626T | 0623T-0626T |
| 0648T | Quantitative magnetic resonance for analysis of tissue composition (eg, fat, iron, water content), including multiparametric data acquisition, data preparation and transmission, interpretation and report, obtained without diagnostic MRI examination of the same anatomy (eg, organ, gland, tissue, target structure) during the same session; single organ | 3 | Optional | Augmentative | Diagnosis | 7/1/21 | Cortechs.ai OnQ Prostate | QUAN MR ALYS TISS COMPJ W/O MRI SAME SESS 1ORGN 0648T |  |
| 0649T | Quantitative magnetic resonance for analysis of tissue composition (eg, fat, iron, water content), including multiparametric data acquisition, data preparation and transmission, interpretation and report, obtained with diagnostic MRI examination of the same anatomy (eg, organ, gland, tissue, target structure); single organ | 3 | Optional | Augmentative | Diagnosis | 7/1/21 | Cortechs.ai OnQ Prostate | QUAN MR ALYS TISS COMPOSITION W/MRI 1ORGN 0649T |  |
| 0689T | Quantitative ultrasound tissue characterization (non-elastographic), including interpretation and report, obtained without diagnostic ultrasound examination of the same anatomy (eg, organ, gland, tissue, target structure) | 3 | Optional | Augmentative | Diagnosis | 1/1/22 | Koios DS Breast, Koios DS Thyroid | QUAN US TISS CHARAC I&R W/O DX US SAME ANAT 0689T |  |
| 0690T | Quantitative ultrasound tissue characterization (non-elastographic), including interpretation and report, obtained with diagnostic ultrasound examination of the same anatomy (eg, organ, gland, tissue, target structure) | 3 | Optional | Augmentative | Diagnosis | 1/1/22 | Koios DS Breast, Koios DS Thyroid | QUANTITATIVE US TISS CHARACTER I&R W/DX US SM ANAT 0690T |  |
| 0691T | Automated analysis of an existing computed tomography study for vertebral fracture(s), including assessment of bone density when performed, data preparation, interpretation, and report | 3 | Required | Autonomous | Diagnosis | 1/1/22 | Nanox.AI HealthOST | Not in Cosmos |  |
| 0694T | 3-dimensional volumetric imaging and reconstruction of breast or axillary lymph node tissue, each excised specimen, 3-dimensional automatic specimen reorientation, interpretation and report, real-time intraoperative | 3 | Optional | Assistive | Diagnosis | 1/1/22 | Clarix, MOZART iQ, IzoView | 3D VOLUMETRIC IMG&RCNSTJ BRST/AX LYMPH NODE TISS 0694T |  |
| 0697T | Quantitative magnetic resonance for analysis of tissue composition (eg, fat, iron, water content), including multiparametric data acquisition, data preparation and transmission, interpretation and report, obtained without diagnostic MRI examination of the same anatomy (eg, organ, gland, tissue, target structure) during the same session; multiple organs | 3 | Optional | Augmentative | Diagnosis | 1/1/22 | Perspectum CoverScan | QUAN MR ALYS TIS COMPJ WO MRI SAME SESS MLT ORGN 0697T |  |
| 0698T | Quantitative magnetic resonance for analysis of tissue composition (eg, fat, iron, water content), including multiparametric data acquisition, data preparation and transmission, interpretation and report, obtained with diagnostic MRI examination of the same anatomy (eg, organ, gland, tissue, target structure); multiple organs | 3 | Optional | Augmentative | Diagnosis | 1/1/22 | Perspectum CoverScan | QUAN MR ALYS TISS COMPOSITION W/MRI MLT ORGANS 0698T |  |
| 0710T | Noninvasive arterial plaque analysis using software processing of data from non-coronary computerized tomography angiography; including data preparation and transmission, quantification of the structure and composition of the vessel wall and assessment for lipid-rich necrotic core plaque to assess atherosclerotic plaque stability, data review, interpretation and report | 3 | Required | Augmentative | Diagnosis | 1/1/22 | Elucid PlaqueIQ | Not in Cosmos |  |
| 0711T | Noninvasive arterial plaque analysis using software processing of data from non-coronary computerized tomography angiography; data preparation and transmission | 3 | Required | Augmentative | Diagnosis | 1/1/22 | Elucid PlaqueIQ | N-INVAS ARTL PLAQ ALYS DATA PREP & TRANSMISSION 0711T |  |
| 0712T | Noninvasive arterial plaque analysis using software processing of data from non-coronary computerized tomography angiography; quantification of the structure and composition of the vessel wall and assessment for lipid-rich necrotic core plaque to assess atherosclerotic plaque stability | 3 | Required | Augmentative | Diagnosis | 1/1/22 | Elucid PlaqueIQ | N-INVAS ARTL PLAQ ALYS QUAN STRUX&COMPOS VSL WAL 0712T |  |
| 0713T | Noninvasive arterial plaque analysis using software processing of data from non-coronary computerized tomography angiography; data review, interpretation and report | 3 | Required | Augmentative | Diagnosis | 1/1/22 | Elucid PlaqueIQ | Not in Cosmos |  |
| 0716T | Cardiac acoustic waveform recording with automated analysis and generation of coronary artery disease risk score | 3 | Required | Augmentative | Diagnosis | 7/1/22 | Acarix CADScor | CARDIAC ACOUS WAVFRM REC AUTO ALYS CAD RSK SCORE 0716T |  |
| 0721T | Quantitative computed tomography (CT) tissue characterization, including interpretation and report, obtained without concurrent CT examination of any structure contained in previously acquired diagnostic imaging | 3 | Optional | Augmentative | Diagnosis | 7/1/22 | Optellum Lung Cancer Prediction | QUAN CT TISS CHARAC I&R W/O CNCRNT CT EXAM 0721T |  |
| 0722T | Quantitative computed tomography (CT) tissue characterization, including interpretation and report, obtained with concurrent CT examination of any structure contained in the concurrently acquired diagnostic imaging dataset | 3 | Optional | Augmentative | Diagnosis | 7/1/22 | Optellum Lung Cancer Prediction, Qure.ai qCT LN Quant | QUAN CT TISS CHARACTER I&R W/CNCRNT CT EXAM 0722T |  |
| 0723T | Quantitative magnetic resonance cholangiopancreatography (QMRCP) including data preparation and transmission, interpretation and report, obtained without diagnostic magnetic resonance imaging (MRI) examination of the same anatomy (eg, organ, gland, tissue, target structure) during the same session | 3 | Optional | Augmentative | Diagnosis | 7/1/22 | Optellum Lung Cancer Prediction, Qure.ai qCT LN Quant | QMRCP W/O DIAGNOSTIC MRI SM ANATOMY DRG SM SESS 0723T |  |
| 0724T | Quantitative magnetic resonance cholangiopancreatography (QMRCP), including data preparation and transmission, interpretation and report, obtained with diagnostic magnetic resonance imaging (MRI) examination of the same anatomy (eg, organ, gland, tissue, target structure) (List separately in addition to code for primary procedure) | 3 | Optional | Augmentative | Diagnosis | 7/1/22 | Optellum Lung Cancer Prediction, Qure.ai qCT LN Quant | QMRCP W/DIAGNOSTIC MRI SAME ANATOMY 0724T |  |
| 0731T | Augmentative AI-based facial phenotype analysis with report | 3 | Required | Augmentative | Screening | 7/1/22 | FACE2GENE | Not in Cosmos |  |
| 0740T | Remote autonomous algorithm-based recommendation system for insulin dose calculation and titration; initial set-up and patient education | 3 | Required | Autonomous | Treatment | 1/1/23 | Hygieia’s d-Nav | REM AUTON ALG INSULIN DOSE 1ST SETUP& PT EDUCAJ 0740T |  |
| 0741T | Remote autonomous algorithm-based recommendation system for insulin dose calculation and titration; initial set-up and patient education; provision of software, data collection, transmission, and storage, each 30 days | 3 | Required | Autonomous | Treatment | 1/1/23 | Hygieia’s d-Nav | Not in Cosmos |  |
| 0749T | Bone strength and fracture-risk assessment using digital X-ray radiogrammetry-bone mineral density (DXR-BMD) analysis of bone mineral density (BMD) utilizing data from a digital X ray, retrieval and transmission of digital X-ray data, assessment of bone strength and fracture risk and BMD, interpretation and report | 3 | Required | Augmentative | Diagnosis | 1/1/23 | OsteoApp.ai | Not in Cosmos |  |
| 0750T | Bone strength and fracture-risk assessment using digital X-ray radiogrammetry-bone mineral density (DXR-BMD) analysis of bone mineral density (BMD) utilizing data from a digital X ray, retrieval and transmission of digital X-ray data, assessment of bone strength and fracture risk and BMD, interpretation and report, with single-view digitial X-ray examination of the hand taken for the purpose of DXR-BMD | 3 | Required | Augmentative | Diagnosis | 1/1/23 | OsteoApp.ai | Not in Cosmos |  |
| 0764T | Assistive algorithmic electrocardiogram risk-based assessment for cardiac dysfunction (eg, low-ejection fraction, pulmonary hypertension, hypertrophic cardiomyopathy); related to concurrently performed electrocardiogram (List separately in addition to code for primary procedure) | 3 | Required | Assistive | Screening | 1/1/23 | Viz HCM | ASSTV ALG ECG RSK-BASED ASSMT RELATED CNCRT ECG 0764T |  |
| 0765T | Assistive algorithmic electrocardiogram risk-based assessment for cardiac dysfunction (eg, low-ejection fraction, pulmonary hypertension, hypertrophic cardiomyopathy); related to previously performed electrocardiogram | 3 | Required | Assistive | Screening | 1/1/23 | Viz HCM | Not in Cosmos |  |
| 0777T | Real-time pressure-sensing epidural guidance system | 3 | Required | Assistive | Treatment | 1/1/23 | CompuFlo | R-T PRESSURE SENSING EPIDURAL GUIDANCE SYSTEM 0777T |  |
| 0791T | Motor-cognitive semi-immersive virtual reality facilitated gait training, each 15 minutes | 3 | Required | Autonomous | Treatment | 7/1/23 | Gaitbetter | MOTOR COG SEMI-IMMRSV VR FACIL GAIT TRAING EA 15 0791T |  |
| 0794T | Patient-specific, assistive, rules-based algorithm for ranking pharmaco-oncologic treatment options based on the patient's tumor-specific cancer marker information obtained from prior molecular pathology, immunohistochemical, or other pathology results which have been previously interpreted and reported separately | 3 | Required | Assistive | Treatment | 7/1/23 | Perthera.ai, CureMatch | Not in Cosmos |  |
| 0807T | Pulmonary tissue ventilation analysis using software-based processing of data from separately captured cinefluorograph images; in combination with previously acquired computed tomography (CT) images, including data preparation and transmission, quantification of pulmonary tissue ventilation, data review, interpretation and report; in combination with previously acquired computed tomography (CT) images, including data preparation and transmission, quantification of pulmonary tissue ventilation, data review, interpretation and report | 3 | Required | Assistive | Diagnosis | 7/1/23 | Optellum Lung Cancer Prediction | Not in Cosmos |  |
| 0808T | Pulmonary tissue ventilation analysis using software-based processing of data from separately captured cinefluorograph images; in combination with previously acquired computed tomography (CT) images, including data preparation and transmission, quantification of pulmonary tissue ventilation, data review, interpretation and report in combination with computed tomography (CT) images taken for the purpose of pulmonary tissue ventilation analysis, including data preparation and transmission, quantification of pulmonary tissue ventilation, data review, interpretation and report | 3 | Optional | Assistive | Diagnosis | 7/1/23 | XV Lung Ventilation Analysis | PULM TISS VNTJ ALYS IN CMBN CT F/PULM TISS VNTJ 0808T |  |
| 0857T | Opto-acoustic imaging, breast, unilateral, including axilla when performed, real-time with image documentation, augmentative analysis and report (List separately in addition to code for primary procedure) | 3 | Required | Augmentative | Diagnosis | 1/1/24 | Seno medical Imagio | OPTO-ACOUSTIC IMG BREAST UNI AUGMNT ALYS&REPRT 0857T |  |
| 0858T | Externally applied transcranial magnetic stimulation with concomitant measurement of evoked cortical potentials with automated report | 3 | Optional | Assistive | Diagnosis | 1/1/24 | QuantalX Delphi-MD | Not in Cosmos |  |
| 0865T | Quantitative magnetic resonance image (MRI) analysis of the brain with comparison to prior magnetic resonance (MR) study(ies), including lesion identification, characterization, and quantification, with brain volume(s) quantification and/or severity score, when performed, data preparation and transmission, interpretation and report, obtained without diagnostic MRI examination of the brain during the same session | 3 | Required | Augmentative | Diagnosis | 1/1/24 | icometrix icobrain | QUAN MRI ALYS BRAIN W/O DIAGNOSTIC MRI SAME SESS 0865T |  |
| 0877T | Augmentative analysis of chest computed tomography (CT) imaging data to provide categorical diagnostic subtype classification of interstitial lung disease; obtained without concurrent CT examination of any structure contained in previously acquired diagnostic imaging | 3 | Required | Augmentative | Diagnosis | 7/1/24 | ScreenDx, Brainomix e-Lung | AUGMNT ALYS CH CT IMG DATA ILD WO CNCRNT CT EXAM 0877T |  |
| 0878T | Augmentative analysis of chest computed tomography (CT) imaging data to provide categorical diagnostic subtype classification of interstitial lung disease; obtained without concurrent CT examination of any structure contained in previously acquired diagnostic imaging obtained with concurrent CT examination of the same structure | 3 | Required | Augmentative | Diagnosis | 7/1/24 | ScreenDx, Brainomix e-Lung | AUGMNT ALYS CH CT IMG DATA ILD W/CNCRNT CT EXAM 0878T |  |
| 0879T | Augmentative analysis of chest computed tomography (CT) imaging data to provide categorical diagnostic subtype classification of interstitial lung disease; obtained without concurrent CT examination of any structure contained in previously acquired diagnostic imaging; radiological data preparation and transmission | 3 | Required | Augmentative | Diagnosis | 7/1/24 | ScreenDx, Brainomix e-Lung | AUGMNT ALYS CH CT IMG DATA ILD DATA PREP&TRNSMS 0879T |  |
| 0880T | Augmentative Analysis Of Chest Computed Tomography (CT) Data for ILD; interpretation and report | 3 | Required | Augmentative | Diagnosis | 7/1/24 | Imvaria Fibresolve | Not in Cosmos |  |
| 0897T | Noninvasive augmentative arrhythmia analysis derived from quantitative computational cardiac arrhythmia simulations, based on selected intervals of interest from 12-lead electrocardiogram and uploaded clinical parameters, including uploading clinical parameters with interpretation and report | 3 | Required | Augmentative | Screening | 7/1/24 | Vektor Medical vMap | N-INVAS AUGMNT ARRHYT ALYS QUAN CAR ARRHYT SIMUL 0897T |  |
| 0898T | Noninvasive prostate cancer estimation map, derived from augmentative analysis of image-guided fusion biopsy and pathology, including visualization of margin volume and location, with margin determination and physician interpretation and report | 3 | Required | Augmentative | Diagnosis | 7/1/24 | Unfold AI, SmartTarget | NONINVASIVE PROSTATE CANCER ESTIMATION MAP 0898T |  |
| 0899T | Noninvasive determination of absolute quantitation of myocardial blood flow (AQMBF), derived from augmentative algorithmic analysis of the dataset acquired via contrast cardiac magnetic resonance (CMR), pharmacologic stress, with interpretation and report by a physician or other qualified health care professional | 3 | Required | Augmentative | Diagnosis | 7/1/24 | cvi42 | Not in Cosmos |  |
| 0900T | Noninvasive estimate of absolute quantitation of myocardial blood flow (AQMBF), derived from assistive algorithmic analysis of the dataset acquired via contrast cardiac magnetic resonance (CMR), pharmacologic stress, with interpretation and report by a physician or other qualified health care professional (List separately in addition to code for primary procedure) | 3 | Required | Assistive | Diagnosis | 1/1/25 | cvi42 | Not in Cosmos |  |
| 0902T | Algorithmic analysis derived QTc Interval | 3 | Required | Augmentative | Diagnosis | 1/1/25 | BodyGuardian, KardiaMobile | Not in Cosmos |  |
| 0932T | Noninvasive detection of heart failure derived from augmentative analysis of an echocardiogram that demonstrated preserved ejection fraction, with interpretation and report by a physician or other qualified health care professional | 3 | Required | Augmentative | Diagnosis | 1/1/25 | Ultromics EchoGo Heart Failure | N-INVAS DETCJ HEART FAILURE DRV AUGMNT ALYS ECHO 0932T |  |
| 0962T | Assistive algorithmic analysis of acoustic and electrocardiogram recording for detection of cardiac dysfunction (eg, reduced ejection fraction, cardiac murmurs, atrial fibrillation), with review and interpretation by a physician or other qualified health care professional | 3 | Required | Assistive | Diagnosis | 7/1/25 | Eko SENSORA | Not in Cosmos |  |
| 0972T | Assistive algorithmic classification of burn healing (ie, healing or nonhealing) by noninvasive multispectral imaging, including system set-up and acquisition, selection, and transmission of images, with automated generation of report | 3 | Required | Assistive | Diagnosis | 7/1/25 | Spectral AI Deep View, MIMOSA Diagnostics MIMOSA Pro | Not in Cosmos |  |
| 0992T | Noninvasive assessment of cardiac risk derived from augmentative software analysis of perivascular fat without concurrent computed tomography (CT) scan of the heart, including patient-specific clinical factors, with interpretation and report by a physician or other qualified health care professional | 3 | Required | Augmentative | Screening | 1/1/26 | CaRi-Heart | Not in Cosmos |  |
| 0993T | Noninvasive assessment of cardiac risk derived from augmentative software analysis of perivascular fat with concurrent computed tomography scan of the heart, including patient-specific clinical factors, with interpretation and report by a physician or other qualified health care professional (List separately in addition to code for primary procedure) | 3 | Required | Augmentative | Screening | 1/1/26 | CaRi-Heart | Not in Cosmos |  |
| 1020T | Raman spectroscopy of 1 or more skin lesions, with probability score for malignant risk derived by algorithmic analysis of data from each lesion | 3 | Optional | Augmentative | Screening | 1/1/26 | Vita Imaging AURA system | Not in Cosmos |  |
| 1036T | Noninvasive hemodynamic assessment with pulmonary pressures and ejection fraction when performed, including passive acquisition of acoustic and electrical signals, augmentative algorithmic analysis, and generation of a clinical report with review, interpretation, and clinical integration by a physician or other qualified health care professional | 3 | Required | Augmentative | Screening | 7/1/26 | Sensydia's Cardiac Performance System | Not in Cosmos |  |
| 1041T | Augmentative algorithmic analysis of encephalographic waveforms to identify the source and propagation of epileptiform activity, including artifact reduction with analysis of 3D localization of spike sources throughout the examination, 3D animations over time of high-amplitude event locations, high-frequency activity locations, and temporal relationships among locations, with interpretation and report by physician or other qualified health care professional, related to a previously performed electroencephalogram (EEG) | 3 | Required | Augmentative | Diagnosis | 7/1/26 | autoSCORE | Not in Cosmos |  |
| 85025 | Blood count; complete (CBC), automated (Hgb, Hct, RBC, WBC and platelet count) and automated differential WBC count | 1 | Optional | Augmentative | Diagnosis | 1/1/99 | CellaVision, Vision Hema, EasyCell, Techcyte | BLOOD COUNT COMPLETE AUTO&AUTO DIFRNTL WBC 85025 |  |
